# Regional brain-age gaps reveal a shared polygenic architecture and regionally patterned clinical and cell-type associations

**DOI:** 10.64898/2026.11.19.26367314

**Authors:** Yein Hwang, Kyung-Il Park, Seunggeun Lee, the Alzheimer’s Disease Neuroimaging Initiative

## Abstract

A single brain-age estimate may obscure regional differences in clinical vulnerability. Using voxel-level deep learning, we derived whole-brain and nine regional brain-age gaps (BAGs) spanning cortical, subcortical and cerebellar regions from T1-weighted MRI in 46,254 UK Biobank participants. Genetic correlation estimates in 41,067 participants (*r*_g_ = 0.56–0.93) exceeded phenotypic correlations across all 45 pairs, yet regional profiles varied across 24 hospital-recorded diagnoses. Oligodendrocyte annotations predominated for whole-brain and cerebellar BAGs, whereas annotations for caudate BAG included astrocyte and vascular classes. In the Alzheimer’s Disease Neuroimaging Initiative, temporal BAG showed stronger associations with cerebrospinal fluid Aβ42 and cognition than whole-brain BAG when modelled together. A regional model modestly improved discrimination of observed conversion from mild cognitive impairment to Alzheimer’s disease during available follow-up compared with a whole-brain model. Polygenic scores showed cross-cohort associations with hypertension and osteoporosis. Regional BAGs provide complementary clinical information within a shared polygenic architecture.

## Introduction

Brain ageing is a spatiotemporally heterogeneous process characterized by region-specific variability in the timing and magnitude of structural changes across the lifespan^1–3^. Brain-age models estimate an individual’s apparent brain age from neuroimaging and summarize deviation from chronological age as the brain-age gap (BAG)^4–6^. A higher BAG has been associated with mortality, mild cognitive impairment (MCI) and dementia, Alzheimer’s disease (AD)-related biomarkers, and subsequent clinical decline^7–10^. A single whole-brain BAG, however, may obscure regional variation in age-and disease-related brain changes^5,6,11–14^. Regional BAGs may therefore capture clinical and genetic information not represented by a whole-brain estimate.

Previous brain-age studies commonly used predefined morphometric features, such as cortical thickness and regional volumes, or voxelwise grey-matter measures with conventional machine-learning models^13,15–18^. Regional models have identified disorder-specific BAG patterns and provided evidence for the heritability of regional BAGs^13,18^. Deep-learning models can learn spatial features directly from three-dimensional MRI, although many still yield a single whole-brain age estimate^6,19,20^. Other studies have inferred regional contributions to global brain-age predictions using post hoc feature attribution^15,21,22^. Voxelwise local-age models provide another approach by generating spatially resolved age maps that can be averaged within anatomical regions to derive regional estimates^12,14,23,24^.

Genome-wide association studies (GWASs) of whole-brain BAG have identified associated loci and genetic relationships with neurological, psychiatric and systemic traits^25,26^. Multimodal studies have compared the genetic architecture and cell-type enrichment of grey-matter, white-matter and functional-connectivity BAGs^27^. Recent GWASs have also identified genetic associations with deep-learning-derived local brain age across cortical and subcortical regions^28,29^. However, the degree of genome-wide genetic sharing among whole-brain, cortical, subcortical and cerebellar BAGs remains incompletely characterized. Their potential to complement whole-brain BAG in clinical associations and cell-type enrichment, and their genetic relationships with leukocyte telomere length and proteomic age, also require clarification.

Here, we trained separate three-dimensional convolutional neural networks (3D-CNNs) to learn spatial features directly from whole-brain and atlas-masked regional T1-weighted MRI volumes in UK Biobank, yielding one whole-brain and nine regional BAG phenotypes. We assessed associations with hospital-recorded diagnoses in UK Biobank and with AD biomarkers and cognition in the Alzheimer’s Disease Neuroimaging Initiative (ADNI), where we also evaluated discrimination of observed MCI-to-AD conversion. We used these BAGs as quantitative phenotypes for GWAS and post-GWAS analyses and evaluated their polygenic scores in UK Biobank, All of Us and ADNI.

## Results

### Study overview

We combined imaging, clinical and genetic analyses across three cohorts (Fig. 1a–d). In the UK Biobank, we derived one whole-brain and nine regional BAG phenotypes from 46,254 imaging participants. Models were trained using four-fold cross-validation in 25,642 participants meeting prespecified disease-exclusion criteria and evaluated in an independent test set of 20,612 participants. BAG denotes bias-corrected predicted age minus chronological age; prediction accuracy was evaluated before bias correction (Methods). GWASs included 41,067 UK Biobank imaging participants with genotype data and self-reported White British ethnicity. Post-GWAS analyses included gene mapping, heritability and genetic correlation estimation, cell-type enrichment, polygenic risk score (PRS) derivation and validation, PRS-based phenome-wide association studies (PheWAS) in UK Biobank and All of Us and exploratory bidirectional Mendelian randomization. The framework was then applied to ADNI for domain-adapted clinical analyses, exploratory genetic concordance analyses and polygenic follow-up.

**Fig. 1:**
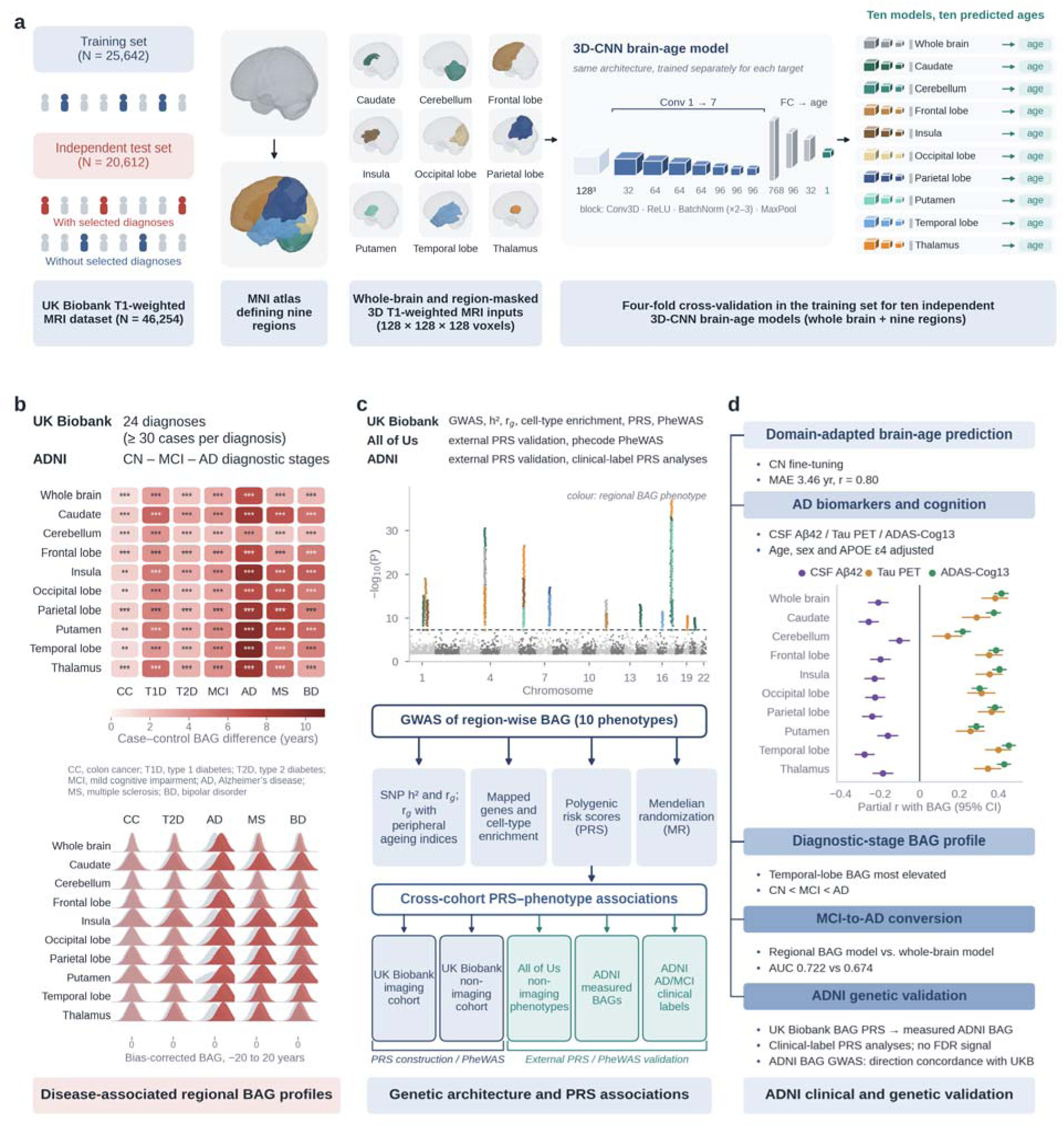
Study design for region-wise brain-age gap (BAG) analysis. **a**, Ten separate three-dimensional convolutional neural networks predicted age from whole-brain or atlas-masked regional T1-weighted MRI in UK Biobank (training, *n* = 25,642; independent testing, *n* = 20,612). **b**, Case–control BAG comparisons across 24 UK Biobank diagnoses and ADNI mild cognitive impairment (MCI) and Alzheimer’s disease (AD) groups. Controls were participants without selected diagnoses in UK Biobank (*n* = 10,776) and cognitively normal participants in ADNI. The heatmap and density plots summarize selected results (Fig. 2a; Supplementary Fig. 3); \**P* < 0.05, \*\**q* < 0.05 and \*\*\**q* < 0.005. **c,** Genome-wide association studies (GWASs), illustrated schematically, followed by heritability, genetic correlation, gene mapping, cell-type enrichment, PRS, PRS-based phenome-wide association studies (PheWAS) and exploratory Mendelian randomization analyses. PRS-based PheWAS was performed in UK Biobank and All of Us; ADNI analyses assessed PRS–BAG correlations and associations with AD and MCI diagnostic status. **d,** ADNI model fine-tuning, biomarker, cognitive and diagnostic-stage associations, observed MCI-to-AD conversion discrimination and PRS analyses.

### Region-wise brain-age prediction

We trained separate voxel-level 3D-CNNs for the whole brain and nine anatomical regions (Fig. 1a; Supplementary Fig. 1). In UK Biobank test participants without selected diagnoses (*n* = 10,776), the whole-brain model achieved a mean absolute error (MAE) of 2.39 years (Pearson *r* = 0.93), and regional MAEs ranged from 3.17 to 4.43 years (*r* = 0.73–0.87). Corresponding MAEs in participants with selected diagnoses (*n* = 9,836) were 2.51 and 3.29–4.45 years, with regional age correlations of *r* = 0.72–0.85 (Supplementary Table 1). In UK Biobank participants both with and without selected diagnoses, regional models showed lower MAEs than those reported in the regional studies summarized in Supplementary Table 1. Diagnosis-specific correlations with chronological age were generally higher than those reported by Kaufmann et al.^18^ (Supplementary Table 2). After fine-tuning in cognitively normal (CN) ADNI participants (*n* = 1,179), cross-validated MAEs were 3.46 years (*r* = 0.80) for the whole-brain model and 3.87–5.10 years (*r* = 0.49–0.75) for regional models (Supplementary Table 1). Diagnosis-specific correlations and 95% confidence intervals (CIs) are reported in Supplementary Table 2.

### Disease-associated regional BAG profiles

We compared BAGs between cases and controls across 24 hospital-recorded diagnoses in the independent UK Biobank test set, adjusting for age and sex (Methods; Supplementary Note 1). Nineteen of the 24 diagnoses were associated with higher BAG in at least one phenotype (*q* < 0.05, Benjamini–Hochberg (BH) correction across the ten BAG phenotypes separately within each diagnosis; Fig. 2a; Supplementary Fig. 3; Supplementary Table 3). All seven metabolic diagnoses were associated with higher BAG in at least nine of the ten phenotypes. Caudate BAG had the largest adjusted case–control difference among the ten BAGs for six of the seven metabolic diagnoses, including type 1 diabetes (5.91 years) and type 2 diabetes (3.31 years). Bipolar disorder, anxiety disorders and both addiction-related diagnoses were associated with higher BAG in all ten phenotypes. Multiple sclerosis showed the largest differences, reaching 7.52 years for parietal BAG.

**Fig. 2:**
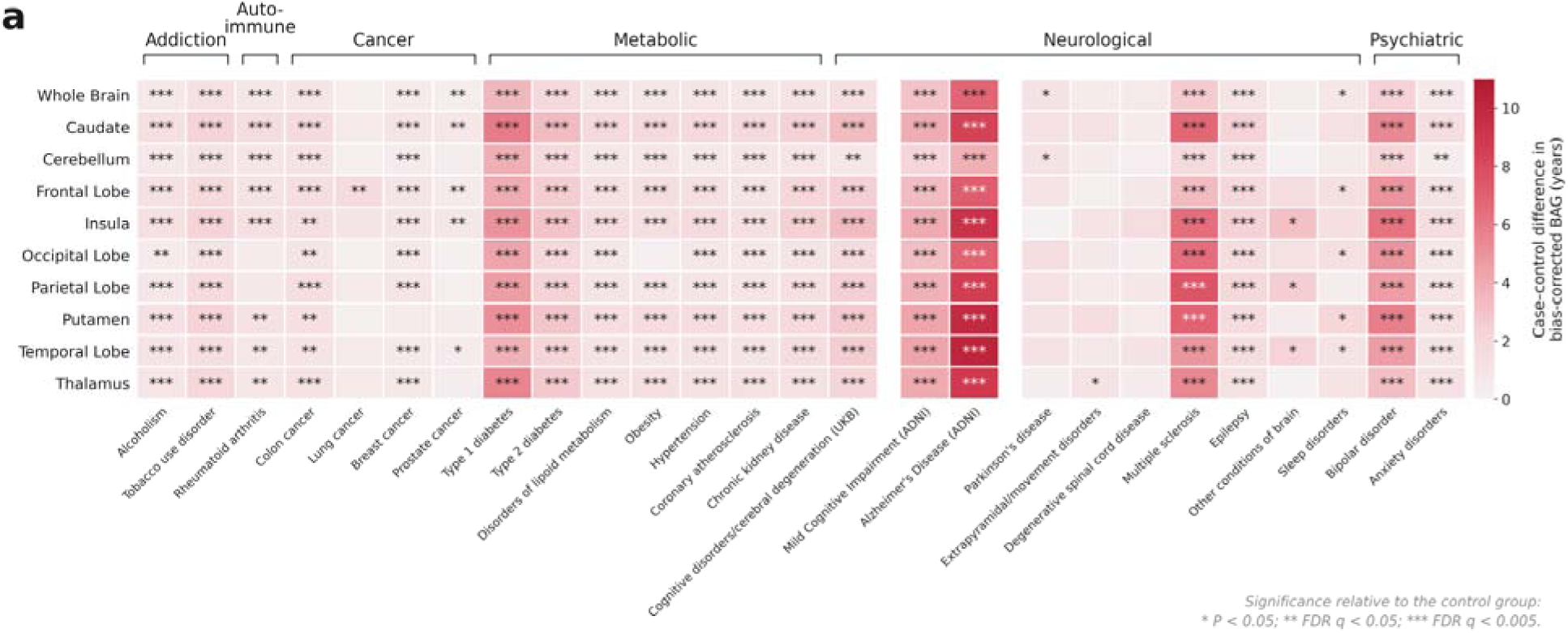
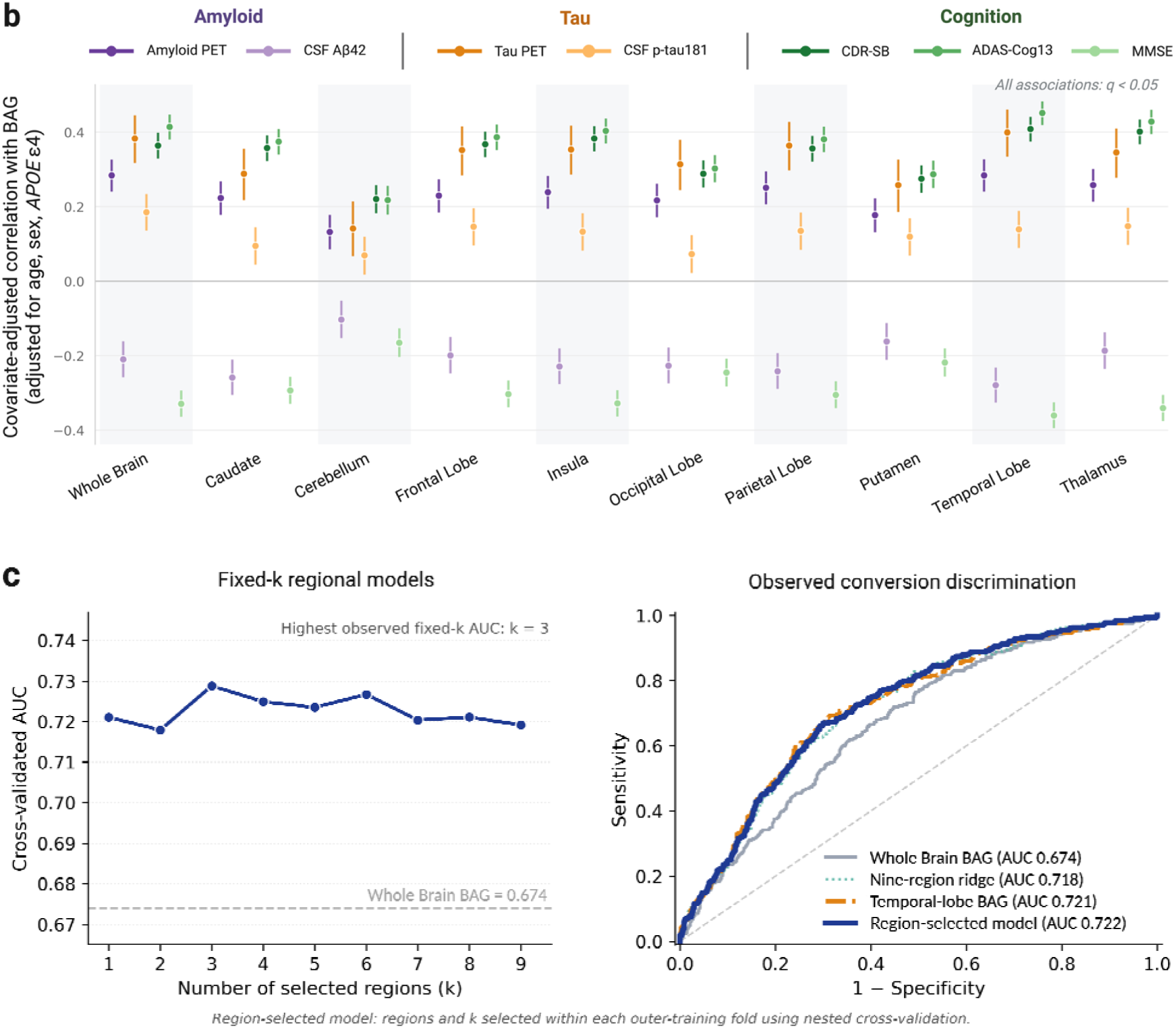
Clinical associations of whole-brain and regional brain-age gaps. **a**, Adjusted case–control differences in BAG years for 24 UK Biobank diagnoses and ADNI mild cognitive impairment (MCI; *n* = 1,232) or Alzheimer’s disease (AD; *n* = 398). Control were participants without selected diagnoses in UK Biobank (*n* = 10,776) and cognitively normal participants (*n* = 1,122) in ADNI. Models adjusted for age and sex, with MRI field strength additionally included in ADNI. Breast-and prostate-cancer analyses were sex-restricted and age-adjusted. Colours show year-scale differences, not standardized effects; negative estimates are displayed with the same colour as zero. Two-sided *P* values were Benjamini–Hochberg (BH)-corrected across ten BAGs within each diagnosis: \**P* < 0.05, \*\**q* < 0.05, \*\*\**q* < 0.005 (Supplementary Table 3). **b**, Correlations between BAGs and AD-related measures in ADNI, adjusted for age, sex and *APOE* ε4 allele count. Points and bars indicate correlation estimates and 95% confidence intervals, respectively. All associations were significant after BH correction across the 70 BAG–measure tests (*q* < 0.05). Tau positron emission tomography (PET) analyses used flortaucipir scans that passed quality control. Lower cerebrospinal fluid (CSF) Aβ42 and Mini-Mental State Examination (MMSE) scores indicate greater pathology or poorer cognition (Supplementary Table 4). **c,** Observed MCI-to-AD conversion discrimination (303 converters; 718 non-converters). *Left*, descriptive cross-validated areas under the receiver operating characteristic curve (AUCs) for fixed numbers of regions (k = 1–9); the dashed line shows whole-brain AUC. *Right*, ROC curves from pooled held-out predictions for the whole-brain, temporal-only, nine-region ridge and region-selected models. All models included age and sex; the region-selected model used nested cross-validation to select the regions and their number (k) (Supplementary Note 9; Supplementary Data 1).

We compared the adjusted case–control difference in each regional BAG with that in whole-brain BAG across 24 UK Biobank diagnoses (nine regions per diagnosis; 216 comparisons). When expressed in years, regional differences were significantly larger in 82 comparisons. After dividing each BAG by its control-group standard deviation, only parietal BAG in multiple sclerosis showed a significantly larger case–control difference than whole-brain BAG, whereas regional differences were significantly smaller in 58 comparisons (Supplementary Table 3, continued).

In ADNI, participants with MCI or AD had higher BAGs than cognitively normal participants across all ten phenotypes after adjustment for age, sex and MRI field strength (all q < 0.05; Fig. 2a; Supplementary Table 3). All ten BAGs were positively associated with baseline diagnostic stage (all *q* < 10^-27^), with the largest coefficient for temporal BAG (Supplementary Table 6).

### AD biomarkers and cognition

We assessed each BAG against seven AD-related measures: amyloid and flortaucipir tau positron emission tomography (PET), cerebrospinal fluid (CSF) amyloid-β 1–42 (Aβ42) and tau phosphorylated at threonine 181 (p-tau181), Clinical Dementia Rating–Sum of Boxes (CDR-SB), 13-item Alzheimer’s Disease Assessment Scale–Cognitive Subscale (ADAS-Cog13) and Mini-Mental State Examination (MMSE). Models adjusted for age, sex and APOE ε4 allele count. All 70 associations were significant after false discovery rate (FDR) correction, with higher BAGs associated with greater amyloid and tau burdens, lower CSF Aβ42, higher CSF p-tau181 and poorer cognitive performance (Fig. 2b; Supplementary Table 4).

Temporal BAG had the largest absolute coefficient in single-BAG models for six of the seven outcomes. A 1-s.d. higher temporal BAG was associated with 160.0 pg ml^-1^ lower CSF Aβ42 and a 4.16-point higher ADAS-Cog13 score (both *q* < 10^-24^; Supplementary Table 4).

For each outcome, we included temporal and whole-brain BAGs in the same regression model and compared their coefficients. Temporal BAG was more strongly associated with lower CSF Aβ42 (coefficient-contrast *q* = 2.0 × 10^-4^) and poorer performance on all three cognitive measures (*q* = 0.0019–0.032) than whole-brain BAG. Whole-brain BAG was more strongly associated with higher CSF p-tau181 than temporal BAG (*q* = 0.017), whereas their associations with amyloid and tau-PET did not differ significantly (Supplementary Table 5).

### Discrimination of observed MCI-to-AD conversion

We compared regional and whole-brain BAG models in 303 observed MCI-to-AD converters and 718 participants whose diagnosis remained MCI during follow-up (*n* = 1,021). The regional model achieved an area under the receiver operating characteristic curve (AUC) of 0.722 versus 0.674 for the whole-brain model (ΔAUC = 0.048; 95% bootstrap CI, 0.019–0.077). The temporal-only model had a similar AUC (0.721; Fig. 2c). The regional–whole-brain AUC difference was smaller under stricter follow-up criteria (ΔAUC = 0.034; 95% CI, −0.002 to 0.069; Supplementary Note 9; Supplementary Data 1).

### Shared and phenotype-restricted GWAS associations

GWASs of the ten BAG phenotypes in UK Biobank identified genome-wide significant associations across multiple chromosomes (Fig. 3a; Supplementary Fig. 4). Functional Mapping and Annotation of GWAS (FUMA) identified 125 distinct lead variants across these analyses; cross-phenotype locus merging yielded 64 genomic risk loci (Supplementary Table 25). Of the 64 loci, 43 were detected in only one BAG GWAS at *P* < 5 × 10^-8^, whereas 21 were detected in two or more (Supplementary Table 25). The strongest association was with thalamic BAG at 17q21.31 (rs2732689; *P* = 5.7 × 10^-38^). This locus overlaps the MAPT-containing inversion region, which has been associated with brain structure and neurodegeneration^30,31^. The region was associated with all ten BAGs and accounted for approximately 85% of genome-wide significant variant–phenotype pairs (Supplementary Table 25). A locus at 6p21.1, annotated to *RUNX2*, reached genome-wide significance for caudate, putaminal, thalamic and whole-brain BAGs. Genome-wide significant associations were also observed in the extended major histocompatibility complex (MHC) region for frontal, insular and temporal BAGs (Supplementary Table 25, continued).

**Fig. 3:**
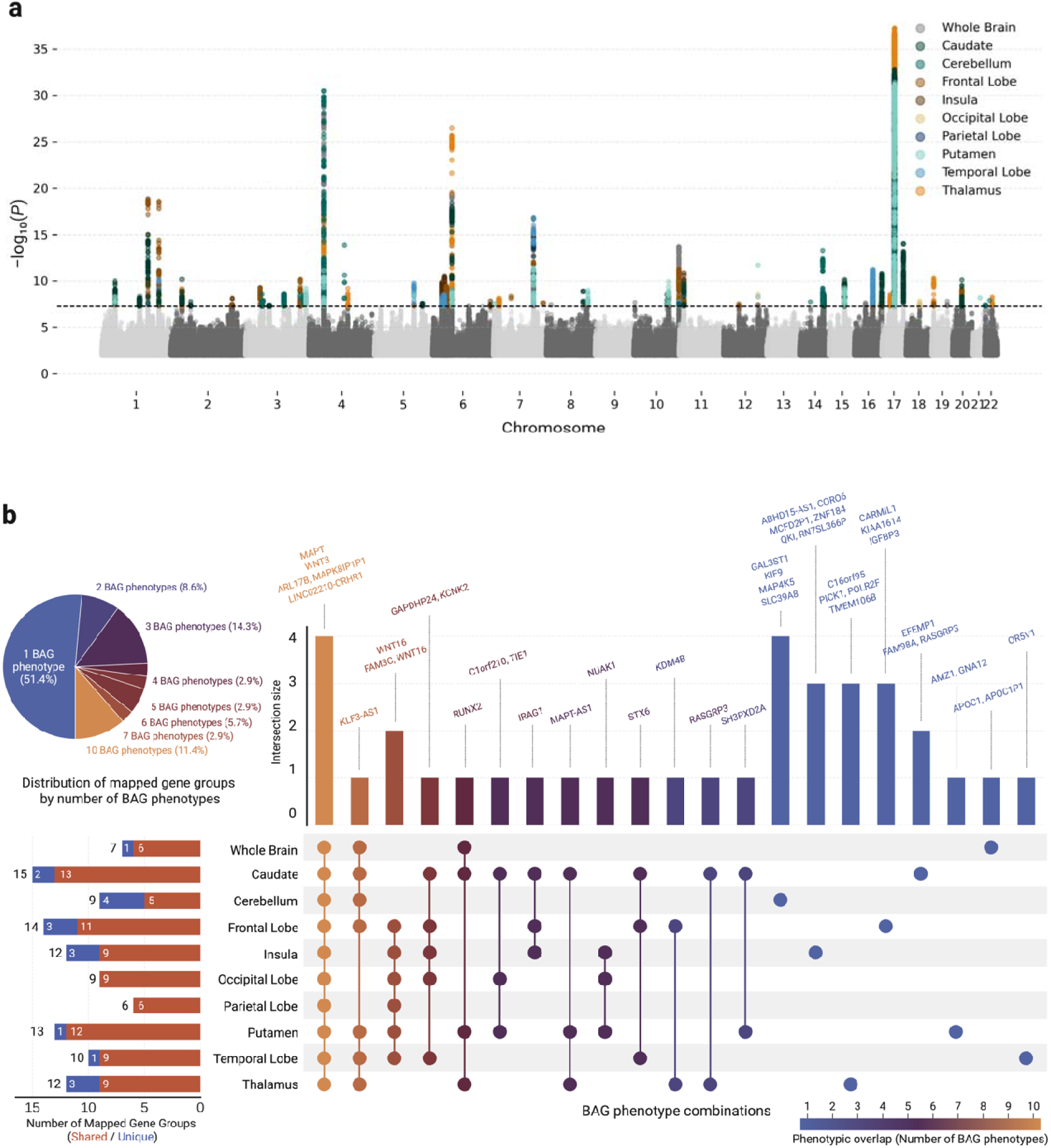
BAG GWAS associations and overlap of mapped gene groups. a, GWAS results for ten BAGs in 41,067 UK Biobank participants. Grey points show associations with *P* < 0.01; genome-wide significant associations are coloured by BAG. The dashed line marks *P* = 5 × 10^-8^ (two-sided; Supplementary Fig. 4). **b,** Overlap of 35 GWAS Catalog mapped gene groups. Assignment to each BAG used annotated variants from the pooled lead set present in that BAG’s FUMA candidate-variant results (Supplementary Note 10). The pie chart and intersection colours indicate the number of BAGs sharing a group. Vertical bars count groups in the combinations marked by connected dots; horizontal bars separate groups assigned to one BAG (unique) from those assigned to multiple BAGs (shared). Labels identify representative groups, which are gene annotations rather than independent loci (Supplementary Table 11).

Of the 125 lead variants, 42 had GWAS Catalog mapped-gene annotations, defining 35 mapped gene groups (Supplementary Table 11). These groups comprise one or more annotated genes, not independent loci or established causal genes. A group was assigned to a BAG when an annotated variant from the pooled lead set appeared in that BAG’s FUMA candidate-variant results (Supplementary Note 10). Eighteen groups were assigned to only one BAG. Four mapped gene groups—*WNT3, MAPT, LINC02210-CRHR1* and *ARL17B/MAPK8IP1P1—*were detected in all ten BAGs and arose from the same extended 17q21.31 locus.

Mapped gene profiles also varied across BAG phenotypes (Fig. 3b; Supplementary Table 11). Caudate BAG yielded the most mapped gene groups, followed by the frontal lobe and putamen; the occipital and parietal BAGs yielded no groups detected in that phenotype alone; and four of the nine cerebellar groups were detected only for cerebellar BAG, the highest such proportion across BAG phenotypes, consistent with reported differences between cerebellar and cerebral ageing^32,33^.

We assigned 444 distinct GWAS Catalog reported traits to 27 curated Experimental Factor Ontology (EFO) categories across the mapped gene groups (Supplementary Table 12). Neuroimaging traits were the most frequent annotations in both phenotype-restricted and shared groups. Cardiometabolic annotations were concentrated in phenotype-restricted groups, largely reflecting SLC39A8 (Supplementary Fig. 7; Supplementary Note 3; Supplementary Table 13).

### Shared polygenic architecture and cell-type enrichment profiles across BAGs

Single-nucleotide polymorphism (SNP)-based heritability was significant for all ten BAG phenotypes. Across the nine regional BAG phenotypes, the estimates ranged from 0.105 for the occipital lobe BAG to 0.184 for the caudate BAG, with a mean of 0.136. The whole-brain BAG showed a comparable estimate (h^2^ = 0.115, S.E. = 0.018; Supplementary Table 14). Only caudate BAG had significantly higher heritability than whole-brain BAG after BH correction across nine comparisons (difference=0.069; S.E.=0.016; q = 1.1 × 10^-4^; Supplementary Table 14, continued).

All 45 pairwise genetic correlations among the ten BAG phenotypes were positive (*r*_g_ = 0.564–0.927) and significant after FDR correction. Correlations were particularly high among the frontal, insular, occipital, parietal, putaminal and temporal BAGs (*r*_g_ = 0.790–0.927). Genetic correlation estimates exceeded the corresponding phenotypic correlations (*r* = 0.335–0.601) in every pair (Fig. 4a; Supplementary Table 15).

**Fig. 4:**
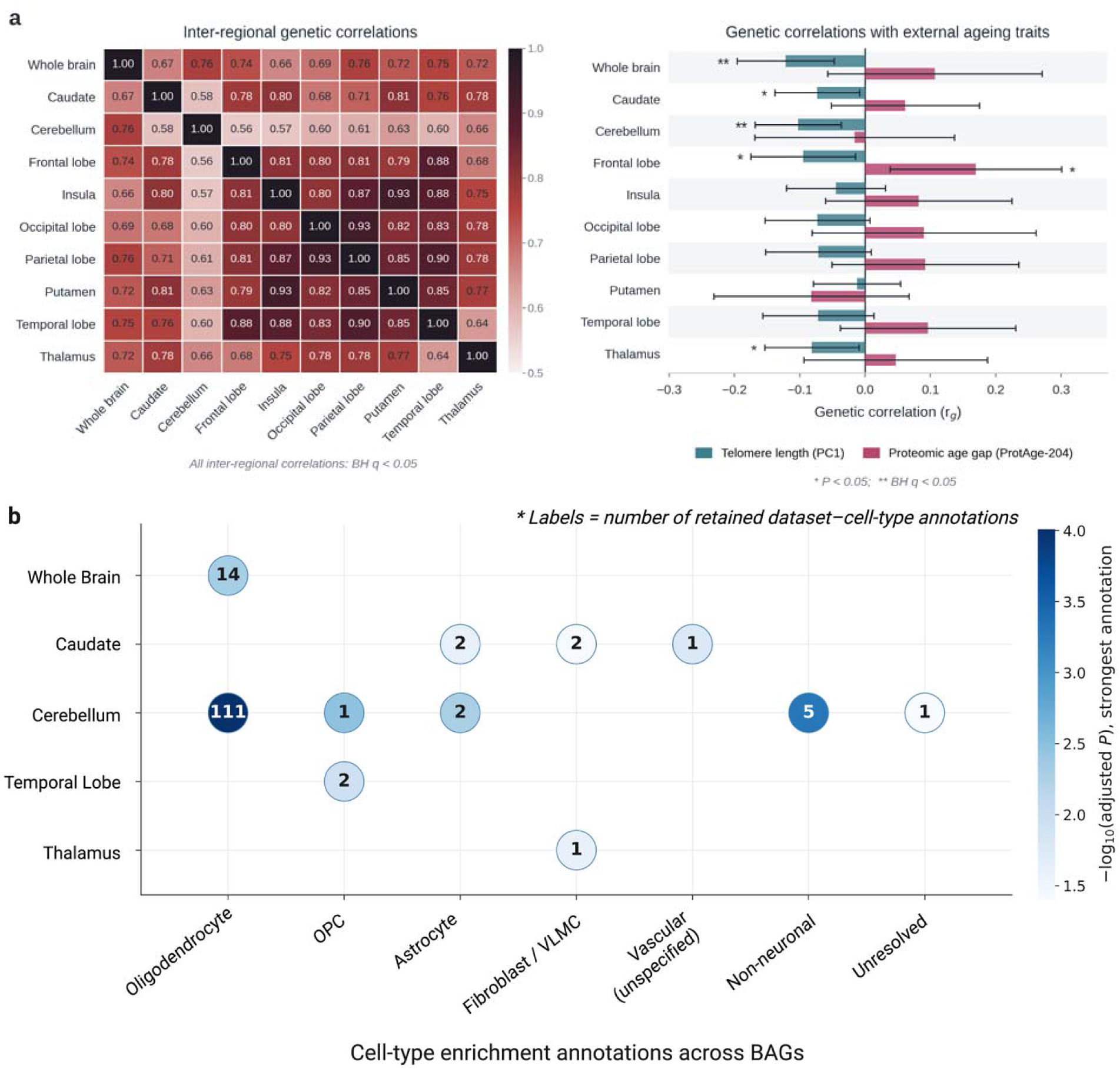
Genetic correlations and cell-type enrichment of BAGs. a, *Left*, pairwise genetic correlations (*r*_g_) among ten BAGs, estimated by cross-trait LD Score regression in 41,067 UK Biobank participants. *Right*, correlations with leukocyte telomere length PC1 and the ProtAge-204 proteomic-age gap; bars indicate 95% confidence intervals. Two-sided P values were Benjamini–Hochberg-corrected separately across 45 BAG–BAG correlations and 21 external-ageing correlations. \**P* < 0.05; \*\**q* < 0.05. The ProtAge-204–telomere correlation belongs to the latter family but is not plotted (Supplementary Table 15). **b,** Retained cell-type annotations from one-sided MAGMA gene-property tests, Bonferroni-corrected within each BAG. Labels show retained annotation counts; bubble colour indicates −log_10_ of the smallest Bonferroni-adjusted marginal *P* value per BAG–cell-class combination. Only retained combinations are shown. Counts represent dataset annotations, not independent cell populations; unspecified and unresolved classes preserve source-label resolution (Supplementary Note 7; Supplementary Table 16). OPC, oligodendrocyte precursor cell; VLMC, vascular leptomeningeal cell.

Genetic correlations with the first principal component (PC1) of leukocyte telomere length measurements^34^ were negative for all ten BAGs (*r*_g_ = −0.122 to −0.013), with those for whole-brain and cerebellar BAGs remaining significant after BH correction across 21 external-ageing tests (*q* = 0.014 and 0.016, respectively). In contrast, the ProtAge-204 proteomic-age gap^35^ showed positive genetic correlation estimates with eight of the ten BAGs (*r*_g_ = −0.082 to 0.169), although none survived after FDR correction (smallest *q* = 0.061, frontal BAG; Fig. 4a). ProtAge-204 and telomere length PC1 were inversely genetically correlated (*r*_g_ = −0.189, *q* = 0.0012; Supplementary Note 6; Supplementary Table 15).

FUMA cell-type enrichment analysis identified 173 significant BAG–annotation associations across five BAGs after Bonferroni correction within each BAG. Of these, 142 met the within-dataset retention criteria (Methods; Supplementary Table 16).

Oligodendrocyte annotations accounted for all 14 whole-brain associations and 111 of 120 cerebellar associations. Caudate BAG retained five annotations: two astrocyte, two fibroblast/vascular leptomeningeal cell (VLMC) and one vascular annotation without subtype resolution. Temporal BAG retained two oligodendrocyte precursor cell (OPC) annotations, and thalamic BAG retained one fibroblast/VLMC annotation. No annotations were retained for frontal, parietal, occipital, insular or putaminal BAGs, or for neuronal or pericyte/mural classes (Fig. 4b; Supplementary Table 16).

### Cross-cohort associations of BAG polygenic risk scores

We derived ten BAG PRS using SBayesRC and GWAS summary statistics from 30,800 White British UK Biobank imaging participants (Methods). All ten PRS were positively associated with their corresponding measured BAGs in the non-overlapping held-out UK Biobank sample (*n* = 9,997). In univariable models, adjusted R² ranged from 1.37% for occipital BAG to 3.83% for caudate BAG (Supplementary Table 17; Supplementary Fig. 8).

We conducted a PRS-based PheWAS in three relatedness-pruned samples excluded from PRS derivation: White British UK Biobank non-imaging (*n* = 359,334) and held-out imaging (*n* = 9,997) participants, and European-ancestry All of Us participants with complete covariates (*n* = 109,774; Supplementary Table 24). At *q* < 0.05 after BH correction within each PRS and sample, the non-imaging analysis identified 191 associations across 84 phecodes; 185 associations linked higher PRS to higher disease odds. In All of Us, we identified 14 significant regional BAG PRS–phecode associations involving eight phecodes, all linking higher PRS to higher disease odds (Fig. 5; Supplementary Tables 18–20). Nine regional PRS–phecode associations were significant in both the UK Biobank non-imaging and All of Us samples, with concordant positive directions, spanning hypertension, essential hypertension and osteoporosis (Supplementary Tables 18–20). In the non-imaging sample, higher PRS for five BAGs were associated with higher odds of type 2 diabetes (*q* = 2.1 × 10^-4^ to 0.021; Supplementary Table 18). In the held-out imaging sample, higher temporal BAG PRS was associated with lower odds of phlebitis and thrombophlebitis, including its lower-extremity subtype (two associations; Supplementary Table 18).

**Fig. 5:**
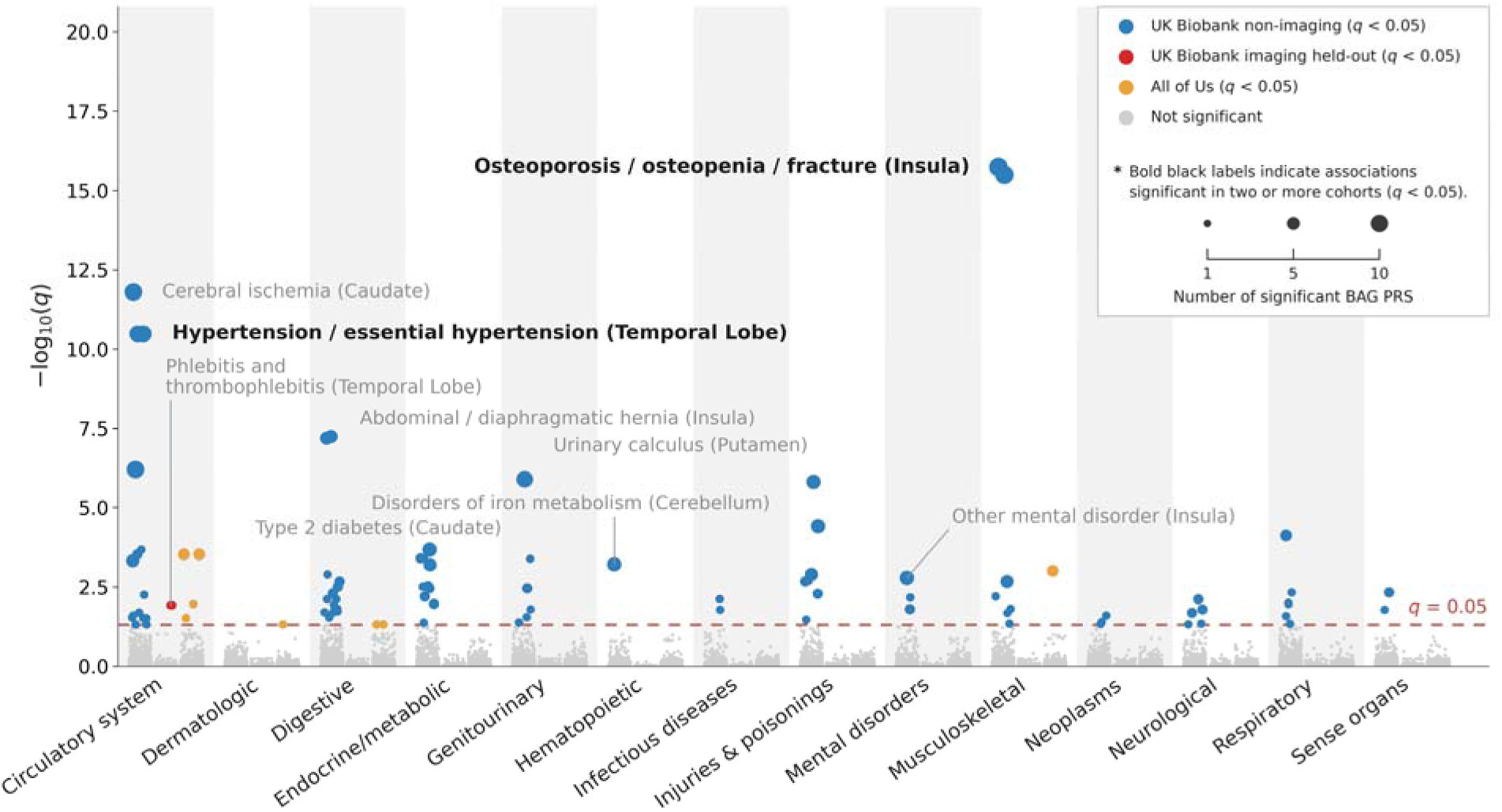
BAG PRS–PheWAS associations in relatedness-pruned target samples. Blue, red and orange points denote significant associations in White British UK Biobank non-imaging (*n* = 359,334), held-out imaging (*n* = 9,997) and European-ancestry All of Us sample (*n* ≤ 109,774), respectively. Highlighted points show the smallest q-value per phecode and sample; marker area represents the number of significant BAG PRS. Bold labels identify groups containing a phecode significant in both UK Biobank non-imaging and All of Us, not necessarily for the same PRS. Logistic models adjusted for age, sex and ten genetic principal components; sex-specific tests were restricted to the designated sex and excluded the sex covariate. Two-sided *P* values were Benjamini–Hochberg-corrected within each PRS and sample (*q* < 0.05). Parent and child phecodes were counted separately. All target samples were excluded from PRS derivation (Supplementary Note 11; Supplementary Tables 18–20).

In ADNI, PRS–BAG correlations were positive for all ten phenotypes but modest (*r* = 0.045–0.152; *n* = 1,693), with eight nominally significant associations (Supplementary Fig. 9a; Supplementary Table 21). No PRS association with AD or MCI status survived after FDR correction (*n* = 1,845; Supplementary Fig. 9b; Supplementary Tables 22 and 23).

### Bidirectional Mendelian randomization

Exploratory bidirectional Mendelian randomization (MR) identified eight inverse-variance-weighted (IVW) associations passing trait-and direction-specific Bonferroni correction and four additional FDR-only associations. IVW estimates linked doctor-diagnosed diabetes and higher body mass index (BMI) to higher whole-brain BAG, and hypertension to higher caudate BAG. With thalamic BAG as the exposure and hypertension as the outcome, the estimate was negative. These estimates were not consistently supported by sensitivity analyses (Supplementary Figs. 10 and 11; Supplementary Note 12; Supplementary Data 2).

## Discussion

Brain-age models trained separately on three-dimensional regional MRI inputs revealed substantial shared genetic influences alongside variation in clinical associations and cell-type annotations. Regional MAEs were lower than those reported in the regional studies summarized in Supplementary Table 1, although comparisons are descriptive because samples, regional definitions and evaluation designs differed. In ADNI models containing both temporal and whole-brain BAGs, temporal BAG was more strongly associated with CSF Aβ42 and cognition, whereas whole-brain BAG was more strongly associated with CSF p-tau181. Their associations with amyloid and tau-PET did not differ significantly. Regional models showed a modest advantage over whole-brain BAG for discriminating observed MCI-to-AD conversion, but combining regions did not clearly outperform temporal BAG alone. In UK Biobank, regional case–control differences were often significantly larger than whole-brain differences in years, but rarely so after scaling each BAG by its control-group standard deviation. Together, these findings support complementary clinical information from regional and whole-brain BAGs, with their relative strengths varying across clinical outcomes.

Jawinski et al.^26^ reported a genetic correlation of 0.74 between grey-and white-matter BAGs, indicating substantial but incomplete genetic overlap. Recent cortical local-age GWAS work distinguished broadly distributed from spatially restricted variant effects^28^. We extended the comparison to genome-wide genetic sharing among whole-brain, cortical, subcortical and cerebellar BAGs. Genetic correlations exceeded phenotypic correlations for every pair; the lower phenotypic correlations may reflect non-genetic variation and measurement error. The recurrent 17q21.31/*MAPT* signal coexisted with loci and mapped gene groups detected in one or a few BAGs, although detection in a single phenotype does not establish a region-specific effect.

Oligodendrocyte enrichment in whole-brain and cerebellar BAGs is consistent with white matter contributions to structural brain ageing^36,37^ and evidence linking oligodendrocyte dysfunction to age-related white matter pathology^38^. Other retained annotations included astrocytes and fibroblast/VLMCs for caudate BAG, OPCs for temporal BAG and fibroblast/VLMCs for thalamic BAG. Comparing BAGs derived from the same participants and imaging modality complements previous multimodal analyses^27^ (Fig. 4b; Supplementary Table 16). These are differences in observed enrichment profiles; enrichment differences between BAGs were not formally tested.

Genetic correlation estimates were negative with telomere length and mostly positive with ProtAge-204. In the relatedness-pruned UK Biobank and All of Us samples, regional BAG PRS shared positive associations with hypertension, essential hypertension and osteoporosis. However, no PRS association with AD or MCI diagnostic status in ADNI was significant after multiple test correction. Exploratory MR findings involving metabolic traits were not consistently supported by sensitivity analyses and do not establish causal effects.

Several limitations warrant consideration. Genetic analyses were restricted to self-reported White British UK Biobank participants or genetically inferred European-ancestry participants in ADNI and All of Us, leaving cross-ancestry generalizability untested. Differences in external GWAS sample sizes also limit comparisons of statistical significance.

Cross-sectional T1-weighted MRI and broad atlas masks cannot resolve within-person regional ageing trajectories, and disease-related changes such as multiple-sclerosis lesions may contribute to BAG. Protein sampling preceded MRI by a mean of 8.60 years, so phenotypic correlations between ProtAge-204 and BAGs do not compare contemporaneous ageing measures (Supplementary Note 6). ADNI was smaller and clinically enriched, and nine of ten BAGs were associated with MRI field strength, although temporal–whole-brain contrasts and diagnostic-stage associations were similar in 3 T-only analyses (Supplementary Tables 5 and 6). Restriction to quality-control-passed flortaucipir scans reduced the primary tau-PET sample, while differences between tracers limited interpretation of the pooled tau-PET analysis. The attenuated conversion-discrimination advantage under stricter follow-up requirements warrants independent longitudinal validation.

In summary, whole-brain and regional BAGs share substantial polygenic influences but show regional variation in their observed clinical associations and cell-type enrichment annotations.

## Methods

### Ethical approval

All research was conducted in accordance with relevant ethical regulations. This study used the UK Biobank Resource under Application 45227; UK Biobank received ethical approval from the North West Multi-centre Research Ethics Committee. ADNI was approved by the institutional review boards of all participating institutions, and the All of Us Research Program was approved by its institutional review board. All participants provided written informed consent.

### Statistics and reproducibility

Analyses included participants meeting the eligibility criteria for each cohort and analysis. Major cohort subsets are summarized in Supplementary Table 24; analysis-specific sample sizes are reported in the corresponding result tables and Supplementary Data 1.

Unless otherwise specified, tests were two-sided. Tests of nonzero SNP heritability and MAGMA tests for positive cell-type enrichment were one-sided. Multiple testing was controlled using the BH procedure at FDR < 0.05 within the families defined for each analysis. BH-adjusted *P* values are denoted by *q*. Genome-wide significance was set at *P* < 5 × 10^-8^, and cell-type enrichment used Bonferroni correction within each BAG phenotype. All confidence intervals were 95%. Analysis-specific statistical methods, validation procedures and software versions are provided in the corresponding Methods sections and Supplementary Notes.

### Participants

UK Biobank was the primary discovery cohort, ADNI provided an external AD-enriched cohort for imaging, clinical and genetic analyses, and All of Us provided an external PRS–PheWAS cohort. ADNI data were downloaded from the LONI Image and Data Archive on 8 April 2026 (genotypes and the diagnostic summary, DXSUM), 14 April–12 May 2026 (T1-weighted MRI), 20 May 2026 (ADNIMERGE2 v0.1.1 and REGISTRY) and 3 June 2026 (PTDEMOG).

Brain-age modelling included 46,254 UK Biobank participants: 25,642 for training and 20,612 for independent testing. BAG GWAS and post-GWAS analyses included 41,067 imaging participants with self-reported White British ethnicity after genotype quality control. ADNI included 2,986 imaging participants: 1,179 cognitively normal participants for fine-tuning and 1,807 for inference (1,303 with MCI, 420 with AD and 84 with significant memory concern).

Genetic analyses in ADNI and All of Us were restricted to genetically inferred European ancestry. All of Us relatedness pruning retained 112,080 candidates; excluding 2,306 participants with missing sex left 109,774 with complete covariates. Related-disease exclusions and sex restrictions further reduced phenotype-specific sample sizes (Supplementary Table 24).

### Image preprocessing

UK Biobank provided skull-stripped T1-weighted images nonlinearly registered to Montreal Neurological Institute 152 (MNI152) space^39^. ADNI scans underwent N4 bias-field correction, brain extraction and nonlinear registration to MNI152 using the Advanced Normalization Tools Python interface (ANTsPy v0.5.4)^40^. Images from both cohorts were resampled to a common 128 × 128 × 128 grid. After resampling and before regional masking, ADNI image intensities were normalized by scaling the 99th percentile of positive voxel values to 105, without upper clipping. The MNI structural atlas^41,42^ defined nine regional inputs: frontal, parietal, temporal and occipital lobes; insula; putamen; caudate; thalamus; and cerebellum.

### Region-□ise brain-age prediction and BAG derivation

Separate 3D-CNNs predicted chronological age from whole-brain and each regional MRI input using a previously described architecture^20^ (Supplementary Fig. 1). Regional atlas labels were binarized, dilated by four voxels on the atlas grid and resampled to the image grid by nearest-neighbour interpolation. The same mask was applied to every participant, setting voxels outside the region to zero.

Models were trained in 25,642 UK Biobank participants selected using previously reported disease-exclusion criteria^20^. Within each fold, parameters were optimized on the other three folds, and the checkpoint with the lowest held-out-fold MAE was retained. The independent test set (*n* = 20,612) was used for neither optimization nor checkpoint selection; test predictions were averaged across the four models. In both cohorts, accuracy was assessed using uncorrected predicted ages (MAE and Pearson *r*; Supplementary Table 1). Diagnosis-specific correlation intervals used the Fisher z transformation, treating fitted predictions as fixed (Supplementary Table 2). Exclusion criteria and training settings are given in Supplementary Note 4.

Age-bias correction coefficients were estimated from pooled fold-held-out training predictions and applied unchanged to all UK Biobank participants^43^. BAG was corrected predicted age minus chronological age, with positive values indicating an older-appearing brain (Supplementary Note 5; Supplementary Fig. 2).

### Disease-associated regional BAG profiles across the UK Biobank and ADNI cohorts

UK Biobank hospital inpatient ICD-10 records were mapped to phecodes v1.2. Cases required at least one qualifying record; 24 phenotypes had at least 30 cases in the independent test set. Self-reported illness codes were not used. Records before and after MRI were included, so these were cross-sectional associations with recorded diagnoses (Supplementary Note 1).

Cases were compared with 10,776 test-set participants without selected diagnoses. Control-selection criteria differed from the training-exclusion criteria (Supplementary Notes 1 and 4). For each diagnosis, we fitted a linear regression of BAG on case status, adjusting for age and sex. The case coefficient represented the adjusted case–control difference in BAG years. Breast-and prostate-cancer comparisons were sex-restricted and did not include sex as a covariate. Regional and whole-brain case coefficients were compared in the same participants, in years and after scaling each BAG by its control-group standard deviation. These contrasts used standard errors clustered by participant. An expanded-control sensitivity analysis added fold-held-out predictions from 17,910 training participants without selected diagnoses, giving 28,686 controls. The same sex restrictions applied throughout (Supplementary Note 1; Supplementary Table 3, continued).

ADNI analyses compared MCI (*n* = 1,232) and AD (*n* = 398) separately with cognitively normal participants (*n* = 1,122), adjusting for age at MRI, sex and field strength (1.5 T or 3 T). We excluded 149 participants with missing MRI field strength and one additional participant whose sex was not coded as male or female. Two-sided Wald tests and 95% confidence intervals were calculated using heteroskedasticity-robust standard errors (HC3). BH correction was applied across ten BAGs within each diagnosis, including the expanded-control analysis. Regional–whole-brain contrasts were corrected across nine regions within each diagnosis, separately for each scale.

### Domain-adapted brain-age estimation in the ADNI cohort

ADNI models used the same masks and architecture, initialized from the UK Biobank checkpoint for the corresponding target and fold. All layers were fine-tuned in 1,179 cognitively normal participants using shuffled four-fold cross-validation with a fixed seed. As in UK Biobank, checkpoints were selected by held-out-fold MAE (Supplementary Note 4). Fold-held-out predictions in cognitively normal participants were used to assess accuracy and estimate ADNI-specific bias correction. Predictions for other participants were averaged across the four models. Correction coefficients estimated in cognitively normal participants were applied unchanged to all participants before BAG calculation.

### AD-related biomarkers and cognitive and diagnostic associations in the ADNI cohort

AD-related measures were obtained from ADNIMERGE2. Outcomes comprised amyloid PET (Centiloids), tau-PET (temporal meta-region standardized uptake value ratio [SUVR] referenced to inferior cerebellar grey matter), Aβ42 and p-tau181, measured using Roche Elecsys, and Clinical Dementia Rating–Sum of Boxes (CDR-SB), the 13-item Alzheimer’s Disease Assessment Scale–Cognitive Subscale (ADAS-Cog13), and Mini-Mental State Examination (MMSE) scores. Each participant contributed one baseline MRI, with PET and CSF measurements matched within 365 days and cognitive assessments within 180 days.

Each BAG–outcome association was tested by complete-case linear regression adjusted for age, sex and *APOE* ε4 allele count. BAGs were standardized using the mean and standard deviation of the full ADNI baseline sample (*n* = 2,986), so coefficients represent native outcome units per 1-s.d. higher BAG. BH correction covered all 70 associations.

To compare regional with whole-brain associations, each model included one regional BAG and whole-brain BAG with the same covariates. Their coefficients were compared by a linear contrast. Regional-coefficient and coefficient-contrast *P* values were BH-corrected separately across 63 tests each (nine regions × seven outcomes).

Baseline diagnostic stage combined CN and significant memory concern (SMC) as stage 0; early MCI, MCI and late MCI as stage 1; and AD as stage 2. BAG–stage regressions adjusted for age, sex and *APOE* ε4 allele count, with BH correction across ten BAGs. These coefficients describe cross-sectional stage associations, not within-person progression.

Primary tau-PET analyses were restricted to 6-mm flortaucipir scans that passed the provider’s quality control (*n* = 684). A sensitivity analysis pooled tau-PET scans acquired with flortaucipir, MK6240 and PI2620. For amyloid PET, sensitivity analyses were restricted to scans passing the provider’s quality control. Separately, models containing regional and whole-brain BAGs and diagnostic-stage analyses were repeated in participants scanned at 3 T. Additional methods are provided in Supplementary Note 8, and results in Supplementary Tables 4–6 and Supplementary Data 1.

### Discrimination of observed MCI-to-AD conversion

Baseline was defined as each participant’s first MRI visit matched to a diagnosis in the ADNI diagnostic summary (DXSUM). Eligibility required MCI at that visit. Conversion required a subsequent AD diagnosis at a visit with MRI. After exclusions, the analysis included 1,021 participants: 303 converters and 718 non-converters whose diagnosis remained MCI throughout the available diagnostic record. At least one post-baseline diagnostic visit was required, but no minimum follow-up duration or fixed conversion window was imposed in the primary analysis.

Logistic regression models included age and sex together with either whole-brain BAG or selected regional BAGs. Performance was evaluated using stratified ten-fold outer cross-validation. Within each outer-training fold, inner five-fold cross-validation selected the number of regional predictors, k = 1–9, with regions ranked using training participants only. Ranking used the univariate AUC of regional BAG residuals obtained using whole-brain adjustment coefficients estimated in cognitively normal participants; the prediction models used the bias-corrected regional BAGs rather than these residuals. Predictors were standardized using training data only. Fixed-k models were evaluated descriptively.

AUCs were calculated from pooled outer-fold predictions. Conditional on precomputed BAGs, 1,000 participant-level bootstrap replicates repeated ranking, k selection, standardization, fitting and held-out evaluation to obtain percentile confidence intervals for the regional–whole-brain AUC difference. Copies of each resampled participant remained in the same fold at every cross-validation level. Sensitivity analyses used all nine regional BAGs in a ridge model, with penalty selection by inner three-fold cross-validation, or required at least 24 months of follow-up for non-converters (Supplementary Note 9; Supplementary Data 1).

### Genome-wide association and functional annotation analyses

BAG GWASs included 41,067 White British UK Biobank participants and used SAIGE v1.1.8 for null-model fitting and v1.1.9 for association testing^44,45^. Each BAG was rank-based inverse-normal transformed across the full sample before additive association testing adjusted for age, sex and ten genetic principal components. A dense genetic relationship matrix modelled relatedness with leave-one-chromosome-out analysis. UK Biobank imputation v3 BGEN files on GRCh37^46^ yielded 35,784,891 tested variants per phenotype after requiring minor allele count ≥10 and missingness ≤15%. Locus definition and annotation retained 9,458,475 variants with analysis-sample minor allele frequency (MAF) ≥0.01 and mfi imputation quality (INFO) ≥0.8 (Supplementary Note 10; Supplementary Table 25).

FUMA v1.8.2^47^ used the 1000 Genomes Phase 3 European reference to identify independent significant variants (P < 5 × 10^-8^; linkage disequilibrium *r*^2^ < 0.6) and lead variants (*r*^2^ < 0.1). Independent signals within 250 kb were merged into genomic risk loci. The MHC interval (GRCh37 chr6:29,614,758–33,170,276) was excluded from FUMA annotation, including lead-variant and locus definition. This exclusion did not apply to GWAS association testing or MAGMA gene analysis.

Mapped gene groups were defined by distinct GWAS Catalog MappedGene annotations rather than genomic intervals. Lead variants were pooled across the ten BAG GWASs. A group was detected in a BAG phenotype when at least one variant from this pooled set carried the group annotation in that phenotype’s FUMA candidate-variant results (*P* < 0.05 and *r*^2^ ≥ 0.6 with an independent significant variant). Groups detected in one phenotype were classified as unique, and those detected in two or more as shared (Fig. 3b). Genotype processing and annotation details are provided in Supplementary Note 10, and trait-category curation in Supplementary Note 3.

Exploratory GWASs were also performed in 1,693 ADNI participants of genetically inferred European ancestry. UK Biobank lead variants selected from the common biallelic variant set were evaluated for directional concordance in ADNI after allele harmonization. Detailed methods and results are provided in Supplementary Note 2, Supplementary Tables 7–10 and Supplementary Figs. 5 and 6.

### SNP-based heritability and correlations among BAGs

SNP-based heritability and pairwise genetic correlations were estimated using LD Score regression (LDSC v1.0.1; commit aa33296)^48,49^ and 1000 Genomes European-ancestry LD scores. Heritability refers to inverse-normal-transformed BAGs (Supplementary Table 14; Supplementary Note 6). Regional–whole-brain heritability differences used the same 41,067 participants and regression SNPs. Standard errors were estimated by a paired jackknife over 200 shared genomic blocks, accounting for covariance between estimates. Two-sided normal-theory P values were BH-corrected across nine comparisons (Supplementary Table 14, continued).

Genetic correlations were estimated for all 45 BAG pairs with unconstrained cross-trait intercepts to account for sample overlap and correlated errors. Phenotypic correlations were unadjusted Pearson correlations between inverse-normal-transformed BAGs in the same 41,067 participants. BH correction was applied separately to the 45 genetic and 45 phenotypic correlations (Supplementary Table 15).

### Genetic and phenotypic correlations with external ageing measures

Leukocyte telomere length PC1 GWAS summary statistics^34^ were obtained from the GWAS Catalog (GCST90435144; *n* = 438,351). PC1 represents the shared component with higher values indicating longer telomeres.

ProtAge-204 was trained locally using the fixed 204-protein panel of Argentieri et al.^35^, with protein quality control and k-nearest-neighbour imputation described in Supplementary Note 6. The proteomic-age gap from out-of-fold predictions was age-bias corrected and inverse-normal transformed before GWAS in 38,806 UK Biobank participants with self-reported British ethnicity and genotype data.

Genetic correlations were estimated using cross-trait LDSC with unconstrained intercepts. BH correction was applied jointly across 21 correlations: ten BAGs with each external ageing measure and the correlation between the two ageing measures (Supplementary Table 15).

Phenotypic correlations between ProtAge-204 and BAGs used the bias-corrected gaps before inverse-normal transformation in 3,784 participants. Partial correlations adjusted for sex, age at protein sampling and the protein–MRI interval, with BH correction across ten BAGs (Supplementary Note 6; Supplementary Table 15).

### Cell-type enrichment analysis

The FUMA cell-type module (FUMA v1.8.2; MAGMA v1.10)^50^ tested whether genes with greater cell-type expression specificity showed stronger gene-level BAG associations. Bonferroni correction was applied within each BAG across dataset–cell-type tests.

Significant annotations were retained when they were the only significant annotation in their dataset or met FUMA’s within-dataset conditional criteria. Cross-dataset pairwise conditional analyses assessed dependence but did not change retention. Source annotations were harmonized into 15 cell classes (Supplementary Note 7; Supplementary Table 16).

### Polygenic risk scores and cross-cohort phenome-wide association analyses

PRS for the ten BAGs were derived using SBayesRC (GCTB v2.5.2)^51,52^, with the UK Biobank European HapMap3 LD reference and baseline-LD v2.2 annotations^53^. SBayesRC was configured with the default five-component SNP-effect mixture prior, with relative variance multipliers of 0, 0.001, 0.01, 0.1 and 1 and annotation-dependent mixture probabilities. Each deposited BAG weight set was generated using a single MCMC chain with 3,000 iterations, 1,000 burn-in iterations, a thinning interval of 10, random seed 20260821 and four threads. PLINK 2.0 was used for quality control and scoring^54,55^. GWASs were repeated with the same model specification in 30,800 White British imaging participants, and variant weights were held fixed in all target samples.

In the relatedness-pruned non-overlapping held-out sample (n = 9,997), each inverse-normal-transformed BAG was regressed on its corresponding PRS, standardized using that sample’s mean and standard deviation, in a univariable linear model. We reported adjusted R² (Supplementary Table 17; Supplementary Fig. 8).

PheWAS used relatedness-pruned White British UK Biobank non-imaging (*n* = 359,334) and held-out imaging (*n* = 9,997) samples, and European-ancestry All of Us participants with complete covariates (*n* = 109,774). UK Biobank non-imaging participants were outside the 46,254-participant imaging cohort. In both UK Biobank targets, participants related to any PRS-derivation participant at KING kinship > 0.0884 were excluded, followed by greedy within-target pruning at the same threshold. All of Us pruning used the All of Us v7 precomputed relatedness exclusion list; no pair in the supplied kinship file remained among retained candidates (Supplementary Note 11; Supplementary Table 24).

ICD-10 records were mapped to phecodes v1.2 and aggregated to the first decimal place. Cases required at least one qualifying record, and related-disease control exclusions were applied in all three samples. Each PRS–phecode association was tested by logistic regression adjusted for age, sex and ten genetic principal components. UK Biobank PheWAS age was 2018 minus birth year; All of Us age was elapsed days from birth to 1 January 2023 divided by 365. Sex-specific phecodes were restricted to the designated sex and did not include sex as a covariate; ambiguous phecode 751.1 remained unrestricted.

Models used complete cases and required at least 30 cases after exclusions and sex restrictions. Unsuccessful fits were excluded. Coefficients and odds ratios are per one-unit increase in the unstandardized summed PRS. BH correction was applied within each PRS and sample (Supplementary Note 11; Supplementary Tables 18–20).

In ADNI, Pearson correlations related each PRS to its corresponding bias-corrected baseline BAG in 1,693 participants (Supplementary Table 21). Clinical-label analyses included 1,845 of 1,856 genotype-QC-passed participants, excluding 11 with missing age or baseline diagnosis. Logistic models compared AD and MCI separately with cognitively normal status, adjusting for age, sex, ten genetic principal components and *APOE* ε4 allele count. Odds ratios are per one-unit increase in the unstandardized summed PRS. Sensitivity analyses excluded *APOE* ε4 allele count from the covariates. BH correction was applied separately across the 20 tests in each analysis (Supplementary Tables 22 and 23).

### Mendelian randomization analysis

Bidirectional two-sample MR examined disease-related traits selected from the UK Biobank BAG profiles (Supplementary Note 1). Disease-related traits were exposures and BAGs were outcomes in forward analyses; reverse analyses exchanged these roles. Instruments met exposure-GWAS *P* < 5 × 10^-8^ and were LD-clumped at *r*^2^ < 0.001 within 10 Mb. IVW was the primary estimator.

Forward coefficients are in inverse-normal-transformed BAG units per exposure-GWAS unit; reverse coefficients are in outcome-GWAS units per transformed-BAG unit, not BAG years. Doctor-diagnosed diabetes used the source GWAS linear-probability scale and BMI used standard-deviation units. BH correction and a Bonferroni threshold of *P* < 0.005 were applied across ten BAGs within each trait and direction. These analyses were exploratory because of data-informed trait selection, sample overlap and limited reverse-direction instruments (Supplementary Note 12; Supplementary Data 2; Supplementary Figs. 10 and 11).

## Data availability

UK Biobank GWAS summary statistics for the whole-brain and nine regional BAGs and the ProtAge-204 proteomic-age gap, together with the ten SBayesRC BAG PRS weight sets, are publicly available through Google Cloud Storage at https://storage.googleapis.com/leelabsg-regional-brain-age/README.md. The BAG summary-statistic files contain the 9,458,475 variants passing the post-GWAS quality-control criteria.

Individual-level data cannot be redistributed by the authors and are available to approved researchers through UK Biobank (Application 45227; https://www.ukbiobank.ac.uk/enable-your-research/apply-for-access), ADNI (https://adni.loni.usc.edu/data-samples/access-data/) and the All of Us Researcher Workbench (Controlled Tier Curated Data Repository v7; https://www.researchallofus.org/data-tools/workbench/).

Leukocyte telomere length PC1 GWAS summary statistics are available from the GWAS Catalog (GCST90435144). Sources and accessions for MR summary statistics are listed in Supplementary Data 2. Supporting statistics are provided in the Supplementary Tables, Supplementary Data files and Supplementary Figures.

## Code availability

Custom code for the region-wise brain-age models, the UK Biobank and ADNI GWAS, LD Score regression, MAGMA cell-type enrichment, SBayesRC polygenic score derivation and scoring, the UK Biobank phenome-wide association analysis, and the ADNI PRS validation is available at https://github.com/leelabsg/Region-Wise-Brain-Age-Estimation

The All of Us Researcher Workbench code is not included in the public repository. Running the pipeline requires approved access to individual-level cohort data.

## Supporting information

Supplementary Data 1: ADNI clinical and conversion analyses

Supplementary Data 2: Mendelian randomization results

## Acknowledgements

This work was supported by the Brain Pool Plus (BP+) Program of the National Research Foundation of Korea (NRF), funded by the Ministry of Science and ICT, Republic of Korea (2020H1D3A2A03100666), and by the Korea Health Technology R&D Project through the Korea Health Industry Development Institute (KHIDI), funded by the Ministry of Health and Welfare, Republic of Korea (RS-2025-02307233).

This research was conducted using the UK Biobank Resource under Application Number 45227. This work uses data provided by patients and collected by the NHS as part of their care and support.

We gratefully acknowledge All of Us participants for their contributions, without whom this research would not have been possible. We also thank the National Institutes of Health’s All of Us Research Program for making the participant data examined in this study available.

Data collection and sharing for the Alzheimer’s Disease Neuroimaging Initiative (ADNI) is funded by the National Institute on Aging (National Institutes of Health Grant U19 AG024904). The grantee organization is the Northern California Institute for Research and Education. In the past, ADNI has also received funding from the National Institute of Biomedical Imaging and Bioengineering, the Canadian Institutes of Health Research, and private sector contributions through the Foundation for the National Institutes of Health (FNIH) including generous contributions from the following: AbbVie, Alzheimer’s Association; Alzheimer’s Drug Discovery Foundation; Araclon Biotech; BioClinica, Inc.; Biogen; Bristol-Myers Squibb Company; CereSpir, Inc.; Cogstate; Eisai Inc.; Elan Pharmaceuticals, Inc.; Eli Lilly and Company; EuroImmun; F. Hoffmann-La Roche Ltd and its affiliated company Genentech, Inc.; Fujirebio; GE Healthcare; IXICO Ltd.; Janssen Alzheimer Immunotherapy Research & Development, LLC.; Johnson & Johnson Pharmaceutical Research & Development LLC.; Lumosity; Lundbeck; Merck & Co., Inc.; Meso Scale Diagnostics, LLC.; NeuroRx Research; Neurotrack Technologies; Novartis Pharmaceuticals Corporation; Pfizer Inc.; Piramal Imaging; Servier; Takeda Pharmaceutical Company; and Transition Therapeutics.

## Author contributions

Y.H. designed this study, developed the methodology, performed all the analyses and wrote the manuscript. K.-I.P. provided clinical expertise and contributed to the interpretation of the results. S.L. conceived this study, acquired funding, supervised the research and revised the manuscript.

All the authors read and approved the final manuscript.

## Competing interests

The authors declare that they have no competing interests.

## Notes

### Competing Interest Statement

The authors have declared no competing interest.

### Author Declarations

This study used the UK Biobank Resource under Application 45227. UK Biobank received ethical approval from the North West Multi-centre Research Ethics Committee. The Alzheimer's Disease Neuroimaging Initiative (ADNI) was approved by the institutional review boards of all participating institutions, and the All of Us Research Program was approved by its institutional review board. All participants provided written informed consent.

