## Supplementary Information for "Regional brain-age gaps reveal a shared polygenic architecture and regionally patterned clinical and cell-type associations"

**Contents**

Supplementary Figures 1–11

Supplementary Notes 1–12

References for the Supplementary Notes

**Separate supplementary files**

Supplementary Tables 1–25 are supplied in an Excel workbook.

Supplementary Data 1 contains ADNI clinical and conversion analyses.

Supplementary Data 2 contains Mendelian randomization results and supporting metadata.

### Supplementary Figures

##### Supplementary Fig. 1 | Architecture of the region-wise three-dimensional convolutional neural network.

**a,** Overall architecture. Each model takes a single-channel 128 × 128 × 128 volume in MNI152 space as input and outputs a scalar age prediction. Seven three-dimensional convolutional blocks are followed by flattening and three fully connected layers; feature-map dimensions are shown below each block as channels × spatial extent. **b,** Composition of the convolutional blocks. Blocks 1 and 2 use a shallow template comprising two convolutions, with three-dimensional dropout applied after the second convolution. Blocks 4–7 use a deep template comprising three convolutions, each followed by batch normalization. Block 3 follows the shallow template but replaces dropout after the second convolution with batch normalization. Block 7 follows the deep template but omits the final max-pooling operation, thereby preserving a 2 × 2 × 2 spatial extent. **c,** Stage-wise summary of layer composition, output channels, output shape and parameter count. All convolutions use 3 × 3 × 3 kernels with stride 1 and ‘same’ padding; max-pooling uses a 2 × 2 × 2 window with stride 2; dropout (*p* = 0.2) is applied only in blocks 1 and 2. Parameter counts include convolutional and fully connected weights and biases, and batch-normalization affine parameters. Each model has 2,984,801 trainable parameters. The same architecture was used for the whole-brain model and each of the nine regional models, which were trained independently; training settings are provided in Supplementary Note 4.

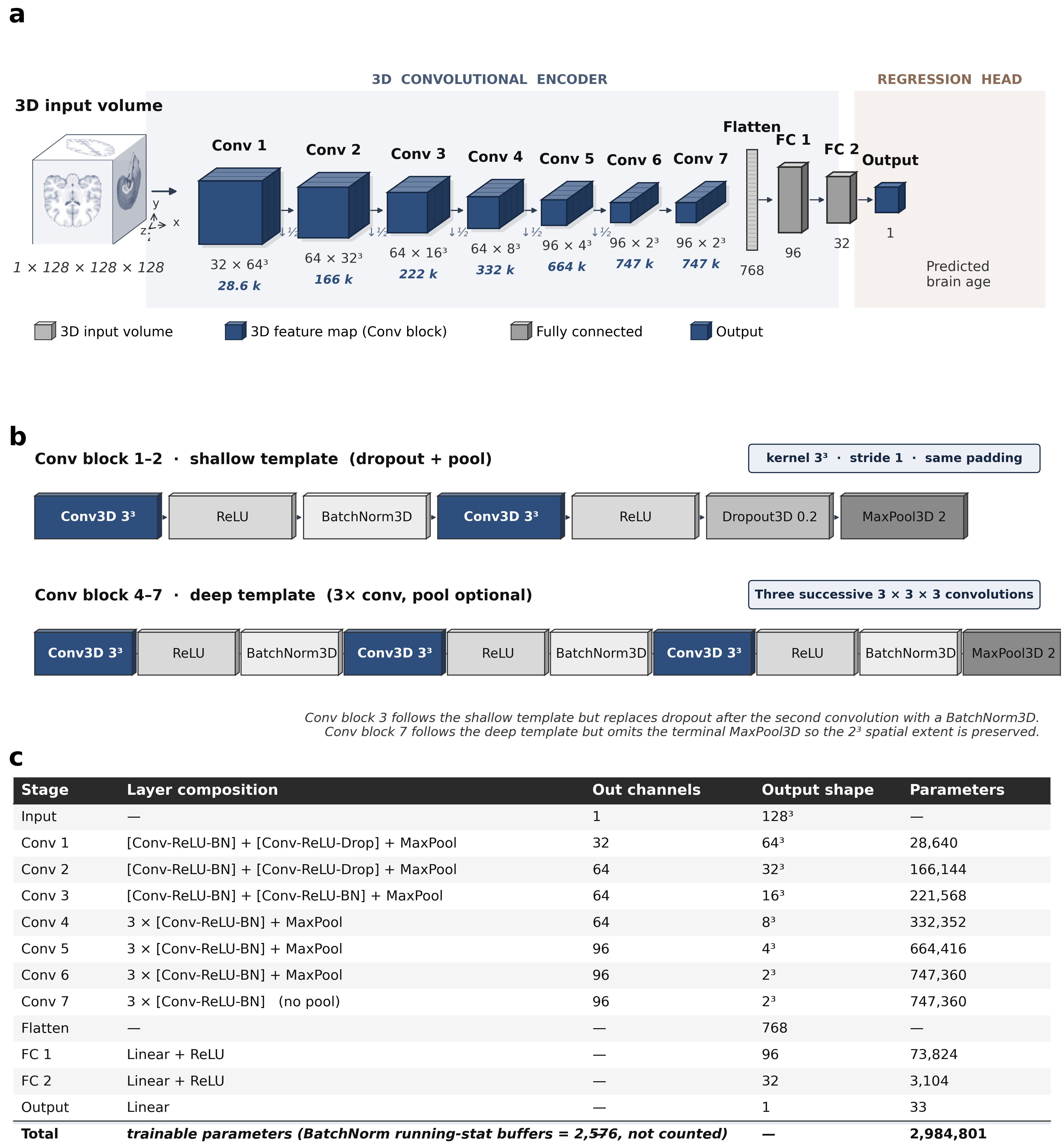

##### Supplementary Fig. 2 | Region-wise calibration and age-dependent bias correction of brain-age estimates.

For each model, panels show chronological age versus (i) uncorrected predicted age, (ii) uncorrected brain-age gap (BAG; predicted age minus chronological age) and (iii) bias-corrected BAG. Blue and orange points represent training participants (n = 25,642) and independent test participants (n = 20,612), respectively. Training predictions were generated out of fold, whereas test predictions were averaged across the four fitted models. Correction parameters were estimated from pooled out-of-fold training predictions and applied unchanged to both sets (Supplementary Note 5). Solid lines show ordinary least-squares fits within each set, with slopes annotated in each panel; dashed horizontal lines indicate zero BAG. Training-set slopes in column (iii) are zero by construction, whereas test-set slopes quantify residual linear age dependence after applying the training-derived correction.

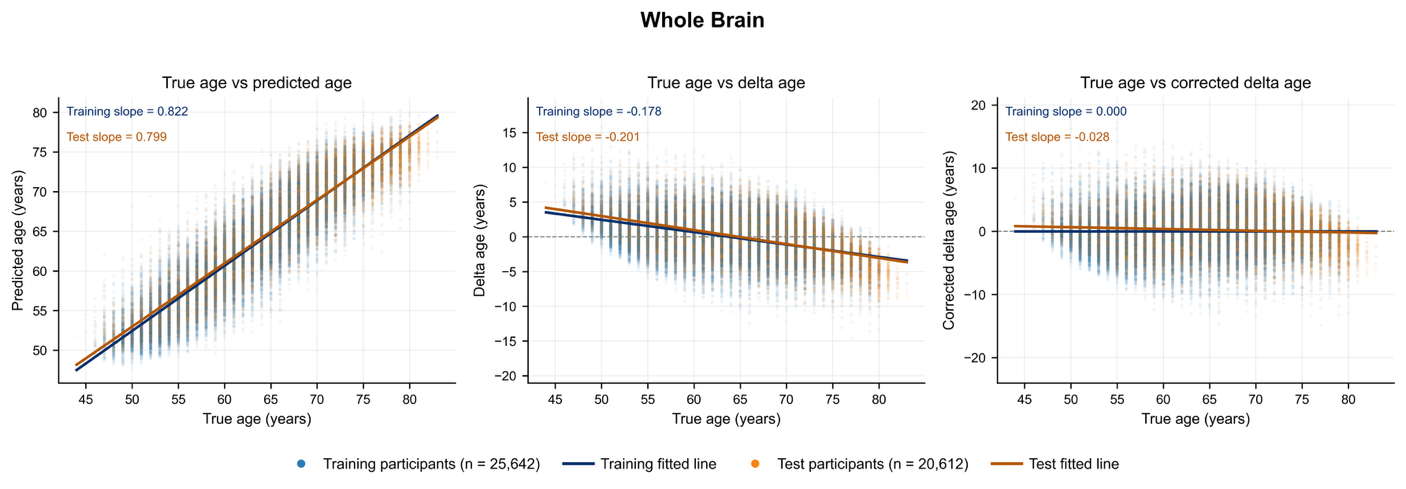

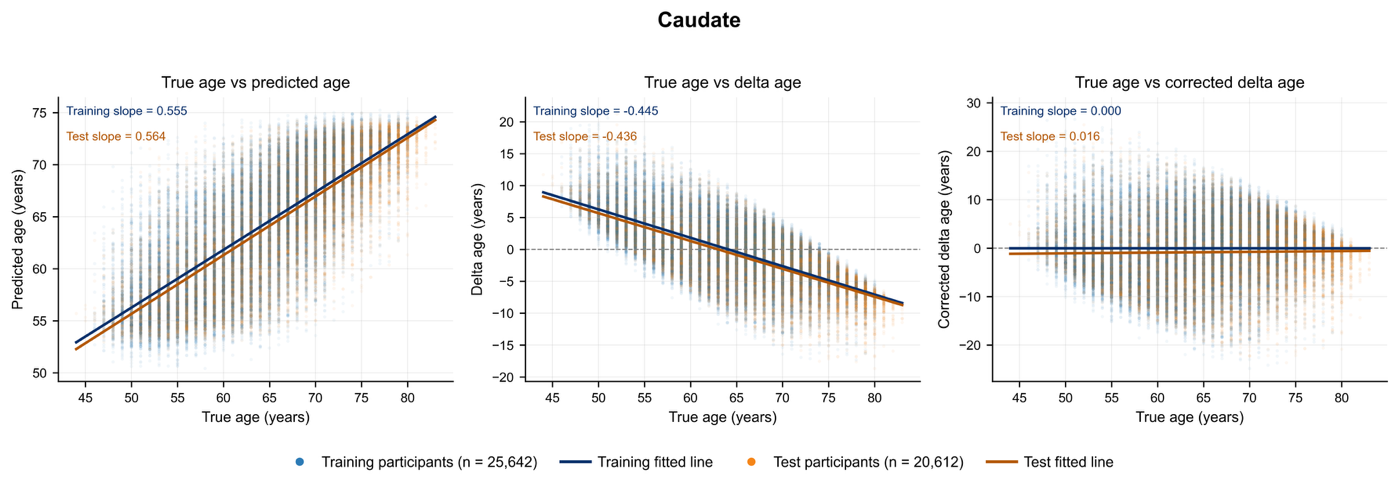

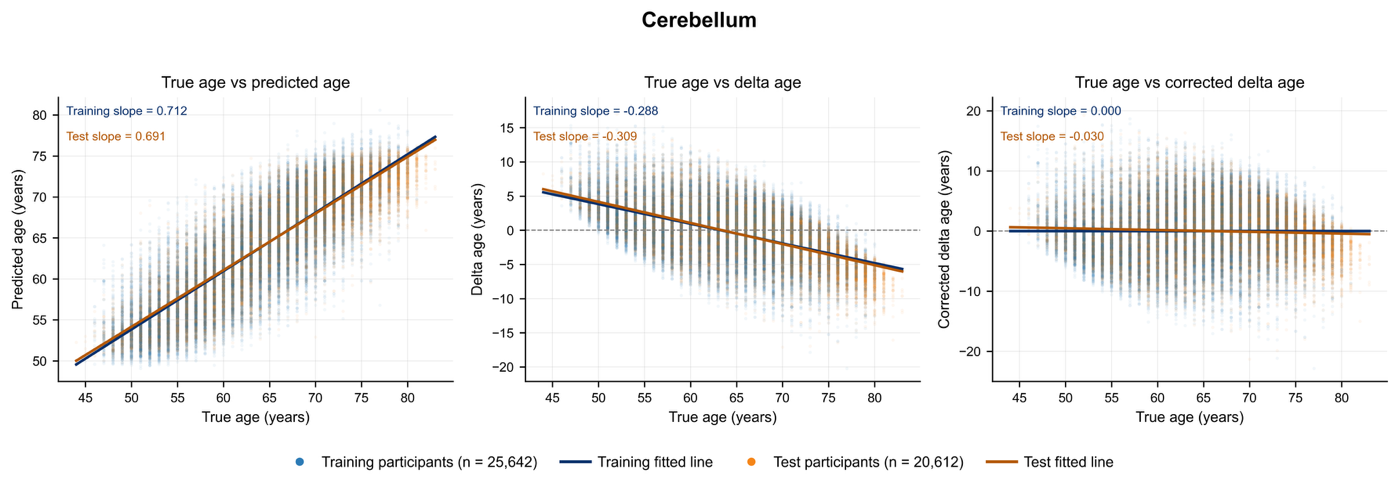

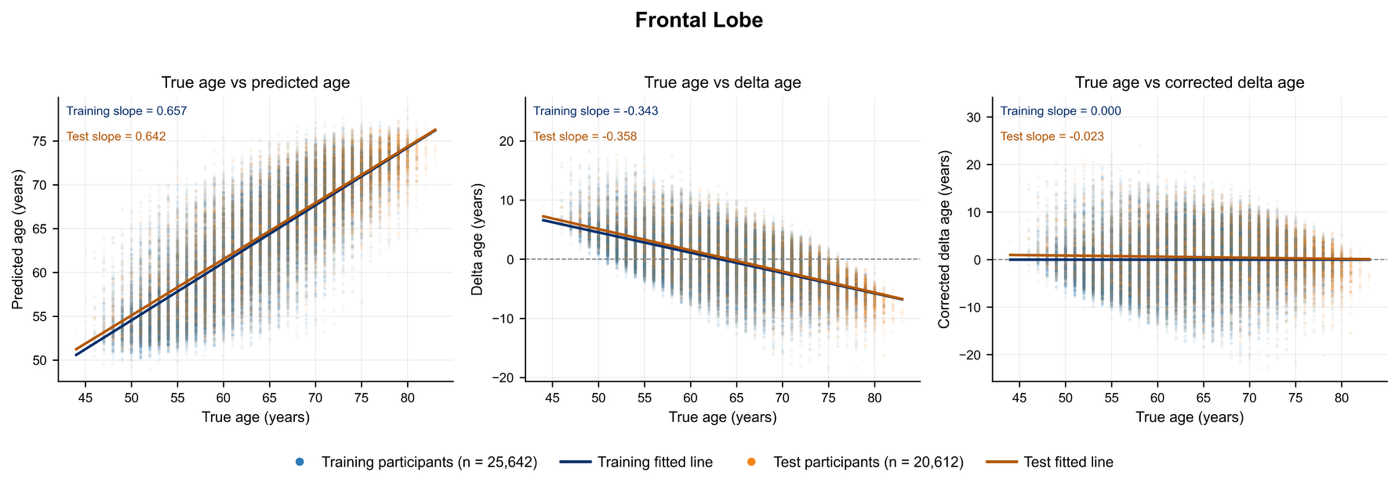

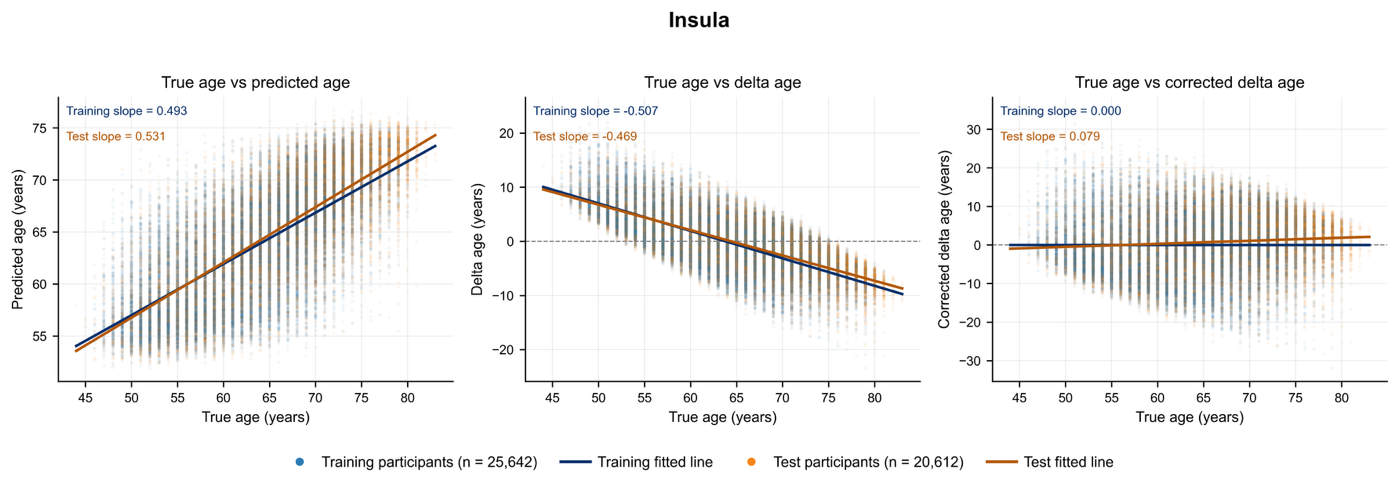

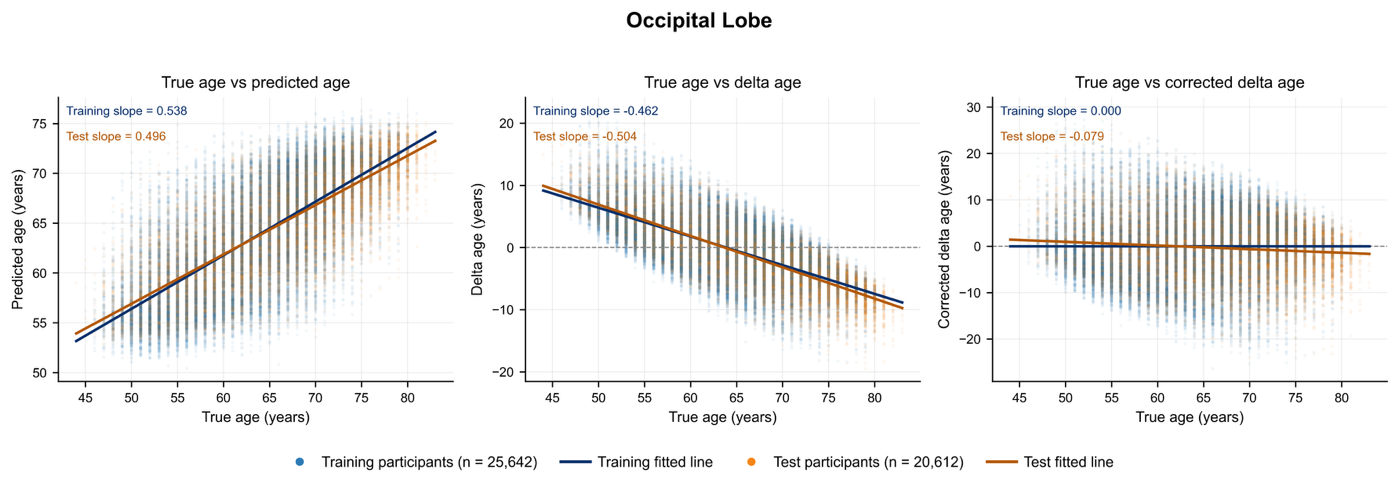

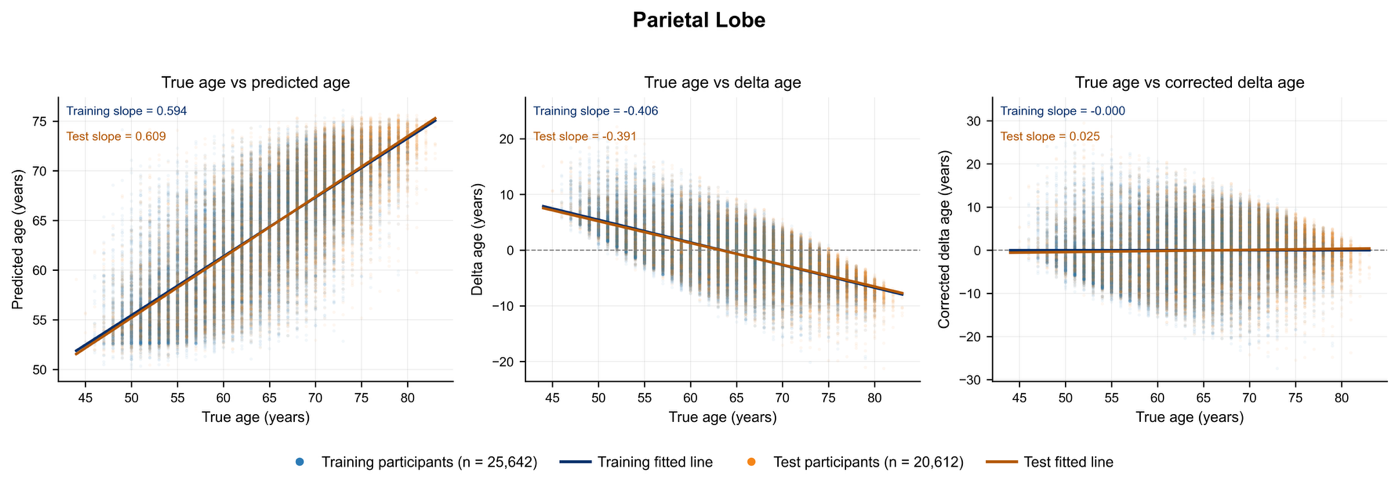

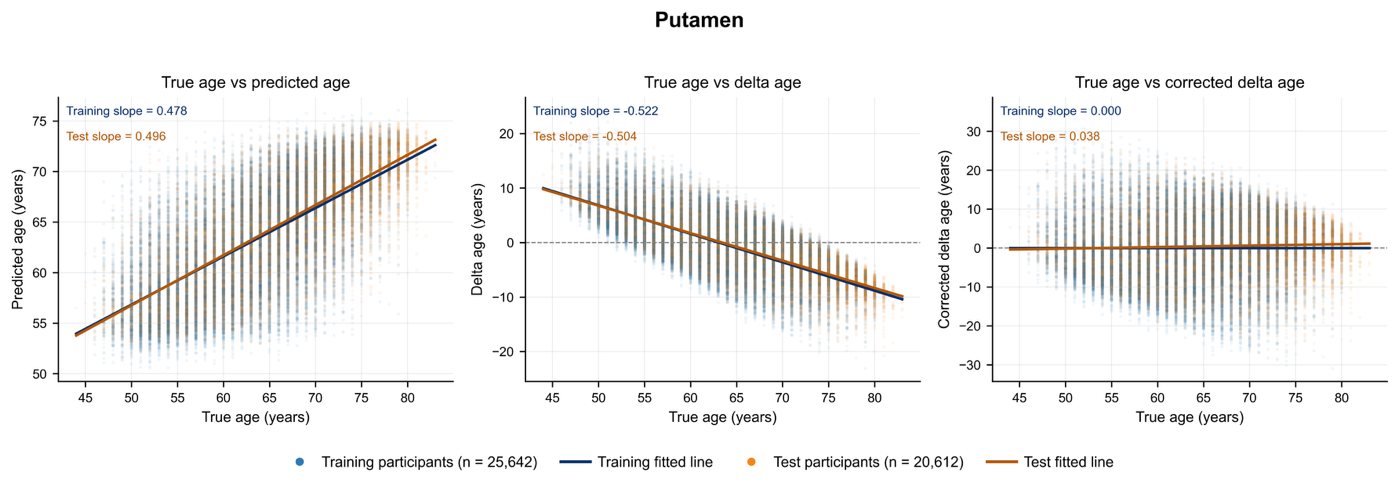

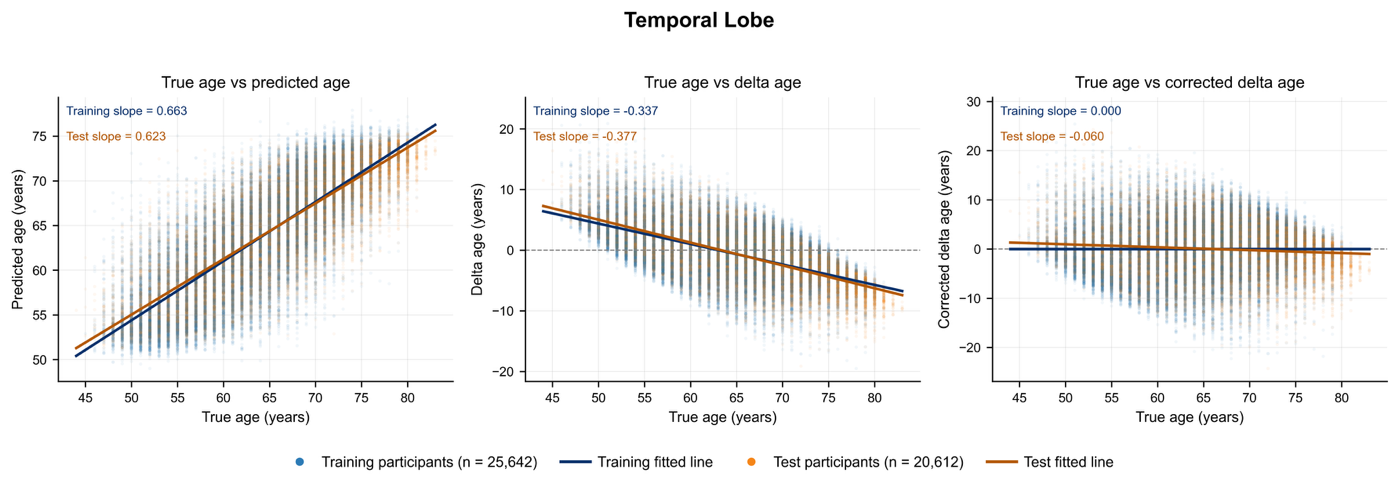

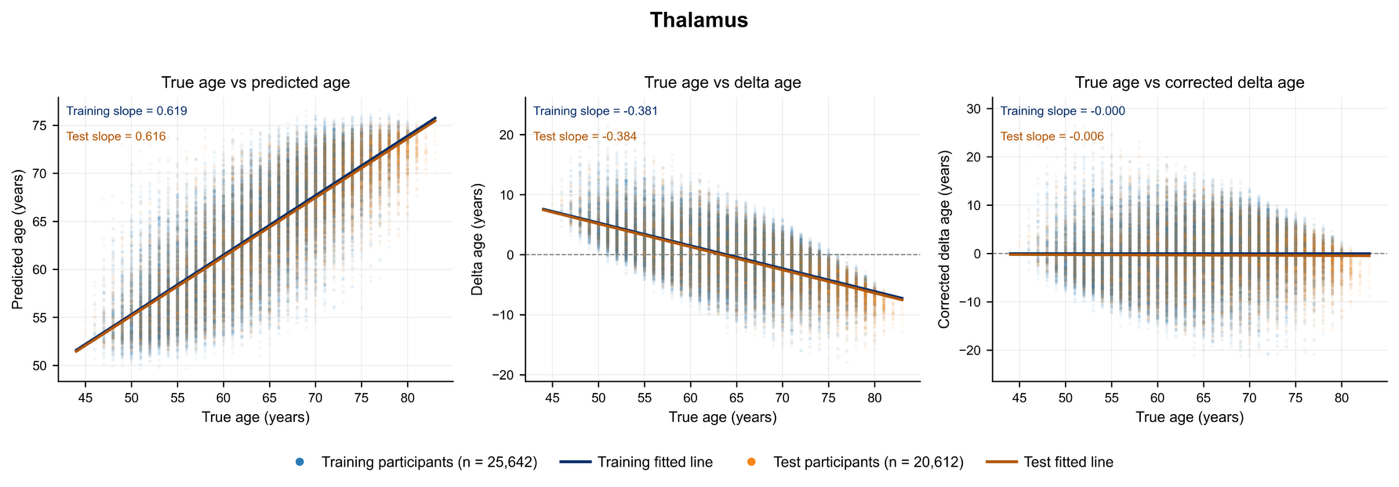

##### Supplementary Fig. 3 | Distributions of regional brain-age gap in representative UK Biobank diagnoses.

Split density plots show bias-corrected BAG distributions across the whole brain and nine regions for seven representative UK Biobank diagnoses. Upper grey distributions represent test-set controls without selected diagnoses (*n* = 10,776), and lower coloured distributions represent cases; case counts are shown above each panel. White lines indicate group medians. Distributions show individual BAG values, whereas colour intensity represents the age- and sex-adjusted case–control difference reported in Supplementary Table 3. The cognitive-disorder/cerebral-degeneration panel combines phecodes 290 and 331.

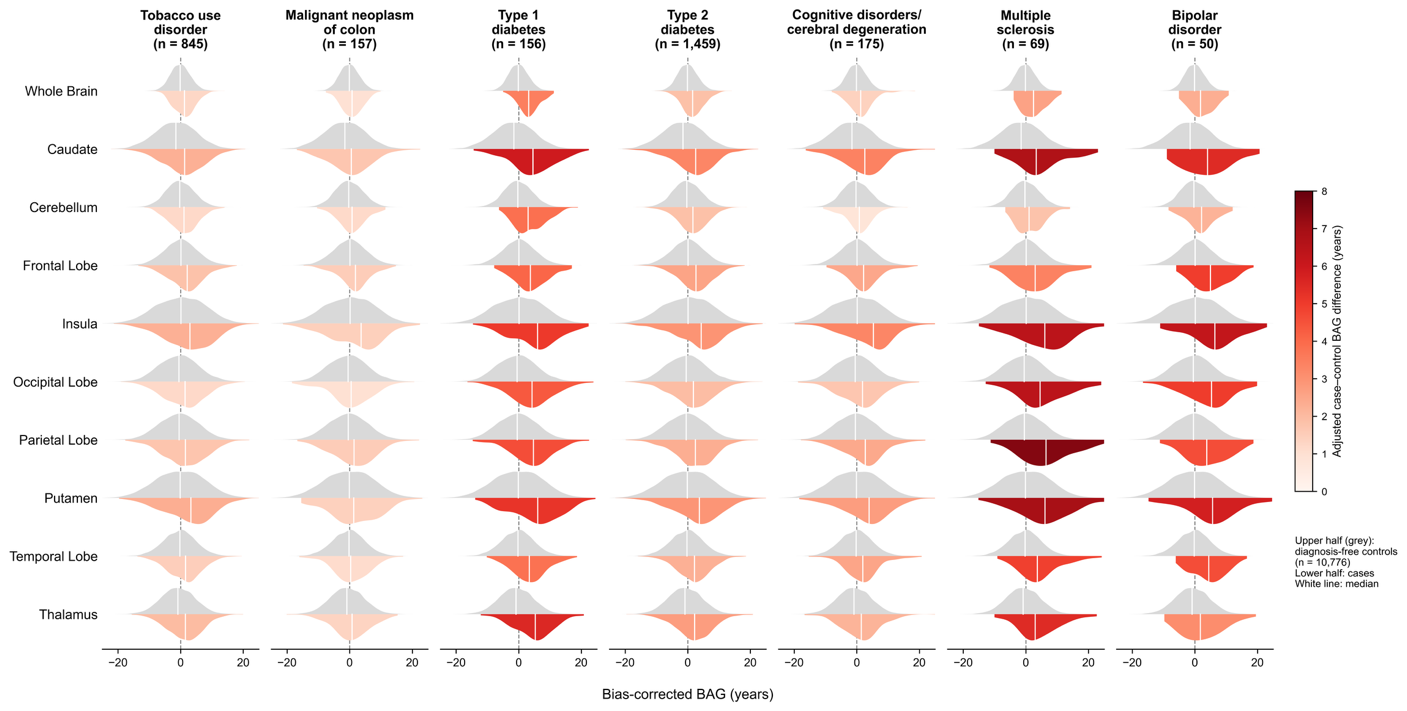

##### Supplementary Fig. 4 | Region-wise Manhattan and quantile–quantile plots for the UK Biobank BAG GWAS.

For each BAG, Manhattan and quantile–quantile plots show the 9,458,475 variants retained after filtering for analysis-sample MAF ≥ 0.01 and UK Biobank mfi INFO ≥ 0.8. Association analyses included 41,067 White British participants and used SAIGE v1.1.8 for null-model fitting and v1.1.9 for testing, adjusted for age, sex and ten genetic principal components, with a dense genetic relationship matrix and leave-one-chromosome-out analysis. The dashed line marks *P* = 5 × 10^-8^. λGC is the genomic-control inflation factor calculated for the plotted variant set; LDSC λGC is calculated for the HapMap3 regression subset in Supplementary Table 14.

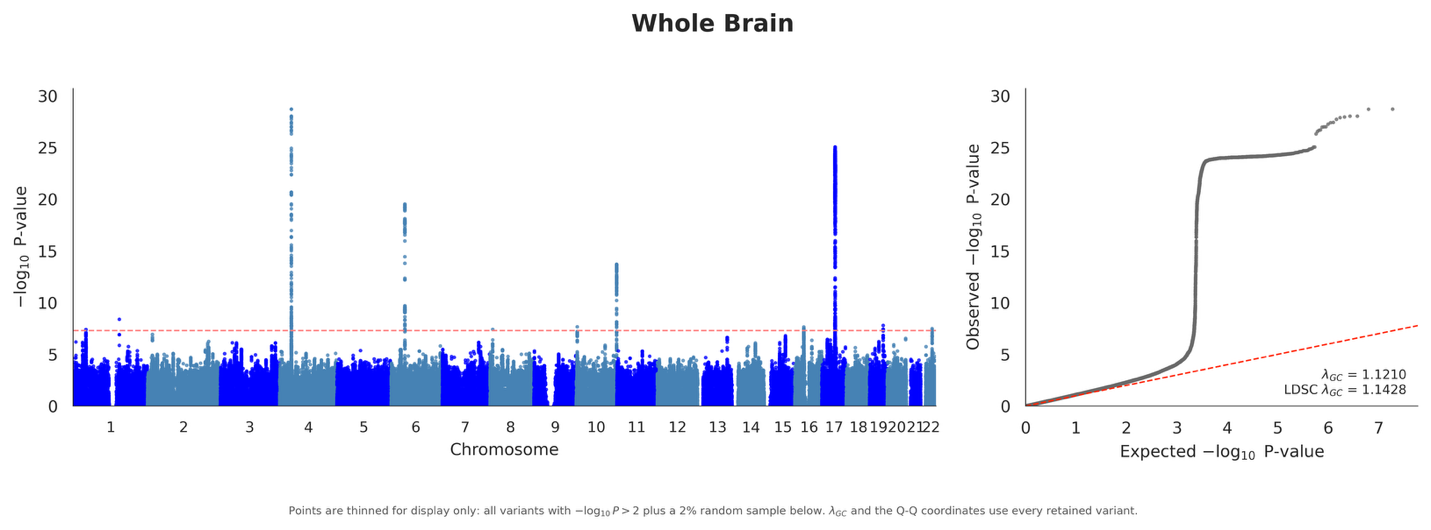

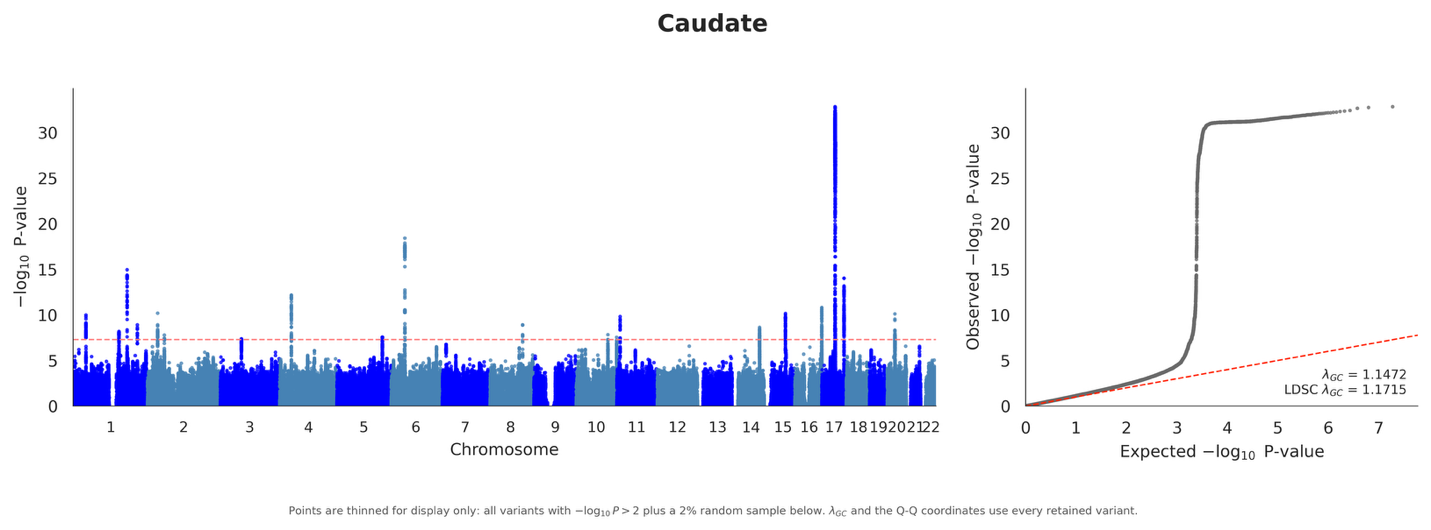

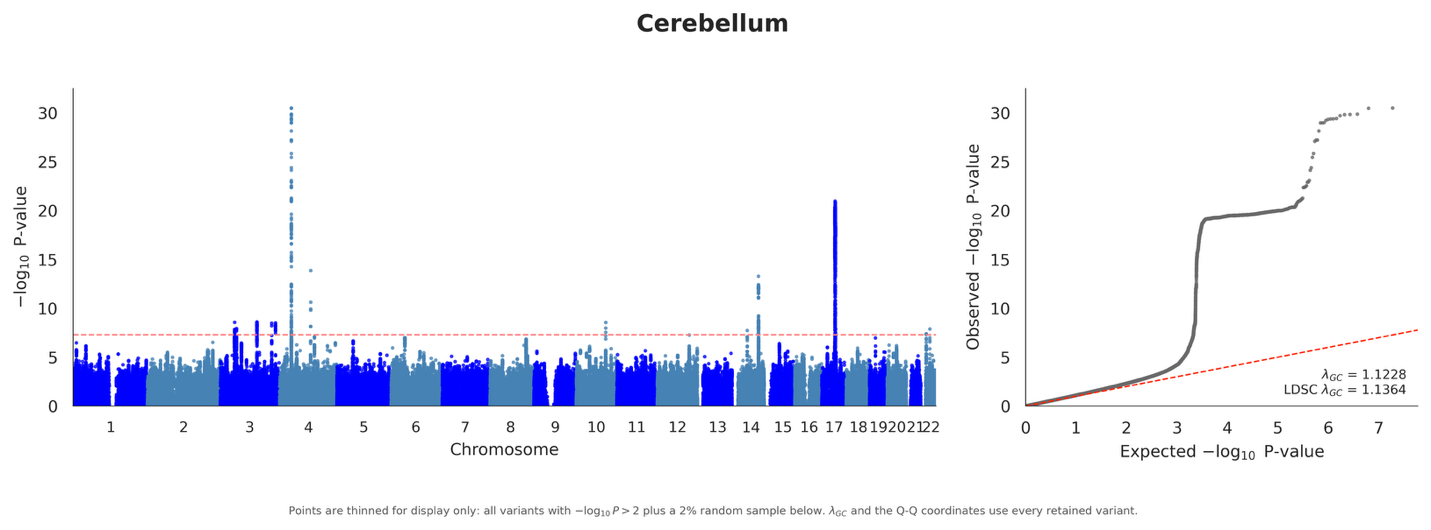

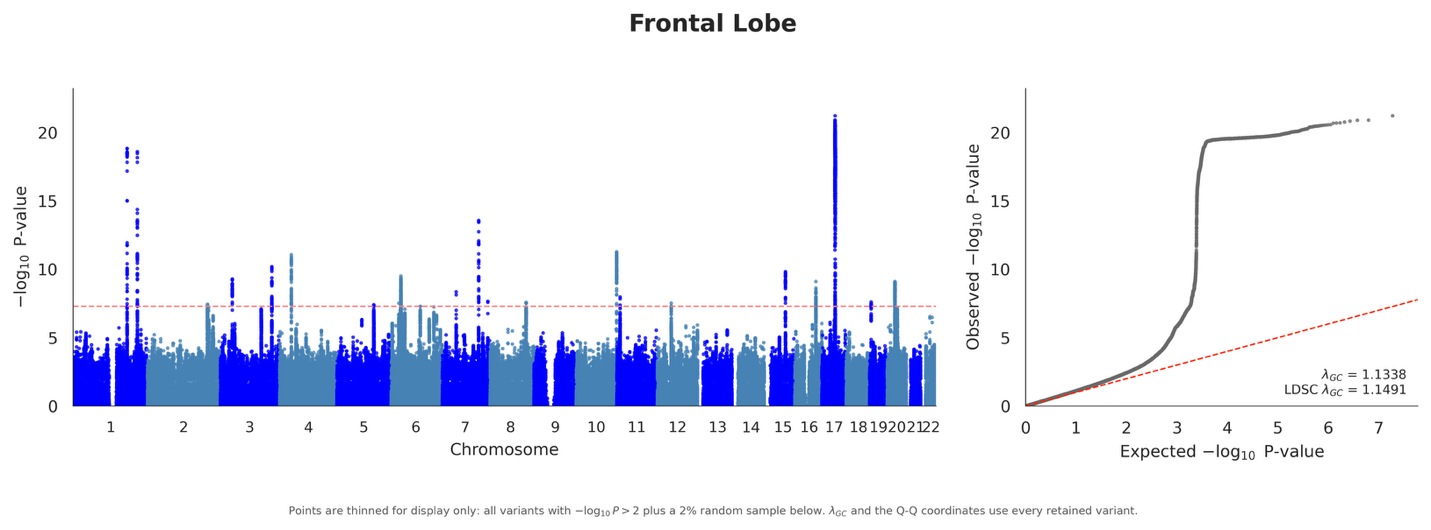

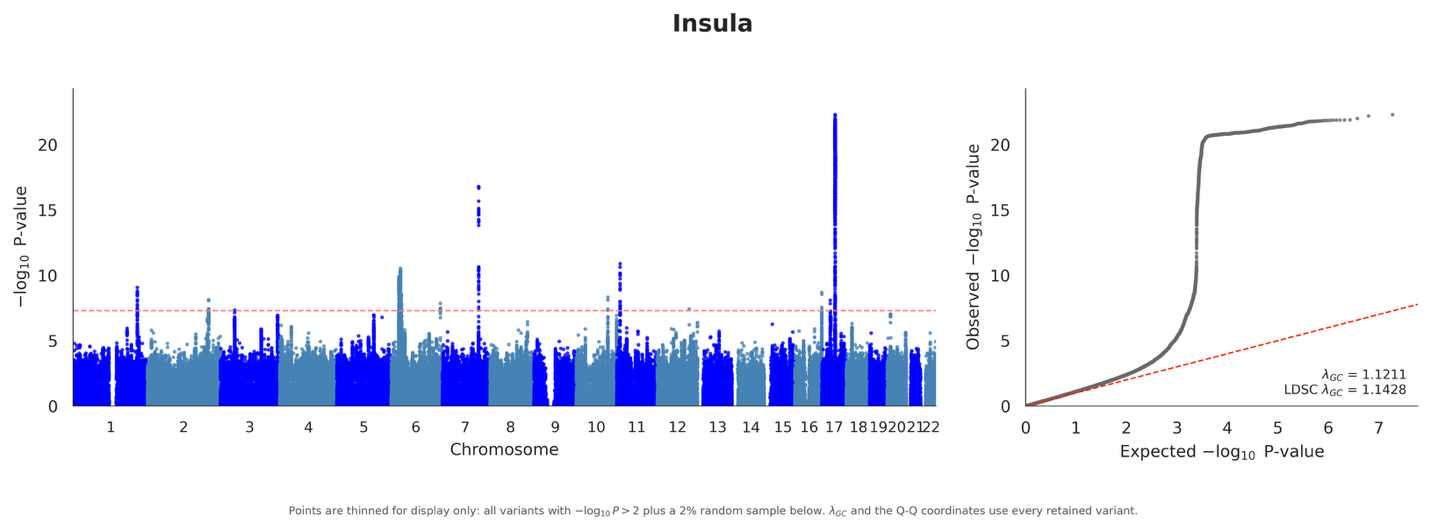

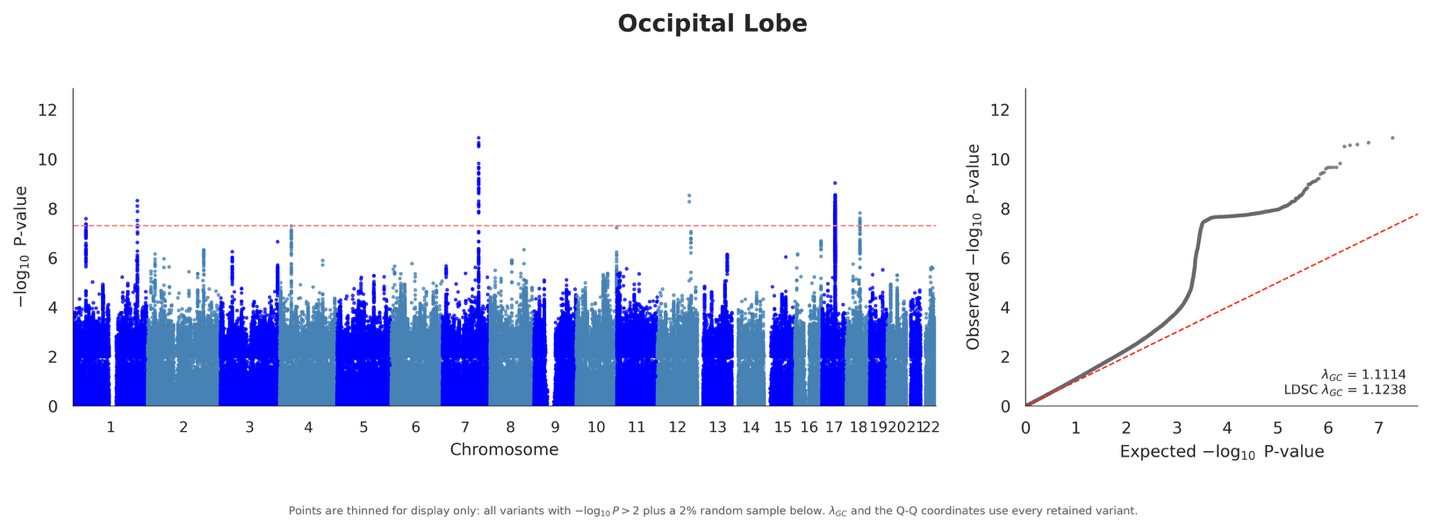

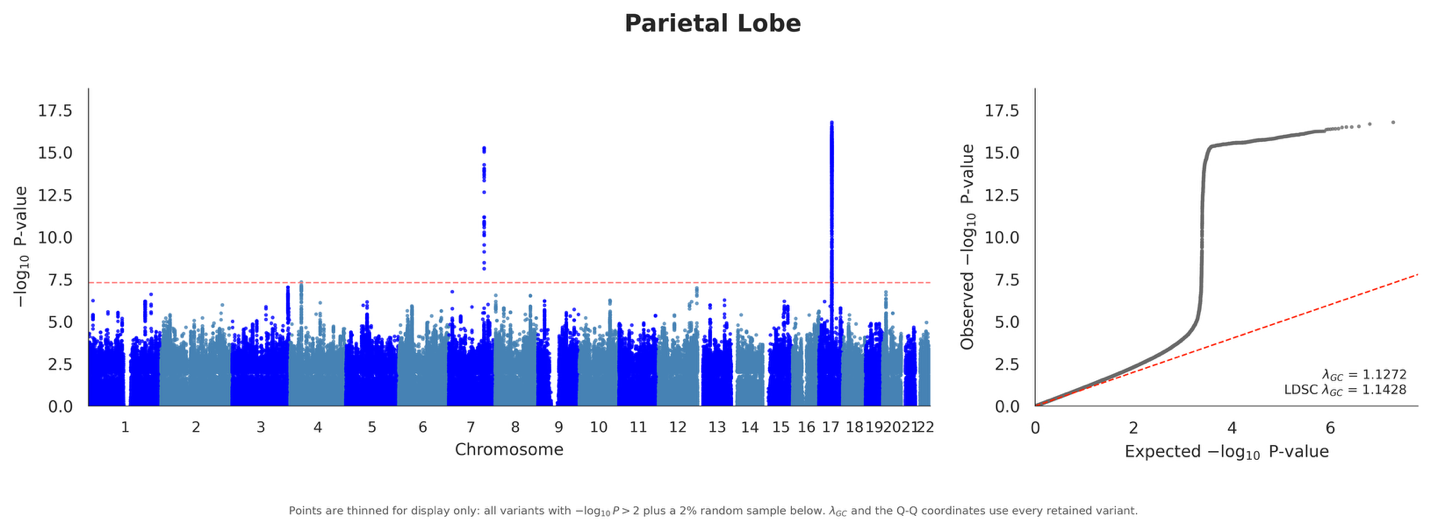

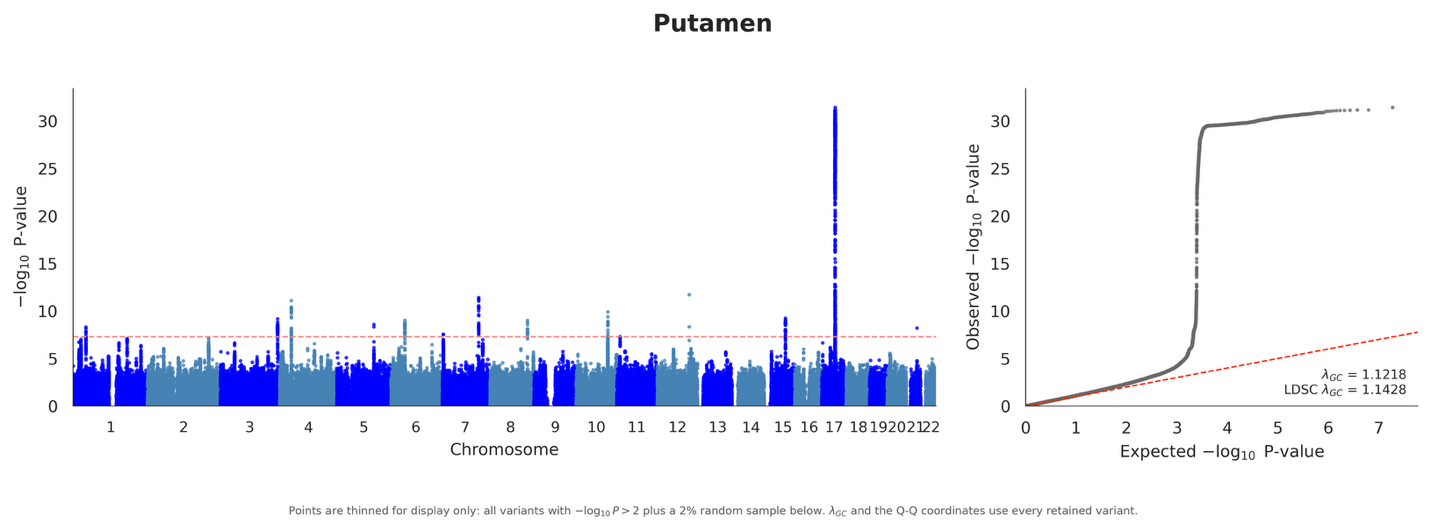

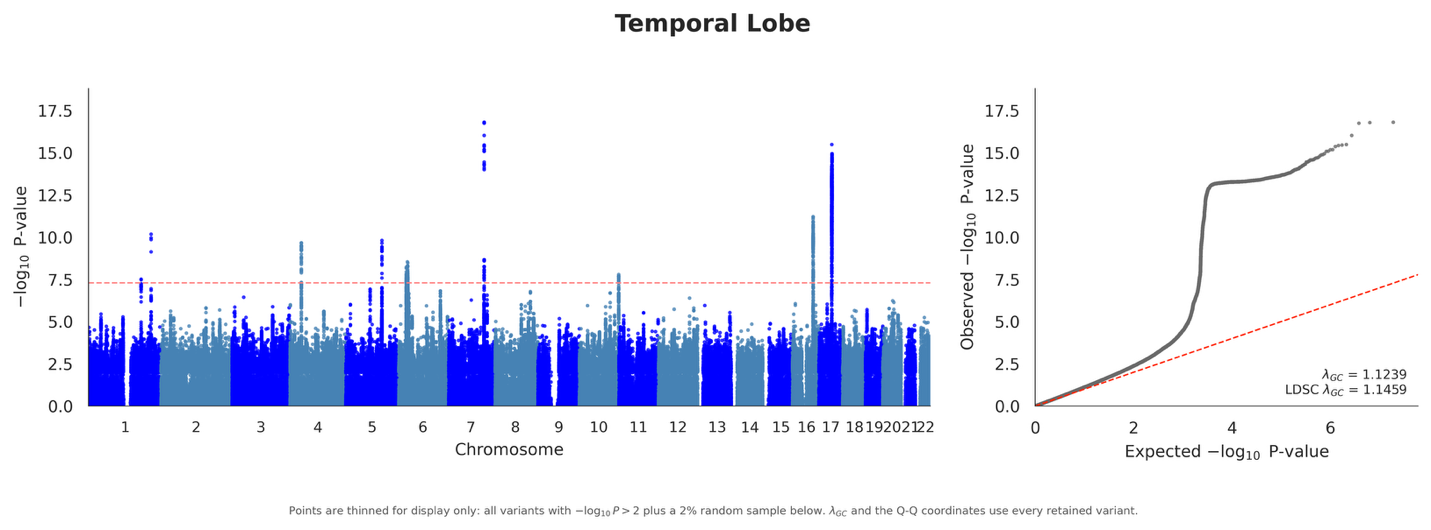

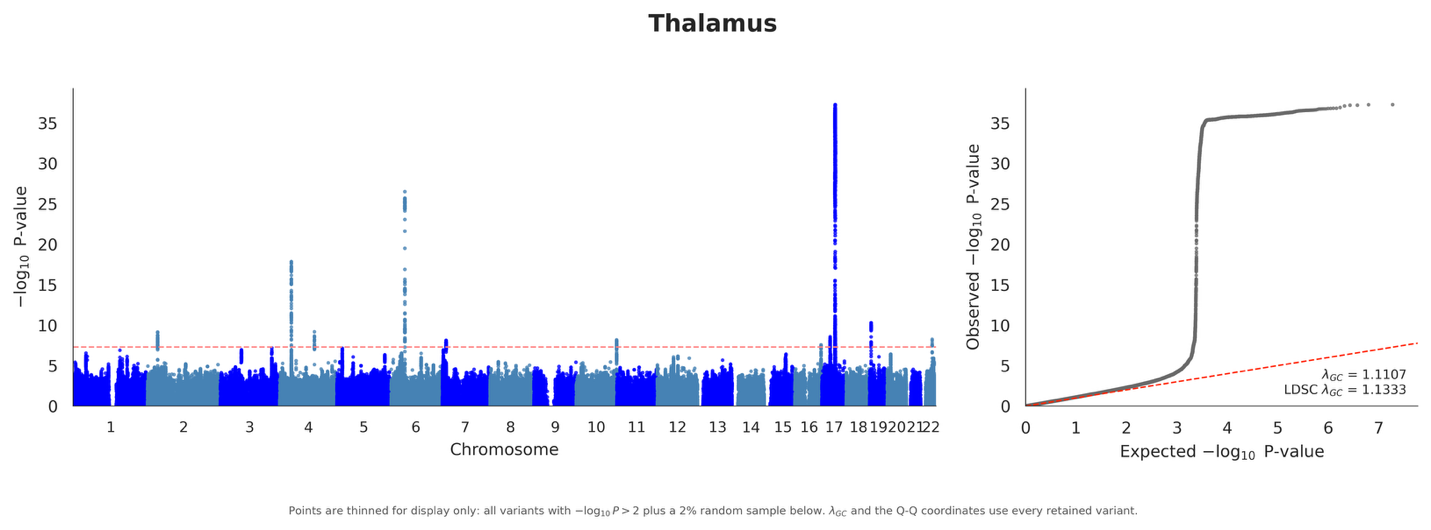

##### Supplementary Fig. 5 | Region-wise Manhattan and quantile–quantile plots for the ADNI BAG GWAS.

Association testing was performed with SAIGE in genetically inferred European-ancestry ADNI participants with genotype data and measured BAG (*n* = 1,693). Participants were assigned to the European-ancestry cluster by projection onto the 1000 Genomes reference principal-component space, after which principal components were recomputed within the retained subset. Models were adjusted for age, sex, the first ten within-ancestry principal components, and four genotyping-batch indicators, with ADNI1 used as the reference batch. BAG phenotypes were rank-based inverse-normal transformed before testing. A common set of 7,910,956 variants was tested for each phenotype after quality control and application of a minor allele count threshold of 10. Alternating colours distinguish adjacent chromosomes. In the Manhattan plots, the red dashed horizontal line denotes the genome-wide significance threshold (*P* = 5 × 10^-8^); in the Q–Q plots, the red dashed diagonal denotes the expected null distribution. Genomic control inflation factors ranged from 0.973 to 1.016 across phenotypes, showing no overall inflation in this sample (Supplementary Table 8). Genome-wide significant variants were observed for whole-brain (*n* = 7), temporal lobe (*n* = 10), thalamic (*n* = 3), and putaminal BAG (*n* = 1) (Supplementary Table 7). Locus-level results are described in Supplementary Note 2.

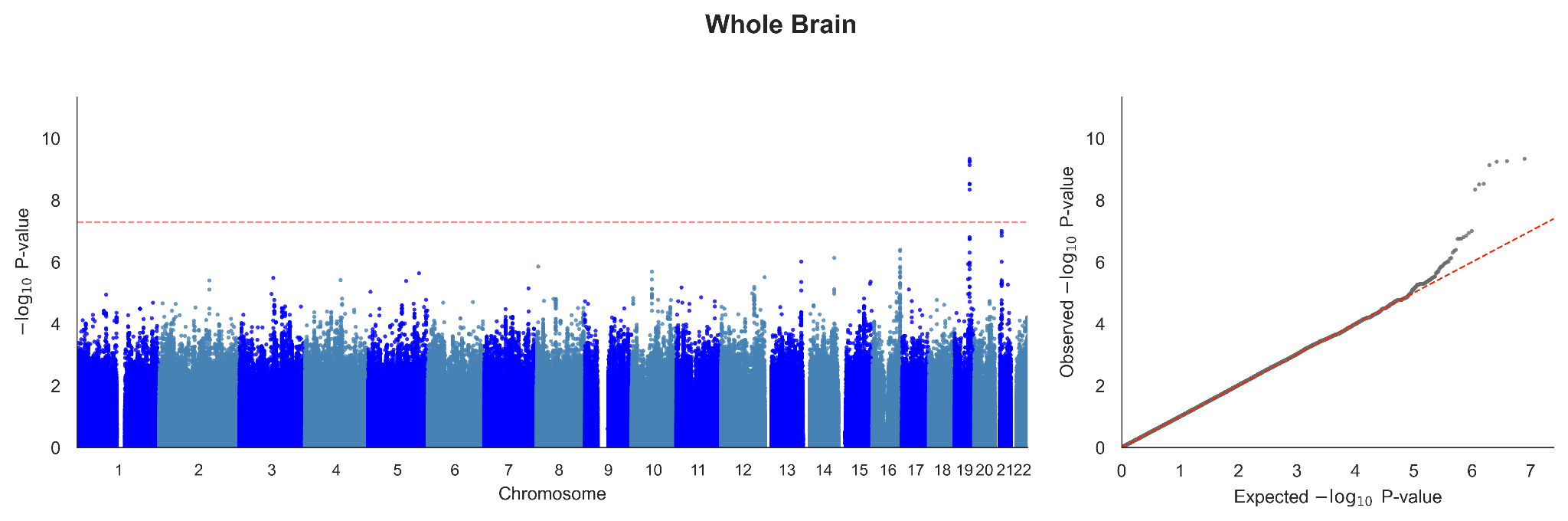

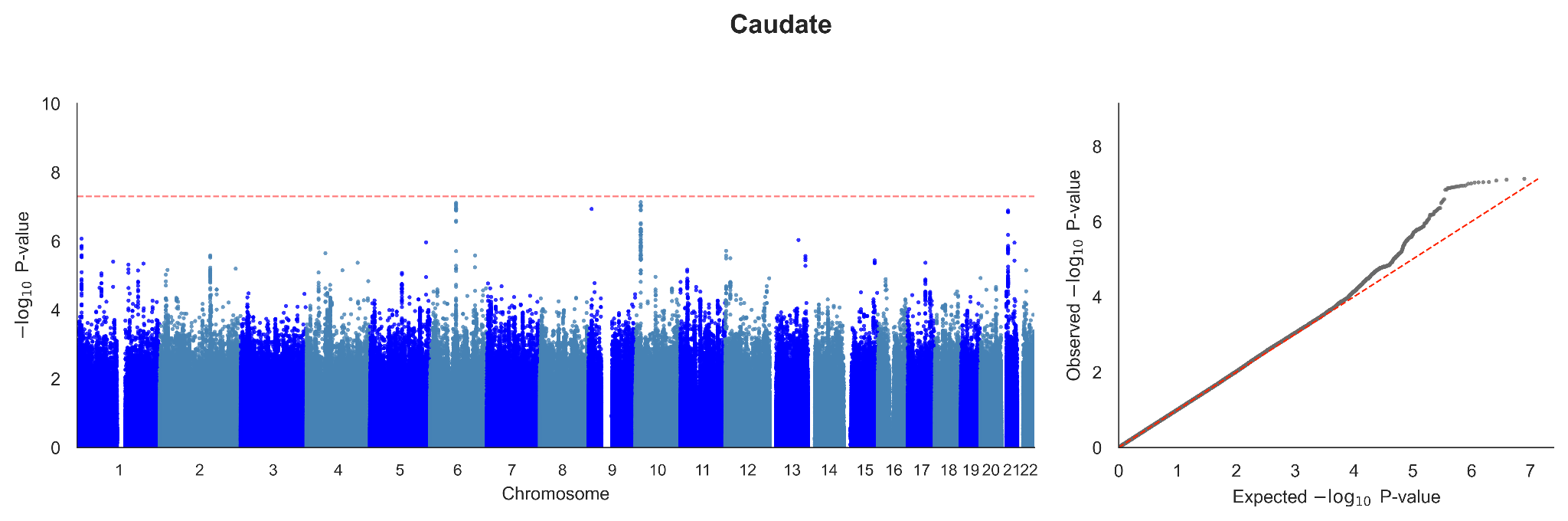

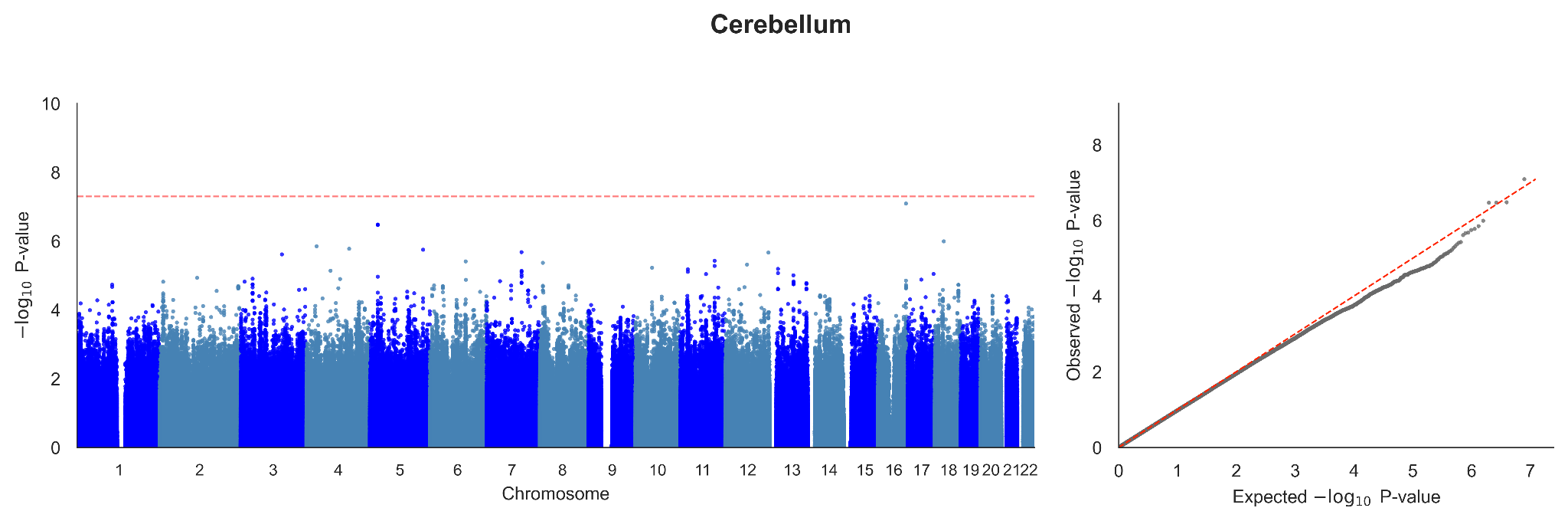

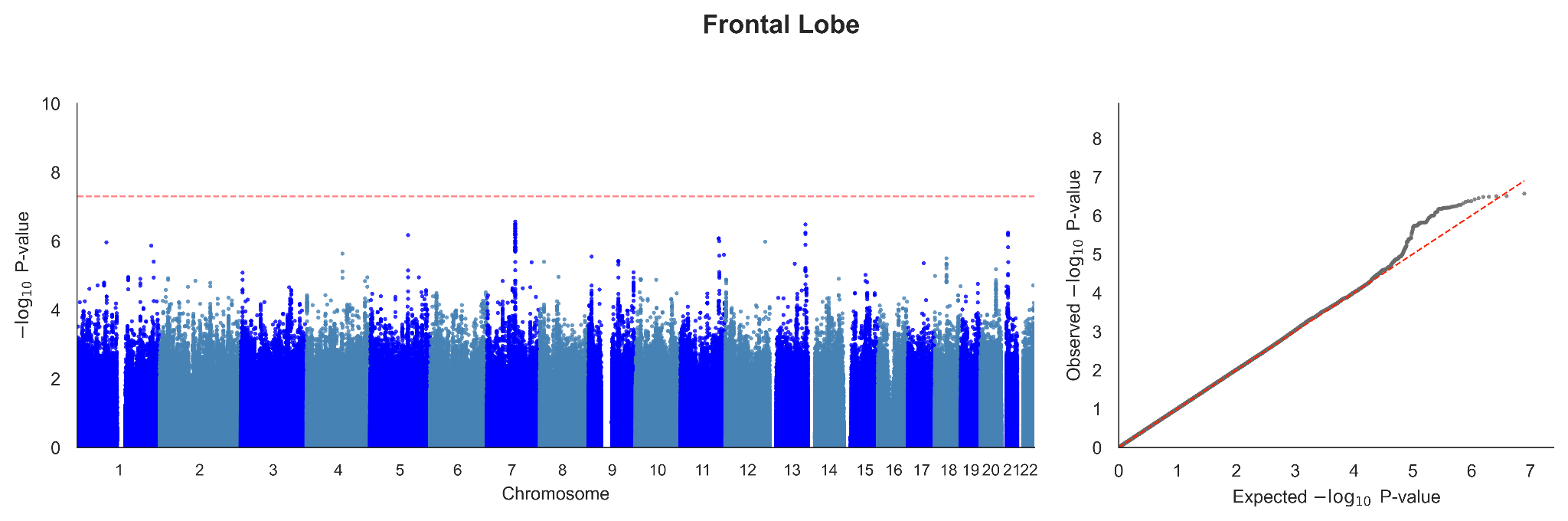

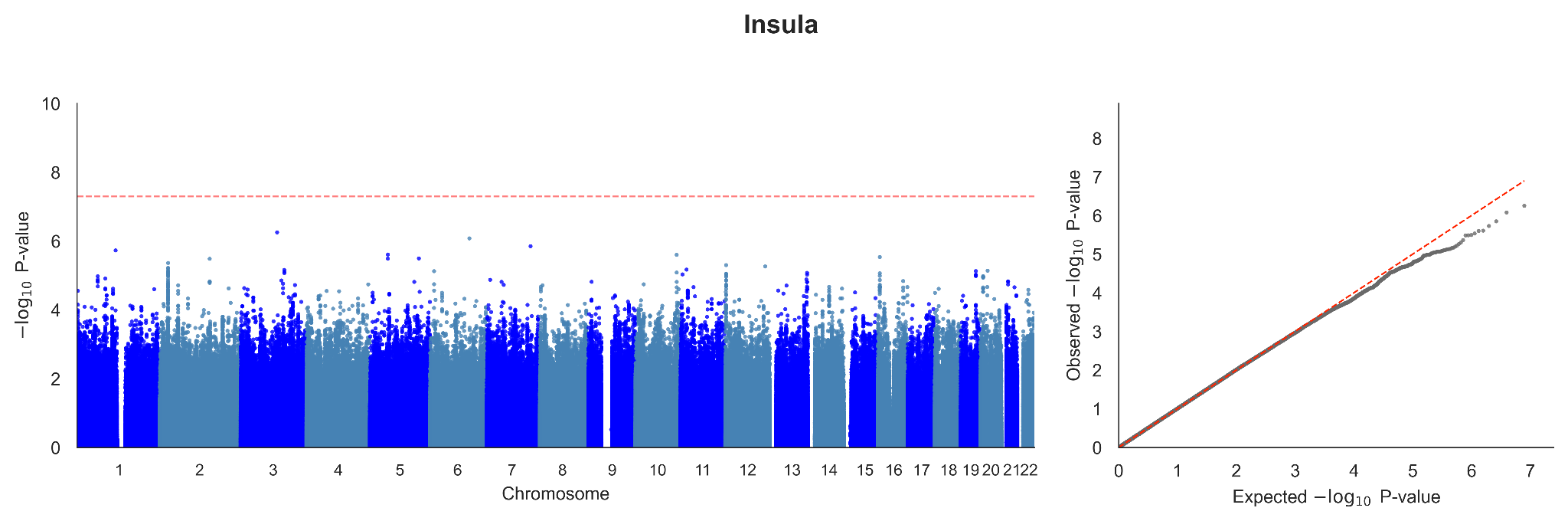

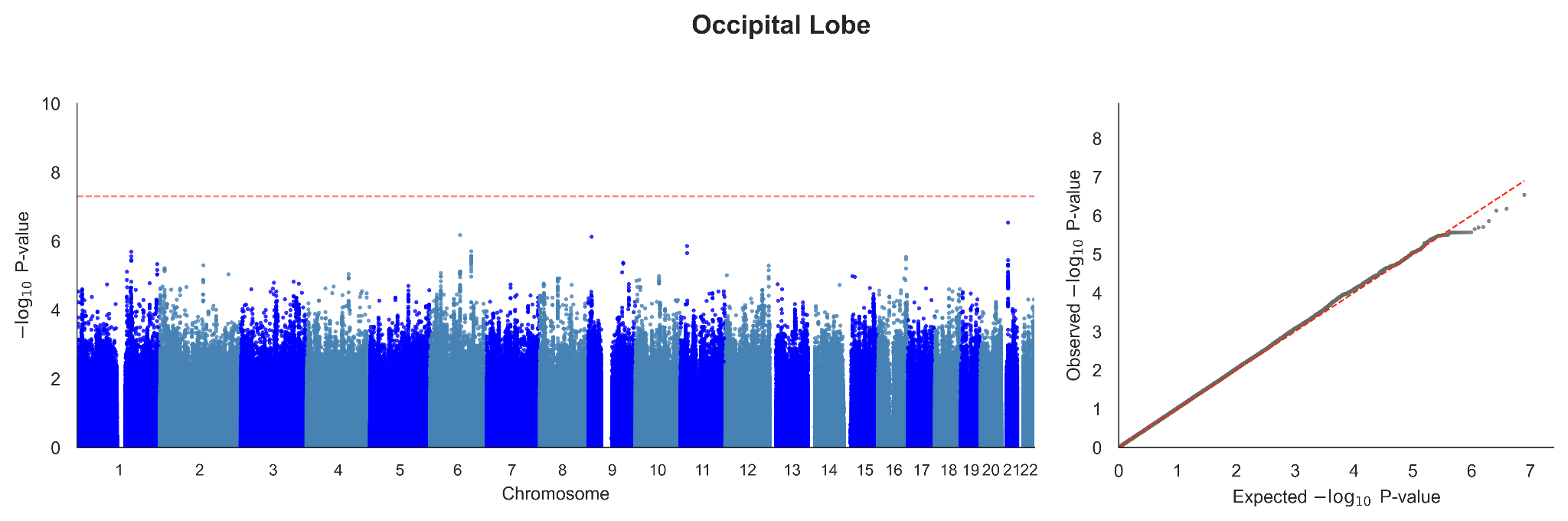

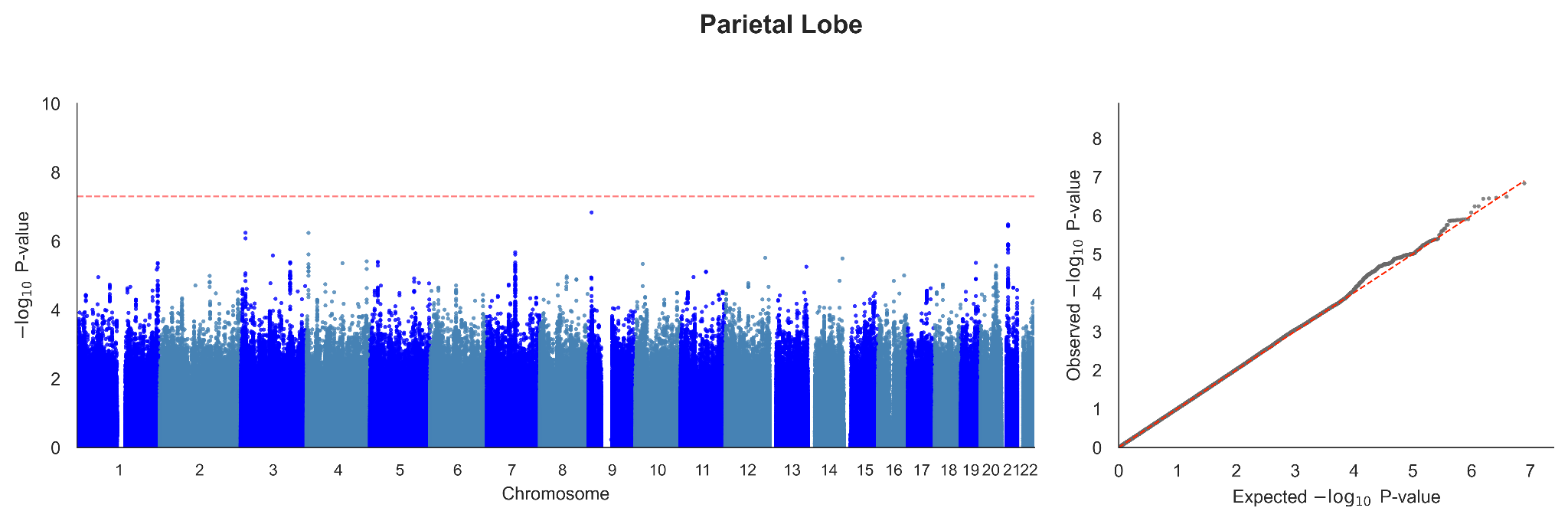

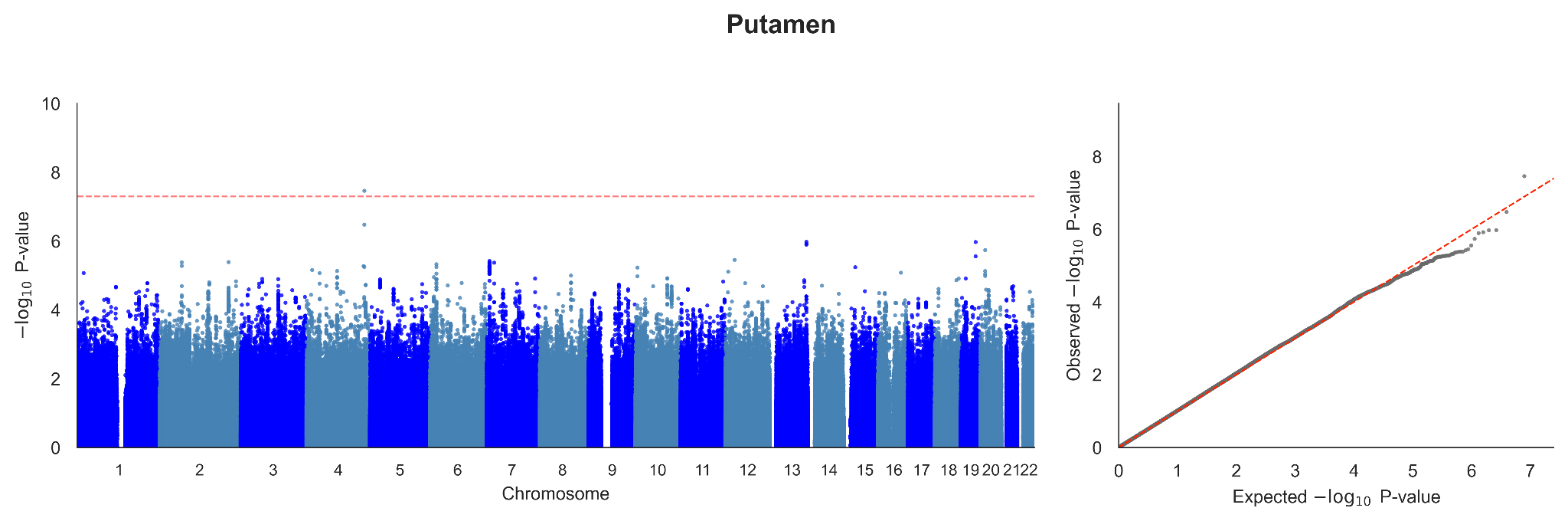

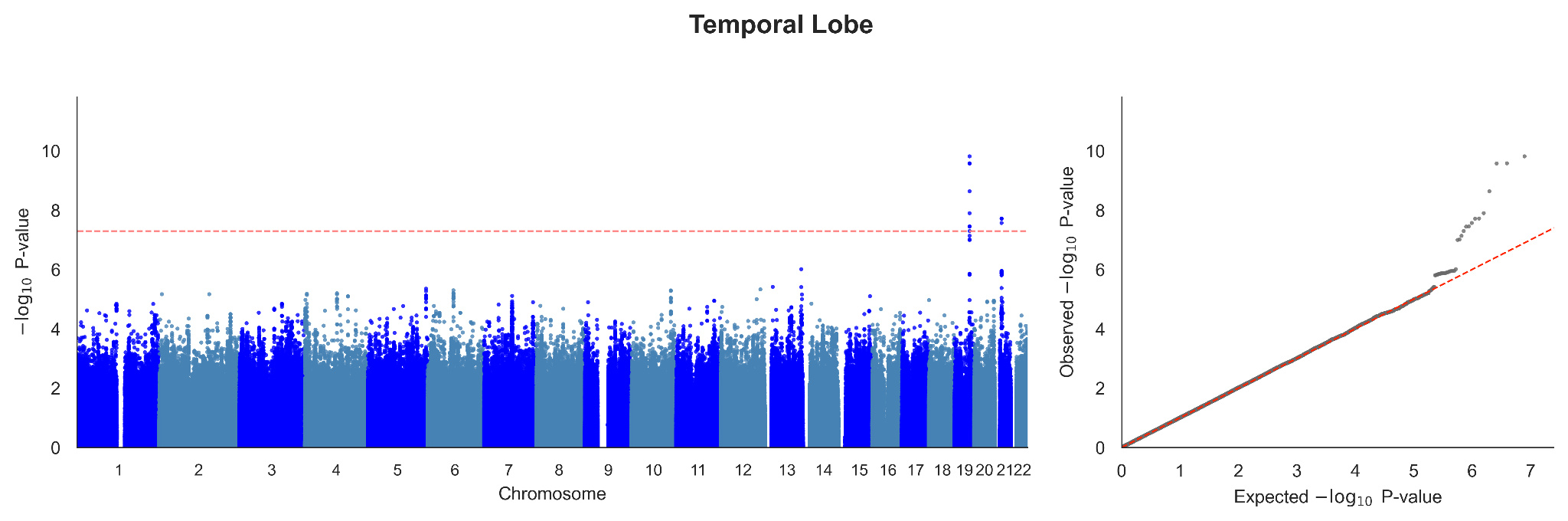

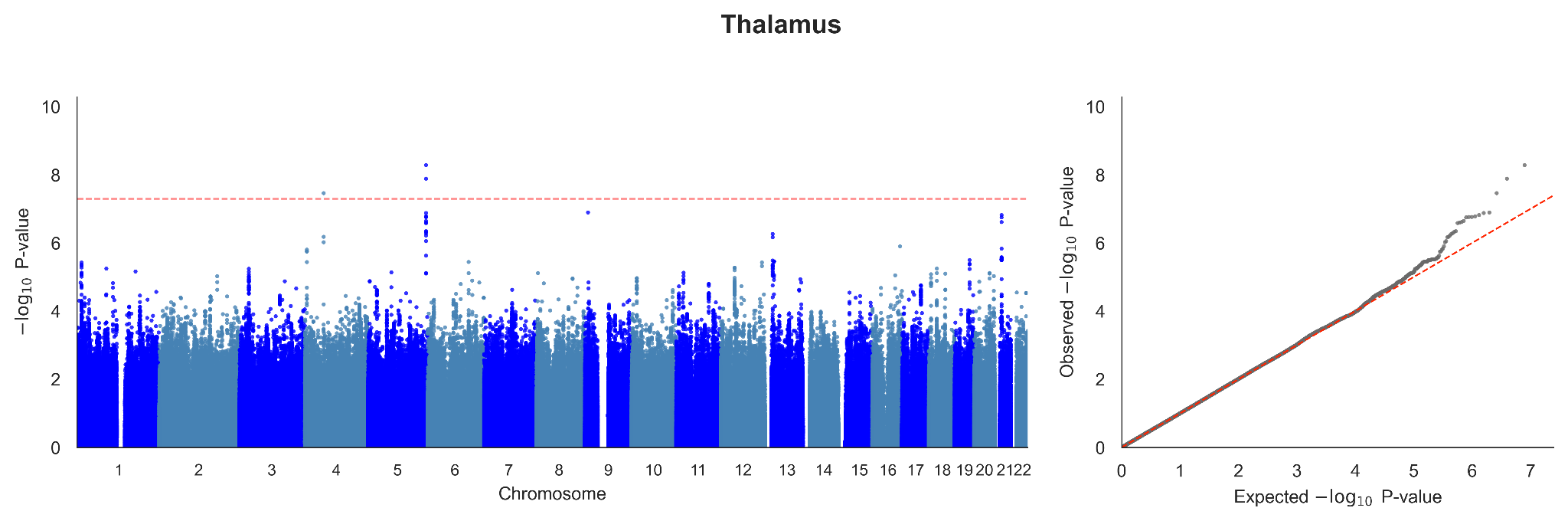

##### Supplementary Fig. 6 | UK Biobank–ADNI directional concordance of BAG GWAS lead effects.

Directional concordance of UK Biobank lead-variant effects in ADNI after effect-allele harmonization. Orange and blue points correspond to UK Biobank selection thresholds of *P* < 5 × 10^-8^ and *P* < 1 × 10^-5^, respectively. Points show concordance percentages, horizontal bars Wilson 95% confidence intervals, and the dashed line the 50% null expectation. Labels give the number of matched variants. Asterisks denote uncorrected two-sided binomial *P* values (**P* < 0.05; ***P* < 0.01; ****P* < 0.001). No BAG passed Bonferroni correction (*P* < 0.005) at the genome-wide selection threshold. Pooled summaries are descriptive because lead–region pairs are correlated (Supplementary Tables 9 and 10).

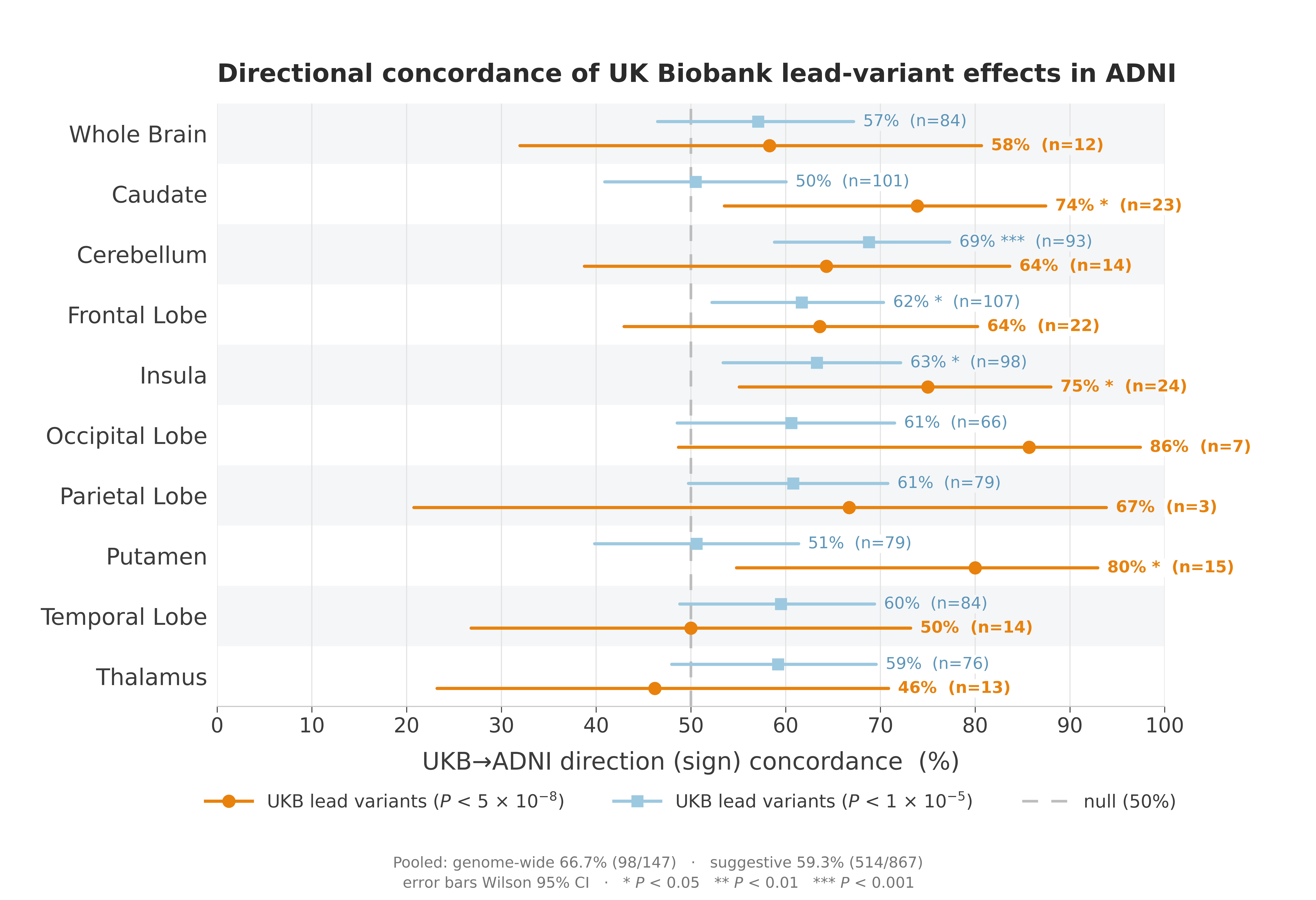

##### Supplementary Fig. 7 | Curated GWAS Catalog trait annotations of the mapped gene groups.

Rows show 27 curated Experimental Factor Ontology (EFO) trait categories; columns show 35 mapped gene groups, separated into 18 phenotype-restricted and 17 shared groups. Within each panel, groups are ordered by the number of BAGs represented and then by total annotated trait count. Cells report distinct GWAS Catalog traits per category and gene group; blank cells indicate none. Counts are descriptive, are not enrichment statistics and are not independent because traits may annotate multiple groups. Broad ‘Other measurement’ terms were reassigned to the displayed categories. Source annotations, keyword rules and curation methods are provided in Supplementary Tables 12 and 13 and Supplementary Note 3.

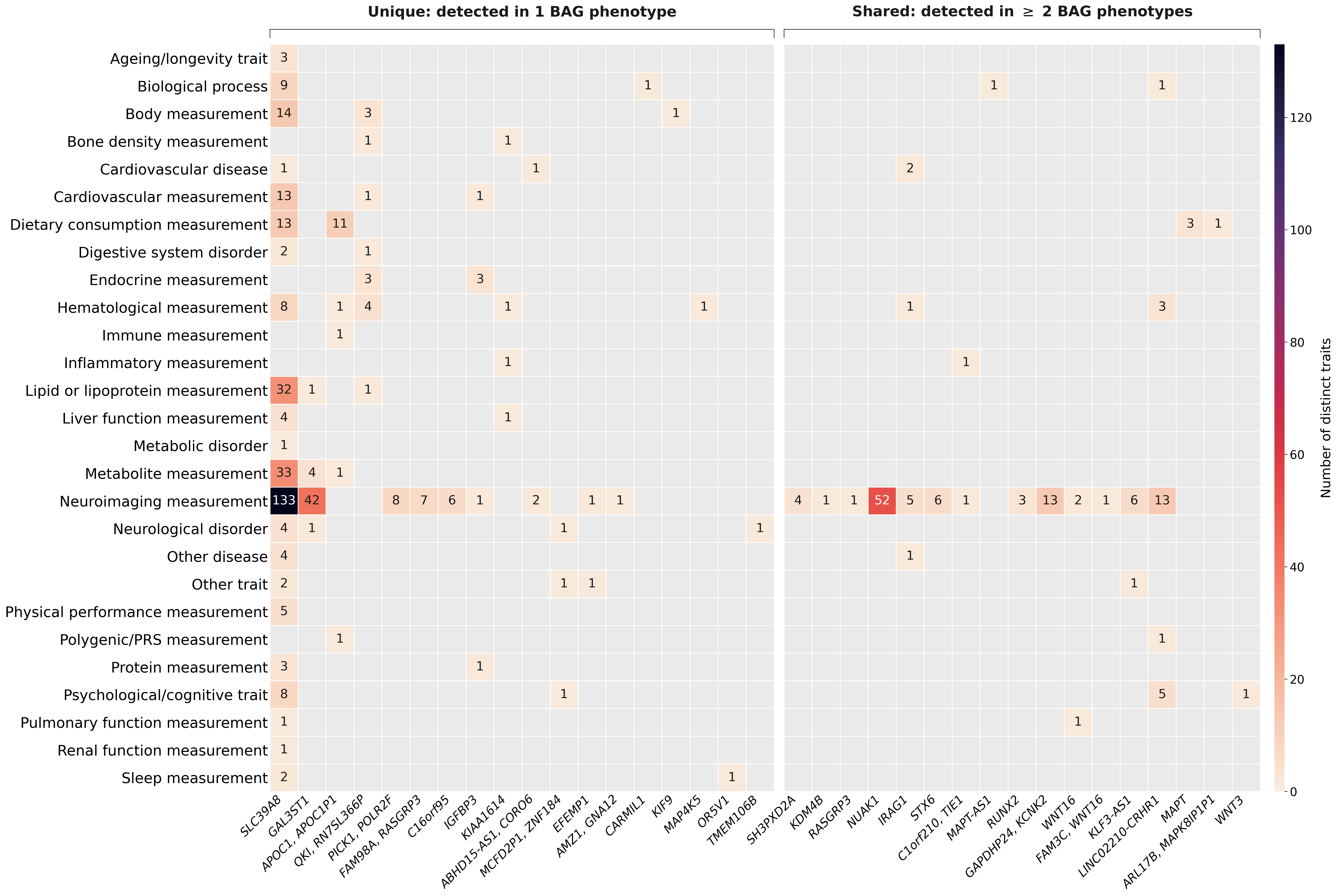

##### Supplementary Fig. 8 | UK Biobank validation of BAG-derived PRS.

Associations of UK Biobank-derived BAG PRS with inverse-normal-transformed BAGs in 9,997 relatedness-pruned held-out participants. *Left*, univariable linear regression of each BAG on its corresponding PRS, displayed on the unstandardized PRS scale, with the fitted line and 95% confidence band, adjusted R² and nominal regression *P* value. No covariates were included. *Right*, BAG distributions in the lowest and highest PRS deciles, compared using a two-sided Welch *t*-test; annotations report |*t*| and *P*. Boxes indicate the interquartile range, centre lines the median, and whiskers the most extreme values within 1.5 × the interquartile range; points beyond the whiskers are shown individually. Higher PRS was associated with higher BAG. Regression results are provided in Supplementary Table 17.

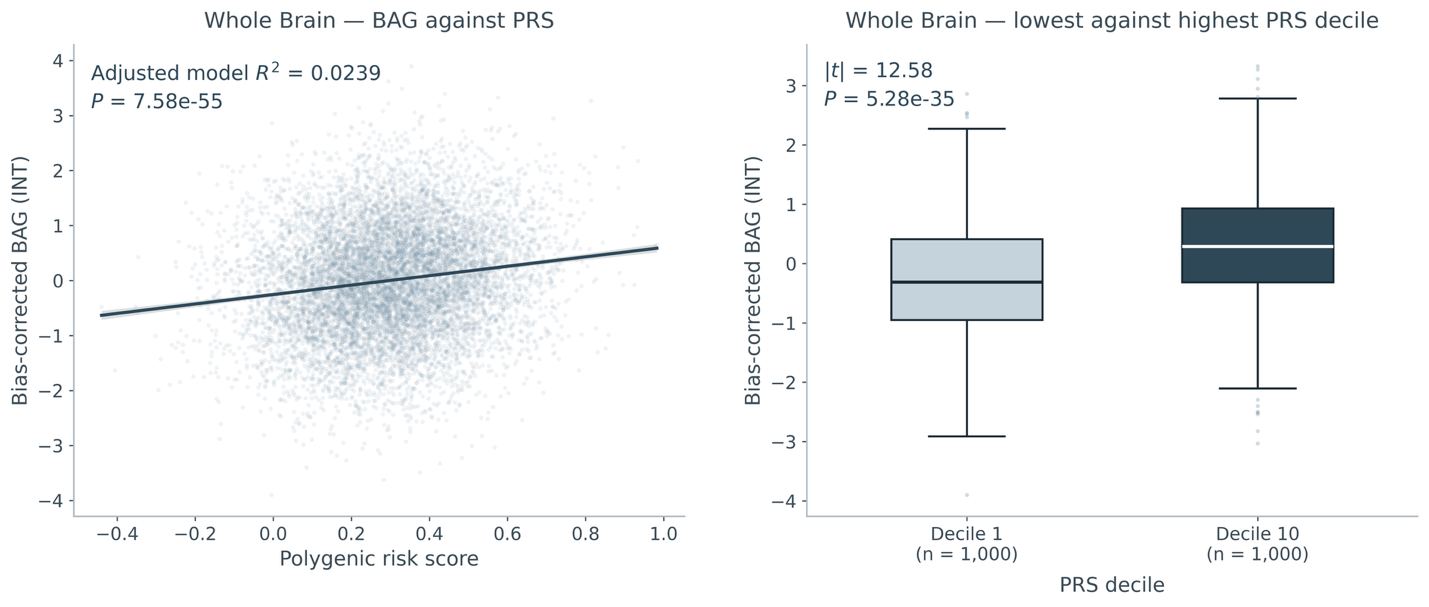

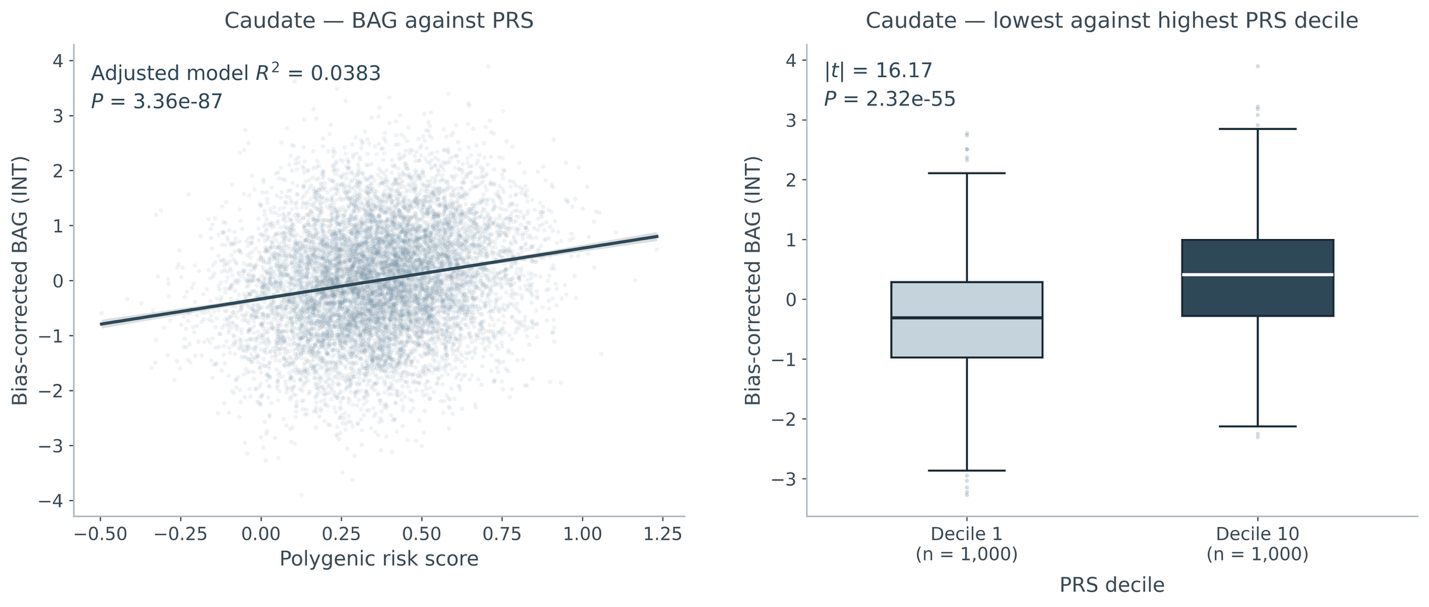

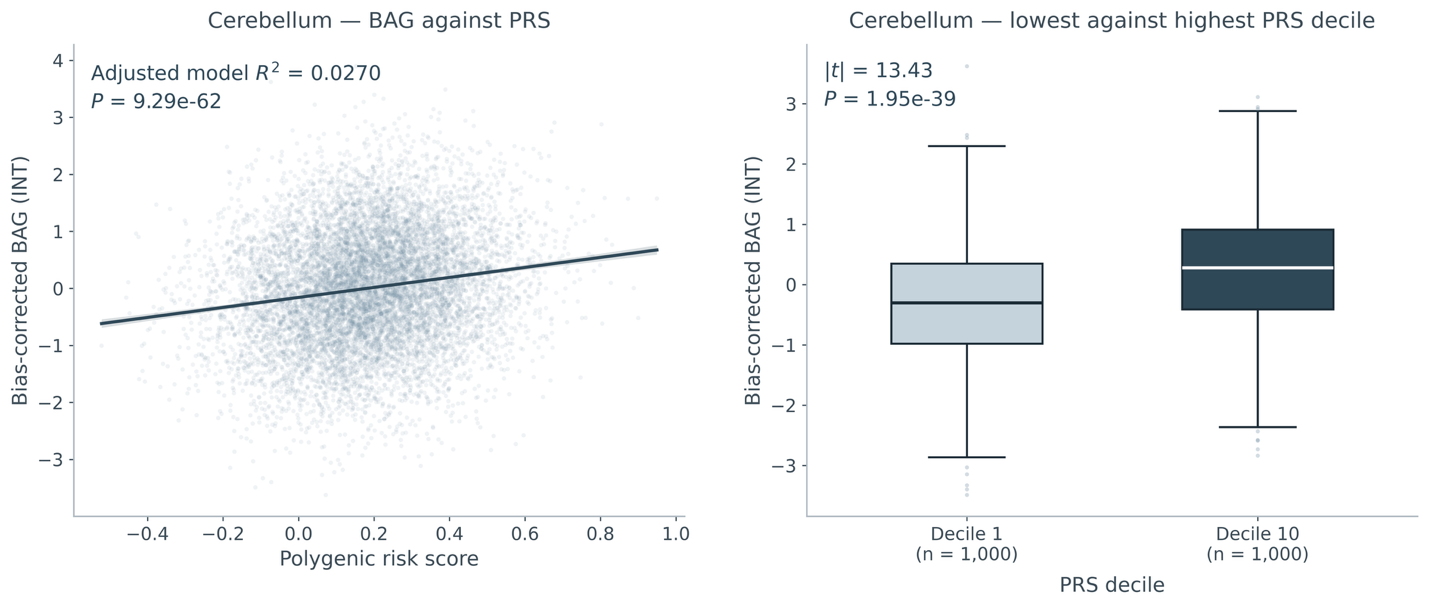

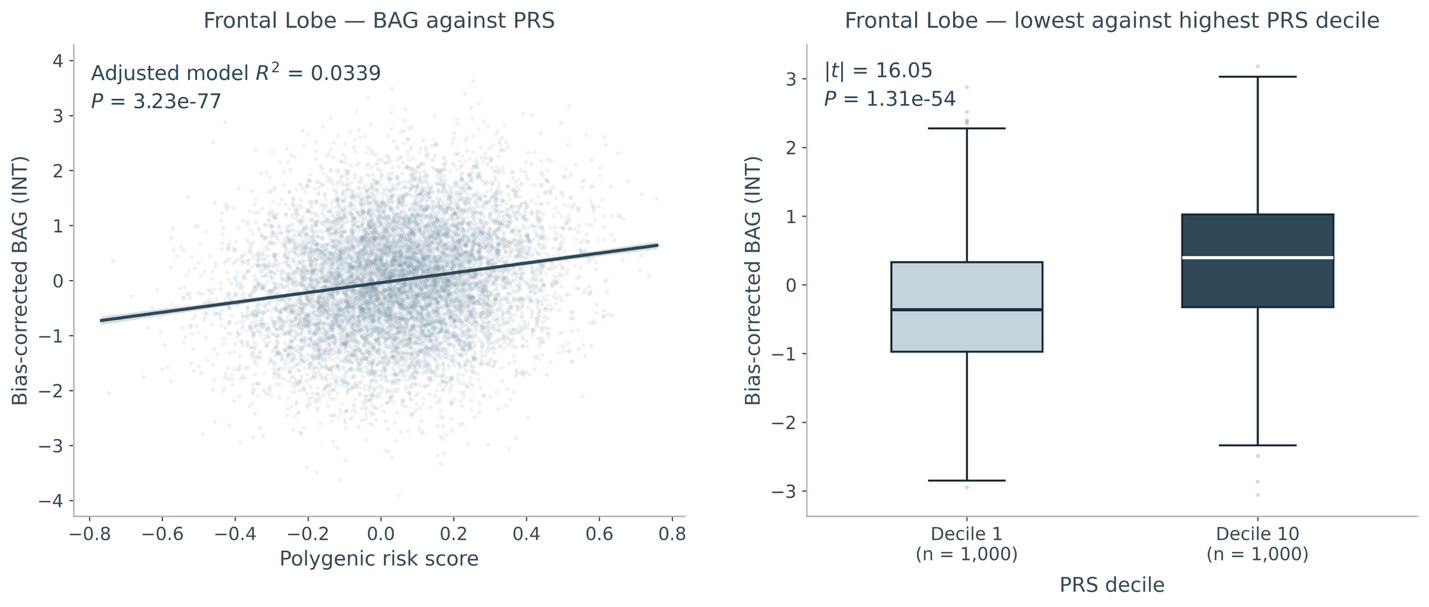

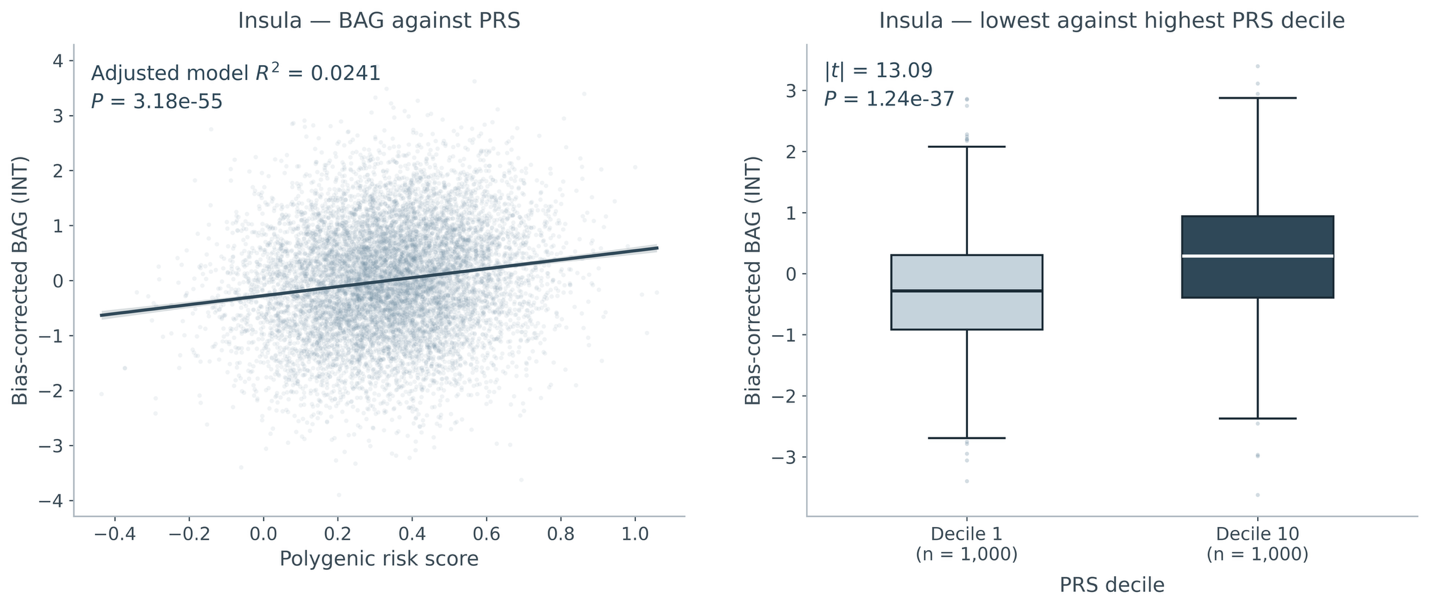

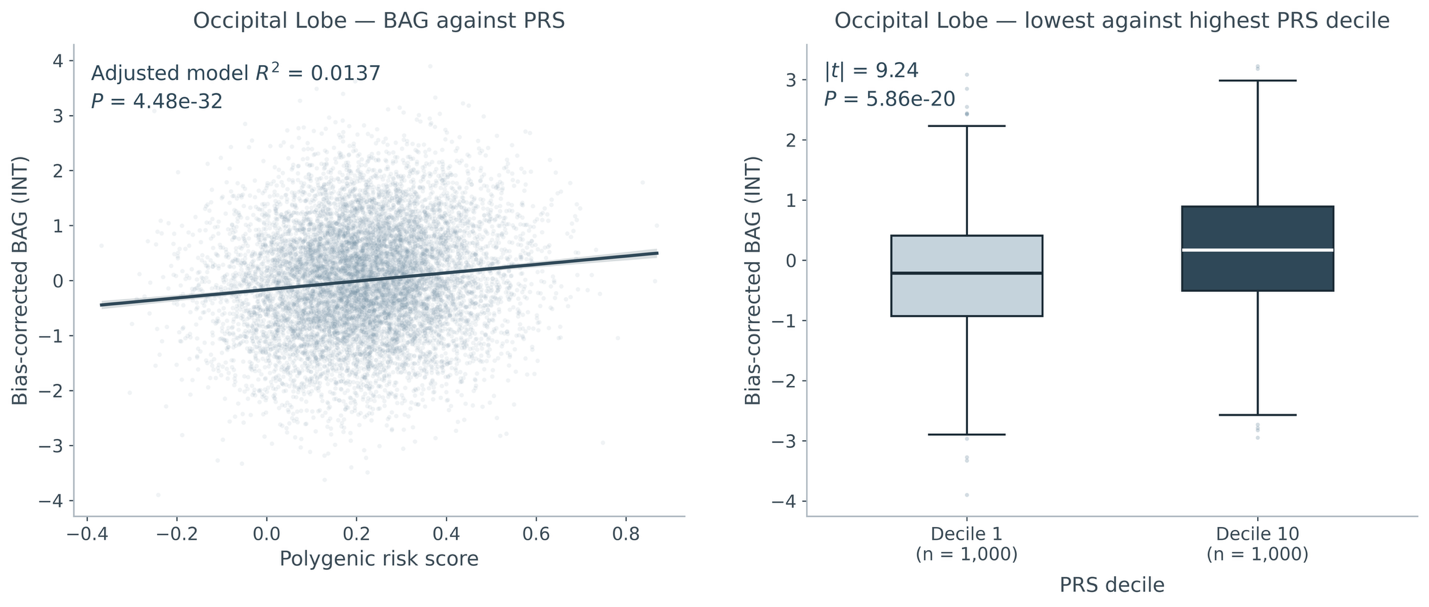

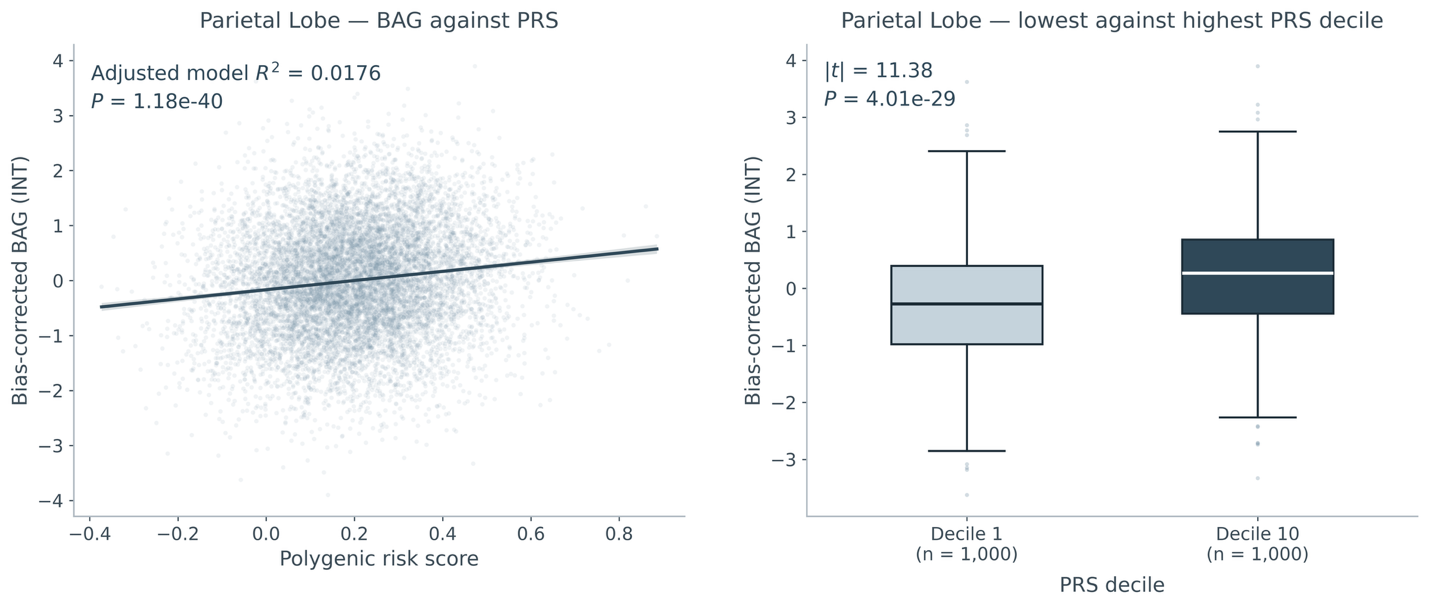

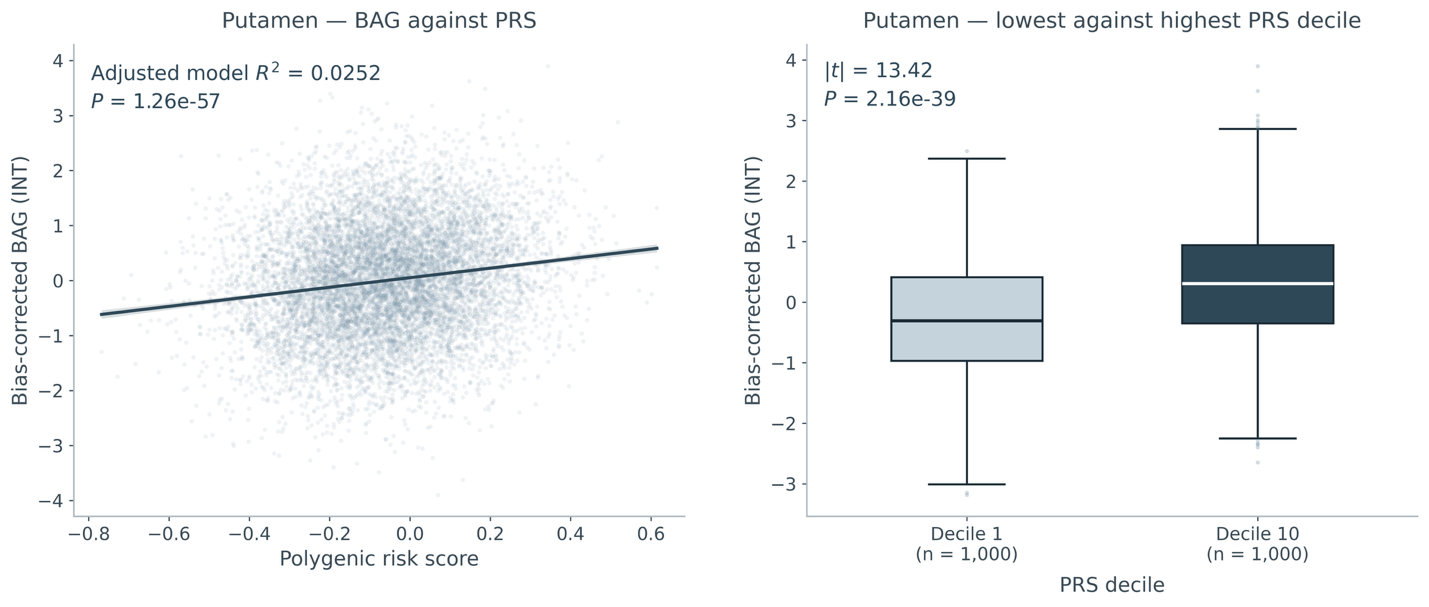

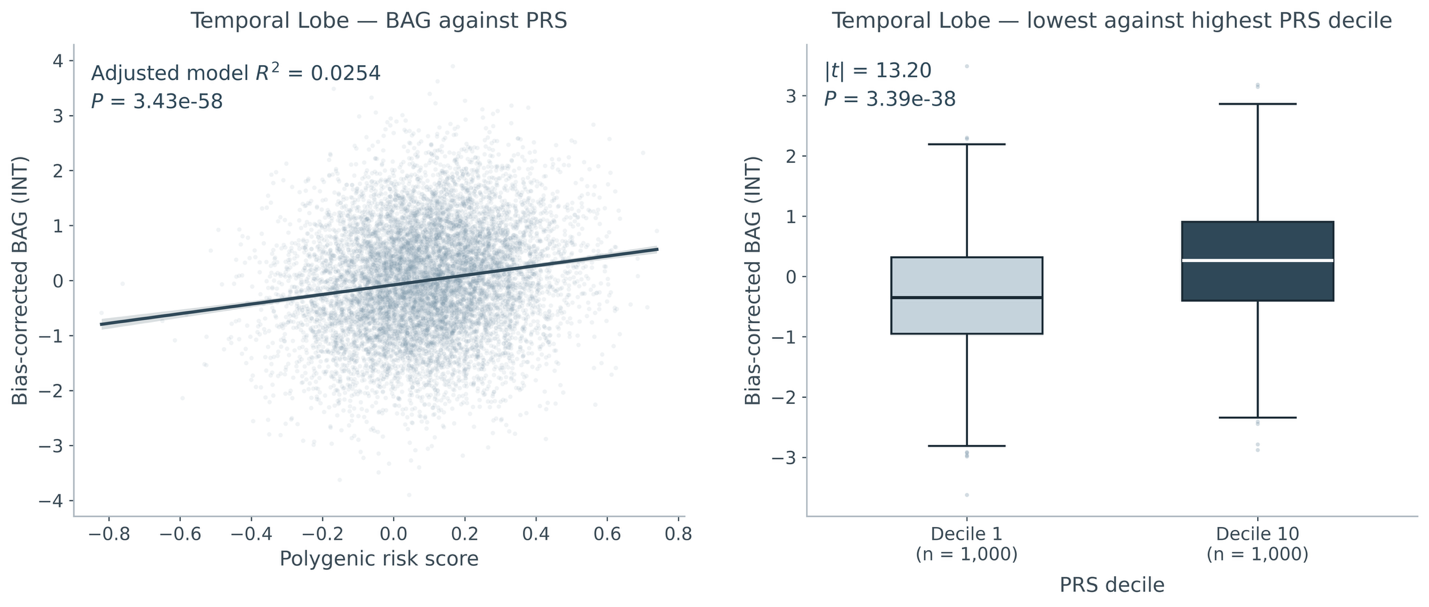

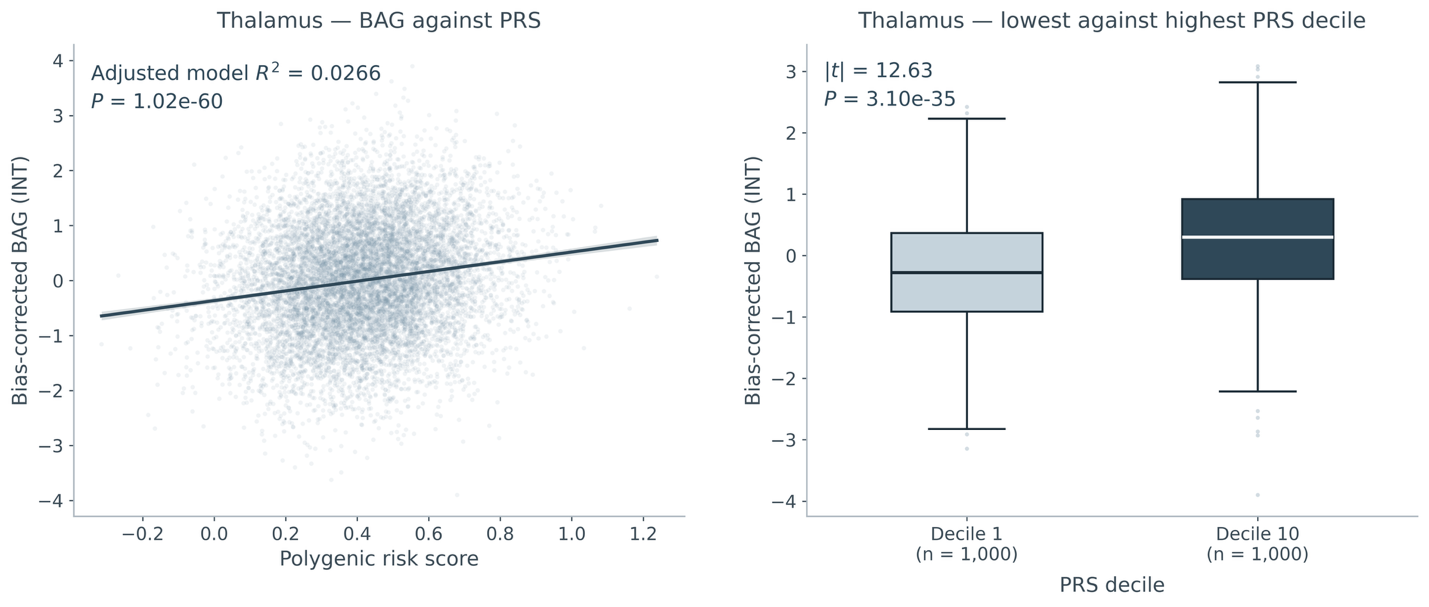

##### Supplementary Fig. 9 | ADNI validation of UK Biobank-derived BAG polygenic risk scores.

**a**, Pearson correlations between UK Biobank-derived BAG PRS and corresponding bias-corrected baseline BAGs in ADNI (*n* = 1,693); dots denote nominal *P* < 0.05. **b**, Associations with AD (329 cases) or MCI (865 cases), each compared with 651 cognitively normal participants. Logistic regression adjusted for age, sex, ten genetic principal components and *APOE* ε4 allele count. Points show odds ratios per one-unit increase in the unstandardized summed PRS, with 95% confidence intervals; the vertical line marks an odds ratio of 1. No clinical-label association survived correction across the 20 tests. Full results and sensitivity analyses without APOE ε4 adjustment are provided in Supplementary Tables 21–23.

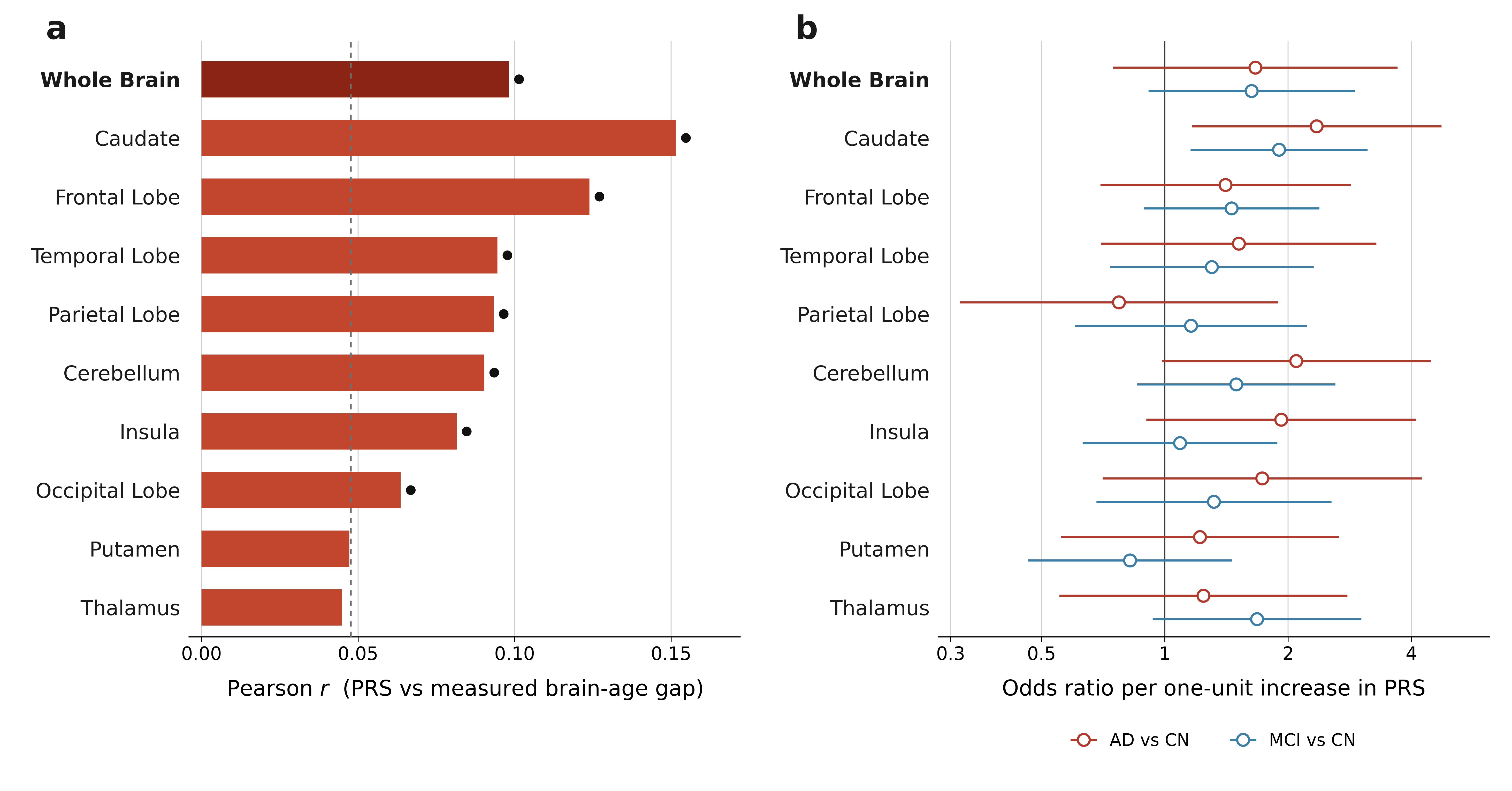

##### Supplementary Fig. 10 | Forward MR estimates for prioritized metabolic exposures and regional BAG.

Forest plots show inverse-variance-weighted (IVW) estimates for genetically proxied doctor-diagnosed diabetes and genetically proxied higher BMI against whole-brain and regional brain-age gap (BAG) phenotypes. These panels show the metabolic exposures highlighted in the main text. BAG outcomes were rank-based inverse-normal transformed; the IVW coefficients are therefore not expressed in years. Points indicate IVW effect estimates and horizontal bars indicate 95% confidence intervals. Square markers denote associations surviving Bonferroni correction across ten BAGs per exposure (*P* < 0.005; dark red; all also FDR-significant), circular orange markers denote associations surviving FDR correction only and circular grey markers denote associations not significant after correction. Doctor-diagnosed diabetes and BMI are shown in separate panels because their exposure scales differ: diabetes effects are on a linear-probability scale from a UK Biobank binary trait analysed with a linear mixed model, whereas BMI effects are per standard deviation. Full estimates and sensitivity diagnostics are provided in Supplementary Data 2.

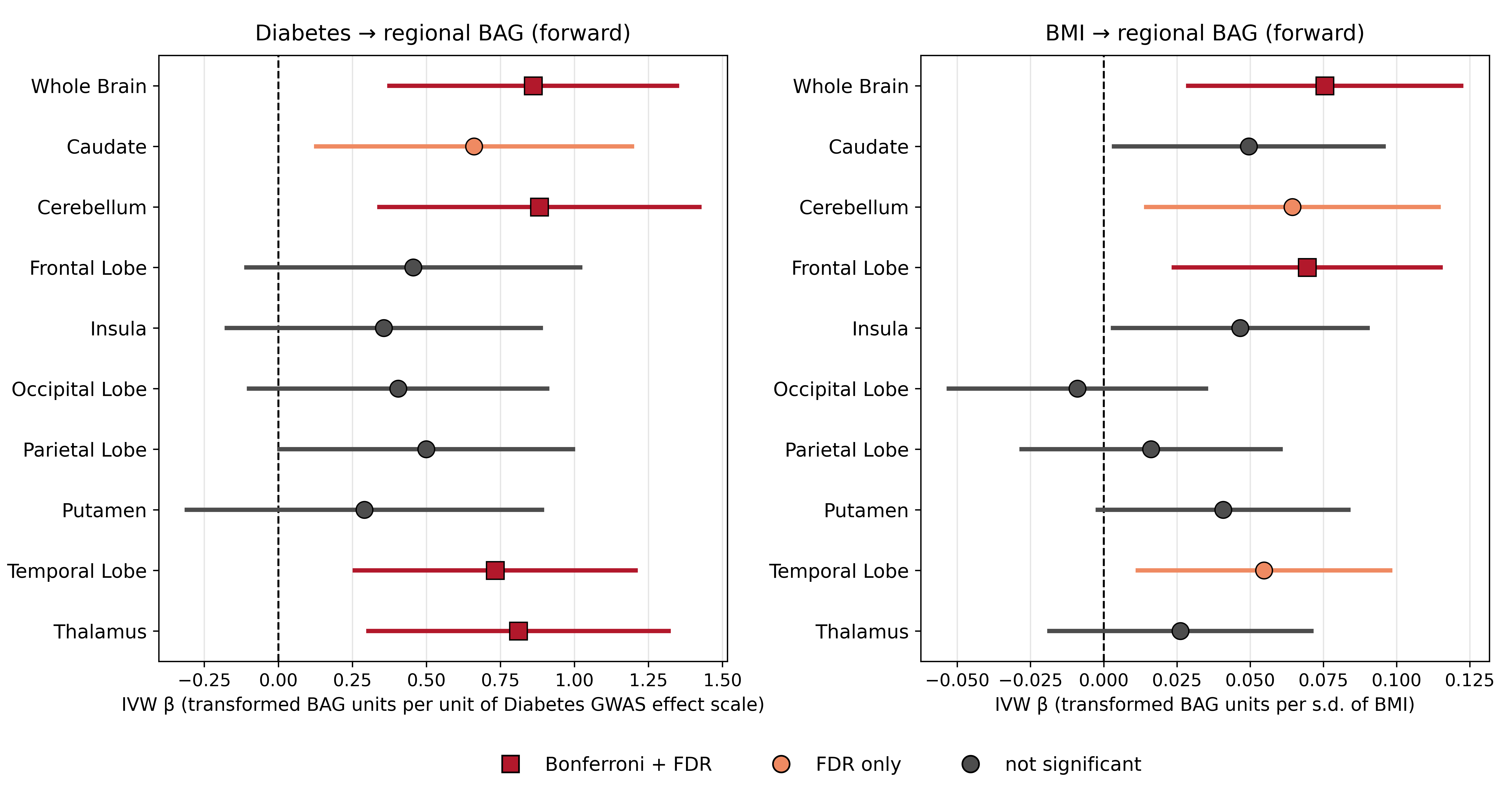

##### Supplementary Fig. 11 | Overview of bidirectional MR evidence across disease-related exposures and regional BAG.

Signed evidence-strength heatmap for forward (trait → BAG) and reverse (BAG → trait) MR across selected disease-related exposures and the whole-brain and nine regional BAG phenotypes. Diabetes, self-reported doctor-diagnosed diabetes; T1D, type 1 diabetes; HTN, hypertension; MS, multiple sclerosis; SCZ, schizophrenia; BD, bipolar disorder; FWD, trait-to-BAG MR; REV, BAG-to-trait MR. Tobacco denotes ever smoked, rather than tobacco use disorder. UKB mfi and 1000G EUR denote the UK Biobank imputation metadata and 1000 Genomes European allele-frequency references. Colour encodes sign(β) × −log₁₀(IVW P), clipped at −4 and +4; red and blue indicate positive and negative estimates, respectively. Colour represents statistical evidence, not effect magnitude. Boxed cells marked ‡ passed Bonferroni correction across ten BAGs per trait and direction (*P* < 0.005); † marks *q* < 0.05 without Bonferroni significance. “n.e.” denotes non-estimable results. The full grid contains eight ‡ cells, four † cells and ten non-estimable forward colon-cancer comparisons. Alzheimer’s disease is shown with two allele-frequency references in each direction (UKB mfi and 1000G EUR). For AD, effect estimates were reconstructed from z-scores, per-variant sample sizes and reference allele frequencies using the formulas in Supplementary Note 12; no liability-scale transformation was applied. These rows are included as a technical sensitivity analysis. Full estimates and sensitivity diagnostics are provided in Supplementary Data 2.

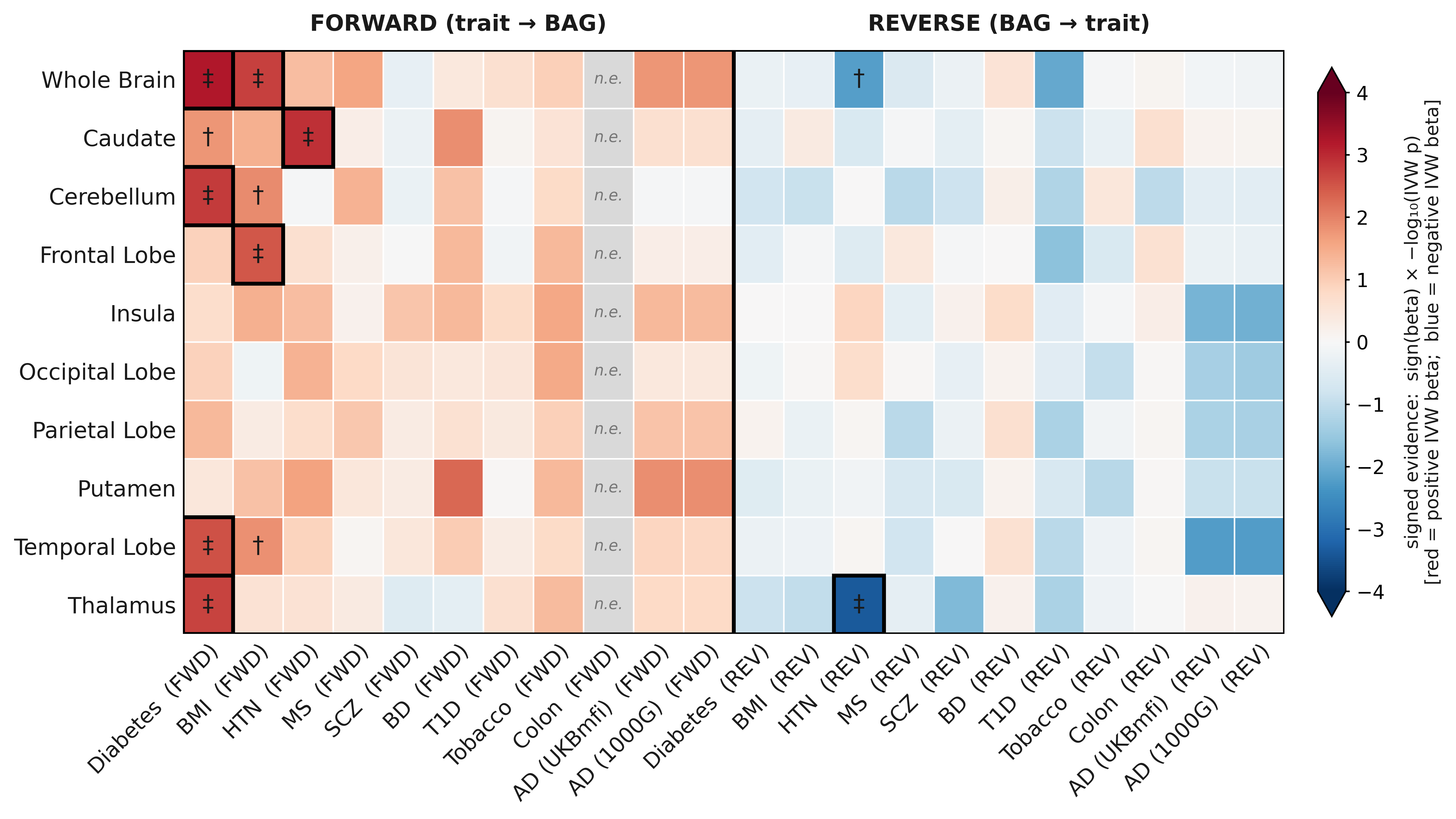

Supplementary Notes

Supplementary Note 1 | Disease phenotype definitions and diagnosis-level context

**Phenotype definitions.** Of the phenotypes selected for disease-association analysis, 24 had at least 30 cases in the independent UK Biobank test set and were retained for testing. Diagnoses were grouped for presentation into addiction-related, autoimmune, cancer, metabolic, neurological and psychiatric categories. Categories did not enter the statistical models or define multiple-testing families.

Composite phenotypes combined phecodes 290 and 331 for delirium, dementia, amnestic and other cognitive disorders and other cerebral degenerations; all 401 subcodes for hypertension; and 272.1, 272.11 and 272.12 for disorders of lipoid metabolism. The 290/331 composite includes delirium and other cognitive disorders in addition to dementia. Phecode 334 denotes degenerative disease of the spinal cord, and phecode 348 denotes other conditions of brain. Complete definitions and case counts are provided in Supplementary Table 3. In ADNI, the MCI group pooled early-MCI, MCI and late-MCI labels.

Test-set stratification and control selection. For test-set stratification, selected diagnoses were defined by the phecode criteria for the UK Biobank phenotypes in Supplementary Table 3 or by any of the following additional phecodes: 191.1, 295.1, 296.2, 301, 305.2, 312, 313, 316, 337, 338.2, 339, 343 and 695.4. Participants with no qualifying hospital inpatient record for any of these diagnoses were classified as without selected diagnoses (*n* = 10,776); participants with at least one qualifying record were classified as with at least one selected diagnosis (*n* = 9,836). The former group provided controls for the disease-association analyses. Control exclusion was applied irrespective of whether a diagnosis met the minimum case-count threshold for association testing.

**Expanded-control sensitivity analysis.** The sensitivity analysis retained the same cases and expanded the control group to 28,686 participants by adding 17,910 training participants without selected diagnoses. Training participants contributed out-of-fold predictions; sex restrictions and regression specifications were unchanged. Of the 171 FDR-significant primary associations, 160 remained significant and none reversed direction; one additional association reached FDR significance (Supplementary Table 3, continued).

Regional–whole-brain comparisons. For each UK Biobank diagnosis–region comparison, we used the test-set cases and controls from Supplementary Table 3. Each participant contributed a regional BAG and a whole-brain BAG, which were analysed together using linear regression with separate intercepts and coefficients for case status, age and sex:

$$Y_{it}=\alpha+\lambda R_{t}+\beta D_{i}+\delta R_{t}D_{i}+\gamma_{A}A_{i}+\eta_{A}R_{t}A_{i}+\gamma_{S}S_{i}+\eta_{S}R_{t}S_{i}+\varepsilon_{it},$$

where $Y_{it}$ is the BAG for participant $i$, $R_{t}=0$ denotes whole-brain BAG and $R_{t}=1$ denotes the regional BAG, $D_{i}=1$ for cases and $D_{i}=0$ for controls, and $A_{i}$ and $S_{i}$ denote age and sex. All coefficients were estimated separately for each diagnosis–region comparison. The whole-brain case–control coefficient is $\beta$, the regional coefficient is $\beta+\delta$, and $\delta$ estimates their difference. Two-sided tests of $H_{0}:\delta=0$ used standard errors clustered by participant to account for the paired observations. Both sex terms were omitted from sex-restricted analyses.

Comparisons were performed in years and repeated after dividing each BAG by its sample standard deviation in the corresponding controls:

$$Y_{it}^{*}=\frac{Y_{it}}{s_{t, C}},$$

where $s_{t,C}$ is the control-group standard deviation for the relevant BAG. Female-only and male-only control standard deviations were used for the breast- and prostate-cancer analyses, respectively. Benjamini–Hochberg correction was applied across the nine regional contrasts within each diagnosis, separately for the two scales.

On the year scale, 82 of 216 regional–whole-brain contrasts were significantly positive and four were significantly negative; 16 diagnoses had at least one significantly larger regional difference. On the control-standardized scale, one contrast was significantly positive and 58 were significantly negative. The positive contrast was observed for parietal BAG in multiple sclerosis (0.39 s.d.; 95% CI, 0.14–0.64; *q* = 0.012). Larger regional differences in years therefore did not generally correspond to larger standardized differences. Full contrast estimates, confidence intervals and P values are provided in Supplementary Table 3 (continued).

**Metabolic disorders (type 1 and type 2 diabetes).** Prior structural MRI studies have reported global and regional brain alterations in diabetes, including deep-grey and cortical changes^1–3^. These findings provide context for the prominent caudate, thalamic and putaminal BAG differences in type 1 diabetes and caudate, insular and putaminal differences in type 2 diabetes.

Neurological diagnoses and ADNI diagnostic groups. ADNI AD–CN differences were significant across all ten BAGs after adjustment for age, sex and MRI field strength (all *q* < 0.05; Supplementary Table 3). Previous neuropathological and imaging studies describe temporal and insular involvement and relative cerebellar preservation in AD^4–7^. The posterior cortical and deep-grey BAG differences observed in multiple sclerosis are also consistent with previous imaging reports^8–11^.

**Psychiatric and addiction-related diagnoses.** Larger BAG differences in bipolar disorder and addiction-related diagnoses commonly involved the insula and striatum. Previous studies of schizophrenia and bipolar disorder have reported subcortical abnormalities and widespread cortical alterations, including the insula^12–16^.

**Cancer diagnoses.** Associations were widespread for colon and female breast cancer and more restricted for prostate and lung cancer. Longitudinal imaging studies in breast and prostate cancer have reported treatment-associated brain changes^17,18^; the present estimates describe associations with recorded cancer diagnoses. This diagnosis-level literature also informed the disease-related traits selected for exploratory MR.

Supplementary Note 2 | Exploratory BAG GWAS in ADNI

We additionally performed exploratory GWAS of the ten BAG phenotypes in genetically inferred European-ancestry ADNI participants (*n* = 1,693; 7,910,956 variants).

**Genotyping and association analysis.** ADNI genotypes comprised five array datasets (ADNI1, ADNI GO/2 sets 1 and 2, and ADNI3 sets 1 and 2). After per-dataset quality control and strand alignment to the HRC reference, each dataset was imputed separately on the Michigan Imputation Server using the HRC r1.1 2016 European reference panel with Eagle phasing on GRCh37; ADNI1 genotypes were lifted over from hg18 before imputation. Variants with imputation *R*^2^ > 0.3 in all five datasets were retained, yielding 11,943,890 shared sites. These were merged and further restricted to biallelic single-nucleotide variants with minor allele frequency > 0.01, yielding 8,296,287 variants.

ADNI participants were assigned to a genetically inferred European-ancestry cluster by projection onto the 1000 Genomes reference principal-component space, after which principal components were recomputed within the retained ancestry subset. Sample quality control included filtering for cryptic relatedness (KING kinship coefficient > 0.0442) and heterozygosity outliers (mean ± 3 s.d.). The final genotyped sample comprised 1,856 participants, of whom 1,693 had a usable baseline MRI and were analysed. Association models were adjusted for age, sex, the first ten principal components computed within this subset, and four genotyping-batch indicators, with ADNI1 as the reference batch. The same set of 7,910,956 variants passing quality control and a minor allele count threshold of 10 was tested across all BAG phenotypes.

**Directional concordance.** ADNI effects were first harmonized to the UK Biobank effect allele in the common biallelic variant set, excluding variants with unresolved allele orientation. For each BAG, UK Biobank lead variants were then selected by lowest-*P* distance pruning within ±500 kb at *P* < 5 × 10⁻⁸ and, separately, *P* < 1 × 10⁻⁵. UK Biobank inputs preceded the post-GWAS MAF/INFO filter used for FUMA. Directional concordance was tested against a 50% null expectation using a two-sided binomial test. Pooled statistics were computed by summing region-specific lead sets and are interpreted as lead–region pairs rather than deduplicated variants.

**Results and interpretation.** Genomic-control inflation factors ranged from 0.97 to 1.02, indicating no overall inflation of the test statistics (Supplementary Table 8). Genome-wide significant variants were observed for four phenotypes only: whole-brain (7 variants), temporal-lobe (10), thalamic (3), and putaminal (1) BAG. All seven whole-brain variants and seven of the ten temporal-lobe variants lay in the chromosome 19 *APOE* region; the remaining three temporal-lobe variants formed a common-frequency locus at chr21:20.2 Mb (minor allele frequency 0.18). The thalamic and putaminal signals were low-frequency variants (minor allele frequency 2.6–5.7%) at chr5:176 Mb, chr4:57 Mb, and chr4:174 Mb, with limited local linkage-disequilibrium support, and were therefore not interpreted as established loci (Supplementary Table 7; Supplementary Fig. 5).

Within the *APOE* region, rs429358 showed concordant effect directions in the two cohorts: the T allele was associated with lower whole-brain BAG in UK Biobank (β = −0.052, P = 1.5 × 10^-7^) and ADNI (β = −0.226, s.e. = 0.037, P = 7.4 × 10⁻¹⁰), corresponding to higher BAG for the C allele carried on the *APOE* ε4 haplotype. The ADNI estimate was approximately fourfold larger, the 95% confidence intervals did not overlap, and the ε4 allele was more frequent in ADNI than in UK Biobank (0.264 versus 0.151). Diagnostic group was not included as a covariate in either GWAS. The concordant effect direction is consistent with a shared *APOE*–BAG association, while the larger ADNI estimate may partly reflect differences in diagnostic composition and *APOE* ε4 frequency. A different lead variant in the *APOE* region, rs814573, nevertheless reached genome-wide significance for whole-brain BAG in UK Biobank (*P* = 1.6 × 10^-8^; Supplementary Table 25, continued).

For whole-brain BAG, effect-allele frequencies were highly correlated between cohorts (r = 0.999), and harmonization changed neither effect orientations nor directional concordance (Supplementary Table 10). Between three and 24 lead variants were evaluated per BAG, with directional concordance ranging from 46.2% to 85.7%. At the genome-wide selection threshold, no phenotype-level test passed Bonferroni correction across the ten BAGs (smallest nominal *P* = 0.023). For whole-brain BAG, seven of 12 variants were concordant (58.3%). Across BAGs, 98 of 147 lead–region pairs were concordant (66.7%); this pooled summary is descriptive because the pairs are correlated across phenotypes (Supplementary Tables 9–10; Supplementary Fig. 6).

Supplementary Note 3 | Curation of GWAS Catalog trait categories for mapped-gene annotation

We annotated mapped gene groups using GWAS Catalog associations from release e0_r2022-11-29 supplied by FUMA. Reported traits were linked to Experimental Factor Ontology (EFO) terms and parent categories using the GWAS Catalog trait-mapping file (r2026-09-13, downloaded on 18 September 2026). The later trait-mapping release updated ontology assignments only; the underlying GWAS associations remained those in the FUMA-supplied 2022 release. For each gene group, we collected associated GWAS Catalog EFO terms and assigned each term to a top-level trait category. EFO terms already mapped to one of the predefined categories were retained as provided. For EFO terms labelled “Other measurement”, we applied an additional rule-based curation step to refine these broad entries into more informative categories.

The refinement used a deterministic keyword-to-category specification applied to case-normalized EFO strings (lower-case). Each EFO term was assigned to the first matching category according to an ordered list of keyword sets. The ordered mapping specification is reported in Supplementary Table 13, including the category name and the corresponding keyword list. If no keyword matched, the term remained in a residual “Other measurement” category. This refinement step was restricted to “Other measurement” terms only and did not alter EFO terms that were already assigned to a specific top-level category.

After refinement, we counted distinct reported traits within each mapped gene group and category. The 27-category summaries were used to compare groups detected in one BAG phenotype with those detected in two or more.

Supplementary Note 4 | Model architecture and training

The UK Biobank training set (*n* = 25,642) was selected by excluding participants with any record of neoplasm (phecodes 145–230), diabetes (249, 250), dementia and related cognitive disorders (290, 290.11, 291, 292), psychogenic, other mental and developmental disorders (303, 306, 315), intracranial haemorrhage and cerebrovascular disease (430, 433), other cerebral degeneration (331), or congenital anomalies of the nervous system (752, 753), following the criteria of Kim et al.^19^. The remaining 20,612 imaging participants formed the independent test set. Using the phecode criteria in Supplementary Note 1, prediction accuracy was summarized separately for participants without selected diagnoses (*n* = 10,776) and those with at least one selected diagnosis (*n* = 9,836; Supplementary Table 1). Participants without selected diagnoses also served as controls for the disease-association analyses, with sex restrictions for breast- and prostate-cancer comparisons (Supplementary Table 3).

Each model received a single-channel 128 × 128 × 128 input and used seven 3D convolutional blocks with batch normalization, six max-pooling operations and dropout. The resulting 768 features were flattened and passed through three fully connected layers with 96, 32 and 1 units; ReLU activations followed the two hidden layers^19^ (Supplementary Fig. 1). In both UK Biobank and ADNI, training–validation splits were generated using shuffled fourfold cross-validation with a fixed random seed. UK Biobank models were trained for up to 40 epochs per fold with mean squared error loss and AdamW optimization (learning rate 1 × 10^-4^, weight decay 5 × 10^-5^, batch size 8) and cosine annealing with warm restarts. ADNI fine-tuning used identical settings without cosine annealing. No augmentation was applied. Within each fold, the held-out fold was used to select the checkpoint with the lowest validation MAE and to obtain the corresponding fold-held-out predictions; it was not used for gradient-based parameter optimization. ADNI cross-validation performance estimates were therefore not independent of checkpoint selection, unlike the UK Biobank independent-test estimates (Supplementary Table 1). Models were implemented in PyTorch 2.4.0 under Python 3.10.15.

Supplementary Note 5 | Brain-age bias correction

For each UK Biobank model, age-related prediction bias was estimated from pooled out-of-fold predictions in the 25,642 training participants:

$P_{i}=\beta_{0}+\beta_{1}A_{i}+\varepsilon_{i}$,

where $P_{i}$ is uncorrected predicted age and $A_{i}$ is chronological age. The estimated intercept and slope were applied unchanged to all UK Biobank participants:

$BAG_{i}=\frac{P_{i}-\hat{\beta}_{0}}{\hat{\beta}_{1}} -A_{i}$.

Training participants contributed out-of-fold predictions, and test participants contributed predictions averaged across the four fitted models. In ADNI, cohort-specific correction parameters were estimated from out-of-fold predictions in cognitively normal participants and applied to all participants. This procedure follows the brain-age bias-correction framework of de Lange and Cole^20^.

Supplementary Note 6 | ProtAge-204 derivation, SNP heritability and ageing correlations

We constructed ProtAge-204 using the fixed 204-protein panel and published model specification of Argentieri et al.^21^, obtained from <https://github.com/miargentieri/proteomic-age-ukb> (commit eda0b4e9fbef3dd971570bb0bc8c47ee97db5c36). Protein selection was not repeated. Baseline UK Biobank Normalized Protein eXpression (NPX) data comprised 53,018 participants and 2,923 assays. Five participants absent from the study participant list were excluded. Assays with ≥50% missing values were removed, followed by participants with ≥54% missing values across the retained assays, leaving 52,315 participants and 2,920 assays. Missing values were imputed in a protein-only matrix using impute::impute.knn, with proteins as rows (k = 10, rowmax = 0.5, colmax = 0.95, maxp = 2,920; seed 362436069). Each protein was min–max scaled and median-centred; no z-standardization was applied. Participants with batch codes 0 or 7 or missing batch information were then excluded, leaving a clock cohort of 44,927. The published 204-protein panel was selected after imputation. Chronological age was calculated as days from the first day of the participant’s birth month to the baseline assessment date, divided by 365.25 and rounded to two decimal places. Chronological age was predicted using a LightGBM DART regression model with 5,000 boosting iterations.

Hyperparameters were selected using 200 Optuna trials with five-fold cross-validation in a fixed 70% development subset (*n* = 31,448; seed 3456). A model fitted in this subset was evaluated once in the remaining 30% (*n* = 13,479), yielding *r* = 0.935, R² = 0.875 and a mean absolute error of 2.28 years. The selected hyperparameters were also used to generate pooled five-fold out-of-fold predictions for all 44,927 participants (*r* = 0.936, R² = 0.877; mean absolute error, 2.26 years; root mean squared error, 2.88 years). Quality control, imputation and scaling preceded data partitioning, and tuning was not nested within the final out-of-fold partitions. These performance estimates are therefore descriptive rather than fully nested validation results. Neither genetic information nor downstream association outcomes were used in model selection.

The raw proteomic-age gap was defined as out-of-fold predicted proteomic age minus chronological age. Age-related prediction bias was removed using an analogous linear calibration framework to that used for the brain-age phenotypes (Supplementary Note 5), with the corrected phenotype defined as the residual from a regression of the raw proteomic-age gap on chronological age. Under the fitted linear model, this residual correction differs from the division-based BAG correction only by a positive scaling constant and therefore preserves participant ranks and yields the same phenotype after rank-based inverse-normal transformation. The transformation was performed in the complete out-of-fold sample of 44,927 participants. Of these, 39,114 self-reported their ethnic background as British (UK Biobank Data-Field 21000, instance 0) and were considered for the genotype intersection described below. Higher values indicate greater age-bias-corrected proteomic-age acceleration.

Genome-wide association analysis used SAIGE v1.1.9 and the LD-pruned Step 1 genotype set used for the BAG GWAS. Among the 39,114 self-reported British participants, 38,857 were present in this genotype set and 38,806 were also present in the imputed BGEN sample. The latter participants were used in both Step 1 and Step 2. Models were adjusted for granular chronological age, sex and the first ten genetic principal components, with a leave-one-chromosome-out approach. Variants with minor allele count below 10 were excluded, and testing used GRCh37 coordinates. SAIGE’s inverse-normalization option was enabled.

For LDSC, merged BAG summary statistics were restricted to analysis-sample MAF >0.01 and HapMap3, without applying the post-GWAS INFO filter. A1 was the effect allele (SAIGE Allele2). After merging with the LD-score panels, 1,176,210 variants were retained for each BAG; counts for the two external ageing traits differed slightly. BAG heritability estimates are reported on the inverse-normal-transformed BAG scale. Heritability estimates were obtained with the two-step LD Score regression estimator (χ² cutoff 30), whereas the cross-trait analysis uses the one-step estimator; the heritability values in Supplementary Table 14 therefore differ slightly from the denominators of the genetic correlations in Supplementary Table 15. Across the ten BAGs, LDSC intercepts ranged from 1.043 to 1.056 and attenuation ratios from 0.196 to 0.335 (Supplementary Table 14); residual confounding cannot be excluded.

**Regional–whole-brain heritability comparisons.** For each of the nine regional BAGs, we calculated the difference between its LDSC heritability estimate and the whole-brain estimate, using the two-step settings described above. Standard errors were obtained by a paired block jackknife using 200 shared genomic blocks to account for covariance between estimates. Two-sided P values were adjusted across the nine comparisons using the Benjamini–Hochberg procedure (Supplementary Table 14, continued).

SNP-based heritability and LDSC diagnostic statistics for the ten BAG phenotypes, telomere length PC1, and ProtAge-204 are reported in Supplementary Table 14. Cross-trait genetic correlations comprised 45 pairwise BAG–BAG comparisons, ten telomere length PC1–BAG comparisons, ten ProtAge-204–BAG comparisons, and one ProtAge-204–telomere length PC1 comparison.

Benjamini–Hochberg correction was applied separately to the 45 BAG–BAG genetic correlations and jointly to the 21 external-ageing genetic correlations.

BAG–BAG phenotypic correlations were unadjusted Pearson correlations between inverse-normal-transformed BAGs in 41,067 participants, with Benjamini–Hochberg correction across 45 pairs. ProtAge-204–BAG phenotypic correlations were calculated between bias-corrected gaps before inverse-normal transformation in the 3,784 participants with both measurements, adjusting for sex, age at protein sampling and the protein–MRI interval. Benjamini–Hochberg correction was applied across the ten BAGs. All ten partial correlations were positive (*r* = 0.081–0.149; all *q* < 1 × 10⁻⁶; Supplementary Table 15). Protein sampling preceded MRI by a mean of 8.60 years (s.d. 2.01; range, 3.45–14.19 years). These phenotypic analyses used a smaller sample and non-contemporaneous measurements, whereas the genetic-correlation analyses used summary statistics from the 38,806-participant ProtAge-204 GWAS.

Supplementary Note 7 | Cell-type enrichment analysis: dataset composition, thresholds, and cell-class harmonization

**Implementation.** Nine BAG phenotypes were analysed using the FUMA web implementation. Cerebellar BAG was analysed locally using the unmodified FUMA script, with the same expression datasets, model specifications and multiple-testing settings. For phenotypes analysed using both implementations, marginal gene-property statistics agreed.

**Datasets and missing values.** The single-cell expression datasets distributed with FUMA^22^ comprise human and mouse data spanning multiple anatomical regions and developmental stages. Each BAG phenotype had 12,531 listed dataset–cell-type annotations, of which 12,522 had valid marginal P values; the same nine annotations were missing for all ten BAGs. Missing results were neither imputed nor treated as non-significant.

**Thresholds and retention.** Step 1 comprised one-sided marginal gene-property tests for positive enrichment, with Bonferroni correction across the 12,522 valid tests within each BAG (*P* < 3.99 × 10^-6^). When multiple annotations in a source dataset were significant, Step 2 evaluated their conditional associations using FUMA’s retention criteria. Of 173 significant phenotype–annotation associations, 142 were retained: 123 had no other significant annotation in the same source dataset, including the sole thalamic association, and 19 met the conditional retention criteria. The remaining 31 were not retained (Supplementary Table 16).

Step 3 comprised pairwise conditional analyses across source datasets for 141 retained annotations. The sole thalamic annotation did not enter Step 3 because no Step 2 analysis was performed for that phenotype. Step 3 assessed cross-dataset dependence and did not alter retention.

**Cell-class harmonization.** After all statistical testing, fine-grained source labels were mapped to 15 canonical classes: excitatory neuron, inhibitory neuron, other neuron, astrocyte, oligodendrocyte, oligodendrocyte precursor cell (OPC), other glia, microglia, progenitor, endothelial, fibroblast/VLMC (vascular leptomeningeal cell), pericyte/mural, vascular (unspecified), non-neuronal, and unresolved. Seven of the 15 classes occurred among retained annotations. Harmonization did not alter the MAGMA statistics or retention status. Neuronal labels lacking excitatory or inhibitory subtype resolution were assigned to other neuron; glial labels lacking a specific lineage to other glia; explicitly non-neuronal labels lacking further resolution to non-neuronal; generic vascular labels lacking subtype resolution to vascular (unspecified); and labels without a resolvable lineage to unresolved. The unresolved cerebellar association retained the source label “unknown” from the human substantia nigra dataset 426_Kamath2022_Human_2022_SN_level1; it was not reassigned to a lineage.

**Neuronal classes.** Annotations assigned to each of the three neuronal classes were tested for every BAG phenotype, but none was retained.

Supplementary Table 16 lists all 142 retained annotations with their source labels, harmonized classes, retention status, and Step 3 statistics. Counts represent dataset–cell-type enrichment signals rather than distinct cell populations or independent replications.

Supplementary Note 8 | ADNI biomarker, cognitive and diagnostic-stage analyses

**Biomarker definitions and visit matching.** Amyloid PET used the UC Berkeley 6-mm summary cortical pipeline. The data provider normalized summary cortical SUVR to the whole cerebellum and converted measurements from florbetapir, florbetaben and NAV4694 scans to Centiloids. CSF analyses included Roche Elecsys measurements of Aβ42 and p-tau181 and excluded measurements from the earlier INNO-BIA AlzBio3 assay.

Among MRI scans designated as baseline, the earliest acquisition for each participant was retained, yielding 2,986 participants. MRI–measure matching used the nearest visit within the modality-specific windows specified in Methods. Median absolute intervals were 30 days for amyloid PET, 44 days for tau PET, 27 days for both CSF measures and 14–18 days for cognitive measures. *APOE* ε4 allele count was unavailable for 540 participants. Complete-case sample sizes ranged from 684 for tau PET to 2,442 for MMSE (Supplementary Table 4).

**Association models.** Each BAG was standardized using its mean and sample standard deviation (ddof = 1) in the full ADNI baseline sample (*n* = 2,986). These constants were used in both the single-BAG and regional-plus-whole-brain models and held fixed across the primary and sensitivity PET analyses. For each BAG-outcome pair, we fitted

$M_{i} = \beta_{0}+\beta_{1}BAG_{i}+\beta_{2}age_{i}+\beta_{3}sex_{i}+\beta_{4}E4_{i}+\epsilon_{i}$,

where $M_{i}$ denotes the AD-related measure, ${BAG}_{i}$ is the standardized BAG trait and ${E4}_{i}$ is the *APOE* ε4 allele count for participant $i$. Associations in Fig. 2b are expressed as partial correlations adjusted for age, sex and *APOE* ε4 allele count. Partial correlations were obtained from the t statistic for the standardized BAG coefficient as $r=\mathrm{sign}(t)\sqrt{t^{2}/(t^{2}+\mathrm{df}_{\mathrm{resid}})}$ , where $df_{\mathrm{resid}}$ is the residual degrees of freedom. Their 95% confidence intervals were calculated on the Fisher z scale using $SE=1/\sqrt{N-k-3}$, where $N$ is the complete-case sample size and k = 3 is the number of covariates partialled out (age, sex and *APOE* ε4 allele count).

To assess regional associations after adjustment for whole-brain BAG, we fitted a separate model for each regional BAG and outcome:

$M_{i} = \beta_{0}+\beta_{R}BAG_{R,i}+\beta_{W}{BAG}_{W,i}+\beta_{2}{age}_{i}+\beta_{3}{sex}_{i}+\beta_{4}{E4}_{i}+\epsilon_{i}$,

where $BAG_{R, i}$ and $BAG_{W, i}$ are the standardized regional and whole-brain BAGs. The regional coefficient assessed the association conditional on whole-brain BAG. The two BAG coefficients were compared by testing $H_{0} :\beta_{R}-\beta_{W} = 0$, accounting for their covariance. Regional coefficients and coefficient contrasts were evaluated in separate Benjamini–Hochberg families of 63 tests each.

Diagnostic groups. Diagnostic-stage analyses combined CN with SMC (n = 1,263) and early MCI with MCI and late MCI (n = 1,303); the AD group comprised 420 participants. These counts refer to the full baseline sample before analysis-specific exclusions. SMC participants were not included in the 1,179 CN participants used for model fine-tuning or in the CN comparator for the ADNI BAG comparisons in Supplementary Table 3.

**Tau PET analyses.** Primary tau-PET analyses were restricted to provider-quality-control-passed 6-mm flortaucipir scans. Tracer and quality-control eligibility were applied before MRI matching, and the nearest eligible scan within 365 days was selected, yielding 684 participants with complete data. Because SUVRs are not directly comparable across tau tracers, the pooled analysis including flortaucipir, MK6240 and PI2620 was retained as a sensitivity analysis. For amyloid PET, sensitivity analyses were restricted to scans passing the provider’s quality-control review (*n* = 1,544; Supplementary Data 1).

Because UK Biobank pretraining used 3 T MRI and field strength varied across ADNI phases, the models containing regional and whole-brain BAGs and the diagnostic-stage analyses were repeated in participants whose MRI used for BAG estimation was acquired at 3 T. Participants with unrecorded field strength were excluded, and the corresponding multiple-testing corrections were repeated within the restricted sample (Supplementary Tables 5 and 6).

Supplementary Note 9 | Discrimination of observed MCI-to-AD conversion

**Cohort definition and follow-up.** Visit-level DXSUM diagnoses were ordered by examination date and linked to MRI visits by visit code or, when unavailable, by the nearest examination within 183 days. Baseline was each participant’s first diagnosis-matched MRI visit, and eligibility required MCI at that visit. This definition identified 1,251 participants before subsequent exclusions; the 1,303 MCI participants in the imaging-cohort summary were grouped using baseline T1-series diagnoses. Converter classification required an AD diagnosis at a subsequent MRI-matched visit. Time to conversion was calculated to the first AD diagnosis recorded in DXSUM, irrespective of MRI availability at that visit. The 24 converters with a diagnostic reversion at some point during follow-up remained classified as converters.

We excluded 70 participants whose diagnoses reverted without reaching AD, 47 with AD recorded in DXSUM but no MRI at the corresponding visit, 33 whose diagnoses varied during follow-up without reaching AD, and 80 with no follow-up visit after baseline. The final sample comprised 303 converters and 718 non-converters. Among converters, median time to AD diagnosis was 2.1 years (interquartile range, 1.1–3.2). Among non-converters, median diagnostic follow-up was 2.1 years (interquartile range, 0.3–4.2); 32.0% had less than one year and 48.1% had less than two years of follow-up. Cohort flow and follow-up distributions are provided in Supplementary Data 1.

**Model fitting and evaluation.** The whole-brain model included age, sex and whole-brain BAG; the region-selected model included age, sex and selected regional BAGs. Performance was evaluated using stratified tenfold outer cross-validation with shuffled splits and a fixed random seed. Within each outer-training fold, fivefold inner cross-validation evaluated k = 1–9. Regions were ranked within each training split by the univariate AUC of regional BAG residuals calculated using whole-brain adjustment coefficients estimated in cognitively normal participants. Residuals were used only for ranking; bias-corrected regional BAGs were entered into the prediction models. The k with the highest pooled inner-validation AUC was selected, after which regions were reranked and the model refitted using the complete outer-training fold. Standardization parameters were estimated from training data only. Outer-test participants were used only for prediction, and AUCs were calculated from predictions pooled across the ten outer folds. Fixed-k results are reported descriptively and were not used to select the primary reported model.

Uncertainty in the AUC difference. We defined ΔAUC as AUC of the region-selected model minus that of the whole-brain model. Percentile 95% confidence intervals were obtained from 1,000 participant-level bootstrap replicates. Conditional on the precomputed BAGs, each replicate repeated the complete conversion-model cross-validation procedure, including region ranking, selection of  k, standardization, fitting and held-out evaluation. Repeated copies of the same participant were assigned to the same fold at every level of cross-validation. Both models were evaluated using the same resampled participants and outer folds.

**Sensitivity analyses.** A ridge model included all nine regional BAGs without region selection; the penalty was selected by threefold cross-validation within the training data. A separate follow-up sensitivity analysis retained the 373 non-converters with at least 24 months of follow-up and all 303 converters, yielding 676 participants. The same evaluation and bootstrap procedures were applied. Fixed-k and ridge-model results are shown in Fig. 2c; all model comparisons are reported in Supplementary Data 1.

Supplementary Note 10 | UK Biobank GWAS and lead-variant annotation

**Genotypes and association testing.** GWASs of the ten BAG traits were performed in 41,067 White British UK Biobank participants using SAIGE v1.1.8 for null-model fitting and v1.1.9 for association testing^23,24^. Association testing used the UK Biobank centrally imputed release v3 on GRCh37, imputed using the Haplotype Reference Consortium and UK10K/1000 Genomes reference panels^25^.

The null-model input comprised 215,514 LD-pruned, directly genotyped variants in 488,377 participants, with fitting restricted to the relevant analysis sample. SAIGE retained variants with a minor allele frequency of at least 0.01 and a missingness rate of at most 15%, yielding 111,914 GRM variants in the 41,067-participant discovery sample and 111,956 in the 30,800-participant PRS-derivation sample. Relatedness was modelled using a dense genetic relationship matrix with a leave-one-chromosome-out approach.

For each BAG, rank-based inverse-normal transformation was performed jointly across the 41,067 discovery participants. The PRS-derivation GWAS used the same procedure, with transformation performed separately within its 30,800 participants. The association-testing inputs contained 93,095,623 variants across the 22 autosomes and had not undergone study-specific filtering by imputation quality. Step 2 applied a minor allele count threshold of 10 and a maximum missingness rate of 15% within each analysis sample, without an additional MAF or INFO threshold. Following association testing, variants were retained for locus definition and annotation if they matched one UK Biobank mfi record by GRCh37 chromosome, position and allele pair, had mfi INFO ≥0.8 and had analysis-sample MAF ≥0.01. This retained 9,458,475 of 35,784,891 tested variants per BAG. Effect estimates, standard errors and P values were unchanged (Supplementary Table 25). Additive variant effects were tested using the following equation:

$y_{i} = \beta_{0}+\beta_{G}\cdot G_{i}+C_{i}\gamma+u_{i}+\varepsilon_{i}$,

where $y_{i}$ is the transformed BAG; $G_{i}$ is the imputed allele dosage; $C_{i}$ are covariates including age, sex and the first ten genetic principal components; $u_{i}$ is the polygenic random effect; and $\varepsilon_{i}$ is the residual error. Association tests used imputed dosages with the alternate-allele-first coding of the UK Biobank imputation v3 BGEN files. Effect estimates refer to SAIGE Allele2, and AF_Allele2 is its frequency in the analysis sample.

**Lead-variant and locus definition.** FUMA^26^ used the 1000 Genomes Phase 3 European reference panel on GRCh37. Independent significant variants were defined at *P* < 5 × 10^-8^ and *r*^2^ < 0.6; lead variants at *r*^2^ < 0.1. Candidate variants met *P* < 0.05, and *r*² ≥0.6 with an independent significant variant. Signals within 250 kb were merged into genomic risk loci. Gene annotation used Ensembl v102. FUMA's reference-panel minimum MAF was 0; uploaded variants had already passed the sample-MAF filter. For all ten BAGs, the default MHC interval (GRCh37 chr6:29,614,758–33,170,276) was excluded from FUMA annotation, including lead-variant and genomic-risk-locus definition (exMHC = 1; MHCopt = annot; extMHC = NA). This exclusion did not apply to GWAS association testing or MAGMA gene analysis. Across phenotypes, overlapping risk-locus intervals or intervals separated by ≤250 kb were merged (Supplementary Table 25).

**GWAS Catalog annotation.** Lead variants were annotated using the NHGRI-EBI GWAS Catalog release e0_r2022-11-29 supplied by FUMA. Across the ten BAGs, 42 distinct lead variants had Catalog annotations. Within each MappedGene annotation, repeated gene names were removed, names were sorted and intergenic separators standardized; hyphens within gene symbols were preserved. The resulting 35 mapped gene groups represent annotations rather than genomic intervals; a gene may occur in more than one group. Lead variants were pooled across the ten BAG GWASs. A group was detected in a BAG when at least one variant from this pooled set carried the group annotation in that BAG’s FUMA candidate-variant results. The variant did not need to be a lead variant for that BAG. Groups detected in one or multiple BAGs were classified as phenotype-restricted or shared, respectively (Fig. 3b; Supplementary Table 11).

Supplementary Note 11 | Polygenic risk score construction and cross-cohort PheWAS

**Score construction and target data.** Each released BAG PRS weight set contained 1,154,522 variants and was derived in 30,800 UK Biobank participants. Scores were calculated as weighted sums of allele dosages using the scored allele specified in each weight file. In the 9,997 relatedness-pruned held-out participants, single-score validation used univariable linear regression of inverse-normal-transformed BAG on its corresponding PRS. Coefficients are reported per 1-s.d. higher PRS, together with adjusted R² (Supplementary Table 17). The interval chr6:28–33 Mb was retained during PRS construction. Within this interval, no variant had a posterior inclusion probability greater than 0.9, and the largest absolute posterior effect estimate was 0.003.

**Exploratory comparison of one- and ten-PRS models.** For each inverse-normal-transformed BAG, the reduced linear model included age, sex, ten genetic principal components and the corresponding PRS; the full model included all ten BAG PRS. Scores were standardized within the 9,997-participant relatedness-pruned held-out sample. We reported increases in unadjusted model R² relative to the covariates-only model and compared the one- and ten-PRS models using their R² difference and a partial F-test of the nine added scores. Models were fitted and evaluated in the same sample; these analyses did not assess out-of-sample predictive improvement (Supplementary Table 17, continued).

**Target samples and All of Us data.** Primary PheWAS used relatedness-pruned White British UK Biobank non-imaging and held-out imaging samples, and genetically inferred European-ancestry All of Us participants. UK Biobank non-imaging participants were outside the 46,254-participant imaging cohort and met the same self-reported British criterion used for PRS derivation. All of Us analyses used the Controlled Tier Curated Data Repository v7 in the Researcher Workbench, accessed in August 2026. Scores were computed from the short-read whole-genome sequencing ACAF threshold callset, converted to PLINK 2.0 format by chromosome. Score variants were lifted over from GRCh37 to GRCh38 and matched by chromosome, position and alleles (Supplementary Tables 18–20 and 24).

**UK Biobank relatedness pruning.** Participants in the 30,800-person PRS-derivation sample and target participants related to them at KING kinship > 0.0884 were excluded. Within each target sample, the participant with the most remaining related pairs was then iteratively removed until no pair exceeded this threshold. Ties were resolved by a fixed random priority (seed 20260930), without phenotype information. In the non-imaging sample, 4,208 participants were removed for relatedness to the derivation sample and 25,503 by within-target pruning, leaving 359,334. The corresponding exclusions were 239 and 31 in the held-out imaging sample, leaving 9,997 (Supplementary Table 24).

**All of Us relatedness pruning.** We excluded participants on the v7 precomputed relatedness-pruning list. Of 116,656 participants with phenotype and PRS data, 4,576 were removed, leaving 112,080; no pair in the supplied relatedness file remained within this sample. Excluding 2,306 participants with missing sex left 109,774 with complete covariates before phenotype-specific exclusions and sex restrictions (Supplementary Table 24).

**Phecode definitions.** Phecode definitions and the ICD-10-to-phecode map were obtained from the PheWAS R package (<https://github.com/PheWAS/PheWAS>), using phecode v1.2 (<https://phewascatalog.org/>). Phecode names and categories were assigned using an export of the PheWAS package’s pheinfo table. Results without a resolved name were retained under their phecode identifiers. Integer-parent groupings were used to combine figure labels. Association testing and multiple-testing correction retained each tested phecode, including separately tested parent codes. UK Biobank ICD-10 records were obtained from Data-Field 41270 and extracted on 18 August 2022. In both cohorts, records were mapped to phecodes v1.2 and aggregated to the first decimal place. Cases required at least one qualifying record, and PRS–phenotype tests required at least 30 cases.

Related-disease exclusions were applied to controls in both UK Biobank samples and in All of Us. UK Biobank used the PheWAS-generated phenotype matrix; All of Us applied exclusion ranges from the phecode v1.2 definitions to case lists, propagating the ranges within aggregated phecode blocks. The definitions-based implementation was slightly more restrictive in the comparison with the UK Biobank matrix.

All UK Biobank PheWAS analyses used age calculated as 2018 minus year of birth. In All of Us, age was calculated as the number of elapsed days between the recorded date of birth and 1 January 2023, divided by 365. For phecodes with an unambiguous sex designation, analyses were restricted to the corresponding sex and omitted sex as a covariate. Phecode 751.1 remained unrestricted because its constituent subcodes had conflicting sex designations. The equation below gives the model for unrestricted analyses; the sex term was omitted in restricted analyses.

**Association models and excluded tests.** For each BAG PRS $r$, we fitted the following model:

${logit Pr(y}_{i} = 1) = \beta_{0}+\beta_{1}\cdot{PRS}_{i,r}+\beta_{2}\cdot{age}_{i} +\beta_{3}\cdot{sex}_{i}+\sum_{k=1}^{10} \gamma_{k}\cdot PC_{i, k}$,

where $y_{i}$ denotes case status for the phecode under analysis, $PRS_{i, r}$ is participant $i$’s unstandardized summed score for BAG phenotype $r$, and $PC_{i, k}$ is the $k$th genetic principal component. Genetic principal components were computed within the All of Us European-ancestry subset.

Models used participants with finite PRS and covariates and an eligible case–control status. Tests with fewer than 30 cases after exclusions, a single outcome category, nonconvergence or fitting errors were excluded. There were 13,440 successful tests in the pruned UK Biobank non-imaging sample, 4,960 in the pruned held-out imaging sample and 15,151 in pruned All of Us. All of Us model-specific sample sizes ranged from 26,862 to 109,774. Benjamini–Hochberg correction was applied separately within each BAG PRS and sample. Significant associations and phecode-level summaries are reported in Supplementary Tables 18–20.

Supplementary Note 12 | Bidirectional Mendelian randomization

**Instrument selection and harmonization.** Variants reaching *P* < 5 × 10^-8^ in the exposure GWAS were clumped using PLINK 2.0 at *r*² < 0.001 within 10 Mb, with UK Biobank imputed genotypes restricted to HapMap3 variants as the LD reference. The diabetes GWAS was ukb-b-10753 (UK Biobank Data-Field 2443, “Diabetes diagnosed by doctor”), a self-reported binary phenotype without restriction by diabetes type. This phenotype differed from the type 2 diabetes phecode used in the UK Biobank disease-association and PheWAS analyses. Exposure and outcome summary statistics were harmonized using the default TwoSampleMR procedure. Palindromic variants were oriented using allele frequencies, and variants that could not be oriented unambiguously were excluded. For AD, variant effects and standard errors were approximated from z-scores using $\beta=\frac{z}{\sqrt{2f(1-f)(N + z^{2})}}$ and $SE = \frac{1}{\sqrt{2f(1-f)(N + z^{2})}}$, where $f$ is the reference allele frequency and $N$ is the per-variant sample size. UK Biobank mfi frequencies were used for the primary reconstruction and 1000 Genomes European frequencies for the sensitivity reconstruction. No liability-scale transformation was applied. Per-variant instrument strength was calculated as $F=\left( \frac{\beta_{x}}{SE\left[ \beta_{x} \right]} \right)^{2}$, where $\betaₓ$ is the variant–exposure effect estimate. No F-statistic threshold was applied. Across instruments used for the ten BAG exposures in reverse MR, the minimum F-statistic was 30.0, and none was below 10.

**Estimation and sensitivity analyses.** Analyses used TwoSampleMR v0.6.16 and MR-PRESSO v1.0 in R 4.3.3. IVW was the primary estimator. Weighted-median and MR–Egger estimates, Cochran’s Q heterogeneity tests, MR–Egger intercept tests, MR-PRESSO, Steiger directionality tests and leave-one-out analyses were obtained where possible. MR-PRESSO was initially run with 1,000 simulations and was restricted to exposure–outcome pairs with at least four instruments. Fifty-nine comparisons with an initially significant MR-PRESSO global test and insufficient simulation resolution to obtain a Bonferroni-adjusted outlier-test *P* < 0.05 were rerun with 20,000 simulations (seed 20261001). The BMI-to-whole-brain and BMI-to-frontal-lobe comparisons were additionally repeated with seed 20261002 to assess Monte Carlo stability. Supplementary Data 2 reports the outlier counts and corrected estimates from seed 20261001. Both comparisons failed the prespecified stability criteria because outlier sets differed and between-seed changes in corrected estimates exceeded 10% of their standard errors. Corrected estimates remained positive with nominal *P* < 0.05, and global-test *P* values remained <5 × 10^-5^, in both runs. Reverse IVW analyses used 2–17 instruments per BAG–trait pair; MR–Egger and MR-PRESSO estimates were unavailable for pairs with two instruments. Multiple-testing correction was applied across the ten BAGs within each disease-related trait and direction (Benjamini–Hochberg q < 0.05; Bonferroni *P* < 0.05/10 = 0.005).

Steiger directionality. Directionality was evaluated on the scale represented by the available exposure summary statistics. For UK Biobank binary traits analysed using linear-probability models (doctor-diagnosed diabetes, hypertension, ever smoked and colon cancer), variance explained was calculated on that observed scale. Binary log-odds traits without the prevalence information required for liability-scale conversion were marked as not evaluable rather than transformed using an assumed prevalence.

**Interpretation and reporting.** MR findings were considered exploratory because of sample overlap between most disease-related GWASs and the UK Biobank BAG GWASs, the small number of reverse-direction instruments and differing exposure scales. GWAS sources, instrument metadata, MR estimates and sensitivity diagnostics are provided in Supplementary Data 2. Supplementary Fig. 10 shows prioritized forward estimates, and Supplementary Fig. 11 summarizes evidence in both directions. Across the trait–BAG comparisons, eight IVW associations passed Bonferroni correction and four additional associations passed FDR correction. MR–Egger intercept tests were nominally significant in 16 of 199 estimable comparisons; these diagnostic P values were not adjusted for multiple testing. Hypertension showed a positive forward association with caudate BAG and a negative reverse association with thalamic BAG at the Bonferroni threshold. For the reverse thalamic comparison, the weighted-median estimate was not significant (P = 0.100), and the MR–Egger intercept test was nominally significant (P = 0.030). These regionally distinct findings and sensitivity diagnostics do not establish a consistent bidirectional causal relationship. No type 1 diabetes IVW association passed either correction. Forward IVW estimates for colon cancer were not obtained because only one independent instrument remained (Supplementary Data 2; Supplementary Fig. 11).
